# Concurrence of Dementia and Mental Disorders Across Populational Epidemiology and Global Disease Burden Perspectives

**DOI:** 10.64898/2026.04.28.26351932

**Authors:** Haoran Zhang, Ting Pang, Yingqi Liao, Xiaowen Lou, Wanheng Zhang, Peige Song, Haoxuan Wen, Zhipeng Xue, Brian Hall, Xin Xu

**Author notes:** **Corresponding author:** Xin Xu, PhD, School of Public Health, the Second Affiliated Hospital of School of Medicine, Zhejiang University, China, Postal address: No. 866, Yuhangtang Road, Hangzhou, Zhejiang, China. 310058 Email address.

## Abstract

**Background:** Dementia and mental disorders are leading causes of disease burden in middle-aged and older adults. Existing evidence suggests close clinical and epidemiological associations between dementia and mental disorders. We aimed to characterize the concurrent features of dementia and eight mental disorders at both the individual level and the population level (temporal and spatial patterns of global burden of disease co-existence and co-occurrence).

**Methods:** The study utilized data from the UK Biobank (UKB, N=502461, 2006-2021) and Global Burden of Disease (GBD, 204 countries, 1990-2021). In UKB, Cox models were used to explore associations between mental disorders and dementia and shared risk factors between the two conditions. In GBD, multi-state Markov models were constructed to quantify the expected time of concurrent burden.

**Results:** There was a substantial concurrence between dementia and mental disorders, along with causal associations and shared risk factors at the individual level. At the country level, although a trend of decreased prevalence and incidence of dementia was observed in most regions over the past 31 years, dementia demonstrated greater co-existence burden of neurodevelopmental disorders (especially attention-deficit/hyperactivity disorder and idiopathic developmental intellectual disability) in Middle East and North Africa. In high-socio-demographic index areas such as Western Europe, and high-income north America countries, it was more linked to mental disorders including anxiety disorders, eating disorders, and schizophrenia. There was a high co-existence burden of neurodevelopmental disorders demonstrated with both early- and late-onset of dementia.

**Conclusions:** Our findings underscore the urgent need for proactive, integrated, and life-course health management strategies to achieve precision prevention and control of dementia, mental disorders and their concurrence.

## 1 Introduction

Neurodegenerative diseases such as dementia are leading causes of disease burden in middle-aged and older adults.[1,2] The number of people of living with dementia worldwide is set to increase over time, especially in lower-income countries, and is projected to increase to 153 million by 2050.[3] Meanwhile, mental disorders, such as depressive disorders (DD) and anxiety disorders (ANX), are increasingly recognised as the leading cause of disease burden.[2] Despite ample research confirming the effectiveness of early detection and intervention in reducing the impact of dementia and mental disorders, improvement on the global burden of both remains a major challenge. [2,3]

Existing evidence suggests close clinical and epidemiological associations between dementia and mental disorders. At the individual level, population-based studies have reported elevated concurrence between dementia and mental disorders, such as schizophrenia,[4] bipolar disorders,[4] and depression.[5] One possible explanation for this co-occurrence is a causal relationship, as mental disorders have been associated with an increased risk of subsequent dementia.[6]

Shared risk factors may represent another potential contributor. Accumulating evidence has revealed overlapping risk factor profiles between dementia and mental disorders, encompassing lifestyle factors (e.g., smoking),[7] metabolic diseases,[8] and shared genetic susceptibility.[3,9] Such overlapping clinical manifestations and risk factors may drive their concurrence ^①^ at both the individual and population levels. However, few studies have systematically estimated these concurrent features within a unified framework, particularly from global perspective.

In light of the recent research development, ongoing changes in social environments and structures, the severity and trajectory of such concurrent burden is likely to be undergoing dynamic fluctuation and exhibiting geographic heterogeneity.[2,10] For instance, studies in high socioeconomic regions have reported a decline in the dementia incidence,[10,11] but an increase in mental disorders prevalence.[2] However, most studies in dementia and mental disorders have focused on single diseases within homogeneous populations.[2,12] Thus, there remains an important research gap in investigating the disorder-specific trends of concurrent temporal pattern and spatial heterogeneity utilising longitudinal and country-level data.

The present study used the data from the UK Biobank (UKB) and Global Burden of Disease (GBD) database 2021 to characterize the concurrent burden between dementia and mental disorders from individual and global perspectives, separately. At the individual level, we summarized the prevalence and diagnostic order of dementia and mental disorders, and examined the associations and shared risk factors between these two conditions. At the country level, we analyzed prevalence and incidence of dementia and mental disorders, and their temporal and spatial individual and concurrent burden across different countries and territories. Given the complex and heterogeneous impact of the later stages of the COVID-19 pandemic on mental health, we restricted our analysis to data up to 2021.[13] Additionally, as aging is an important risk factor for the development of both dementia and mental disorders,[14,15] comparison in the associations between early- or late-onset of dementia with mental disorders was explored in the current study. We sought to better reveal the concurrent burden of these two diseases at both individual and country levels, and to provide evidence-based support for making global dementia and mental health policies and optimizing resource allocation.

## 2 Methods

### 2.1 Overview

The study used data from the UKB and GBD 2021 database. The UKB is a population-based cohort from the United Kingdom. Over 500, 000 participants aged 37-73 years old underwent a comprehensive baseline assessment (2006-2010).[16] We excluded participants with baseline dementia, and further excluded those with baseline mental disorders for the risk factor analysis, thereby restricting the analysis to those at risk of incident disease. Given the predominant focus on mental disorders in the pre dementia phase and the limited number of participants with baseline dementia, we did not examine the reverse association (dementia to incident mental disorders).[8] All study participants signed informed consent before the study. The study was approved by the Institutional Review Board. From the GBD database, the analysis included prevalence estimates and incidence rates for individuals aged 40 years and older across 21 regions and 204 countries and territories from 1990 to 2021 (GBD does not provide dementia estimates for populations under 40).

### 2.2 Definition of dementia and mental disorders

In this study, we adopt ‘concurrence’ as a unifying framework, comprising individual-level concurrence in the UKB and country-level measures of prevalence-based co-existence and incidence-based co-occurrence of dementia and mental disorders in the GBD database.

We included dementia and eight mental disorders subtypes including schizophrenia (SCZ), DD, bipolar disorders (BD), ANX, eating disorders (ED), autism spectrum disorders (ASD), ADHD, idiopathic developmental intellectual disability (IDID). In UKB, the diagnosis of dementia and mental disorders was based on the International Statistical Classification of Diseases and Related Health Problems, Tenth Revision (ICD-10), consistent with the definitions used in the GBD framework (Supplementary Table 1). In the GBD database, we used ‘Alzheimer’s disease (AD) and other dementias’ to refer to dementia and ‘mental disorders’ to refer to overall mental disorders according to the classification of causes by GBD. Conduct disorder was excluded due to missing data in population aged 40 years and older. We excluded ASD, ADHD, and IDID from the co-occurrence and individual analysis in UKB, as these neurodevelopmental conditions typically manifest in childhood or adolescence and have negligible diagnostic cases in populations aged 40 years and older. Details were summarized in Supplementary Table 1.[17]. Additionally, we utilized the socio-demographic index (SDI) measured in 2021 and categorized 204 countries and territories in different development levels.

Dementia was further categorized as early-onset of dementia (EOD) and late-onset of dementia (LOD). EOD referred to onset of dementia between 40 and 64 years old, and LOD referred to those with dementia aged 65 and above.[18] Eight mental disorders were further categorized. Neurodevelopmental disorders including ASD, ADHD, and IDID. SCZ was classified as ‘schizophrenia or other primary psychotic disorders’. DD, BD were classified as ‘mood disorders’. ANX were classified as ‘anxiety or fear-related disorders’. ED were categorized as ‘feeding and eating disorders’.

### 2.3 Definition of concurrence of dementia and mental disorders

At the individual-level, concurrence was defined as participants having diagnoses of both dementia and a mental disorder. Based on the recorded diagnostic dates, we classified the temporal relationship between dementia and each mental disorder into three categories: mental disorder preceding dementia, dementia preceding mental disorder, or concurrent (diagnosed on the same date).

At the country level, we estimated co-existence and co-occurrence patterns using the following steps. Age-standardized rates (per 100,000 population) based on the world standard population were calculated among adults aged 40 and above.[19] Age-standardized rate was calculated using the following equation (1), where *a_i_* is the age specific rate and *w_i_* is the weight in the same age subgroup of the chosen references standard population (in which i denotes the i^th^ age class) and A is the upper age limit.[19]

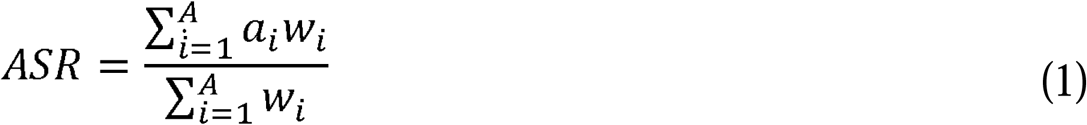

Three steps were followed to categorize the concurrence and co-occurrence patterns of dementia and mental disorders:

Firstly, we categorized the age-standardized prevalence rate (ASPR) and age-standardized incidence rate (ASIR) of both dementia and mental disorders into three classification levels based on tertiles: low (<1^st^ tertile), middle (1^st^-2^nd^ tertile), high (2^nd^ tertile), which served as thresholds for categorization.

Secondly, if the prevalence or incidence rates level of dementia and mental disorders were both at Low, Middle, or High level, the country or territory was classified as Low, Middle, or High-co-existence/co-occurrence group, respectively. If the prevalence or incidence rates level of dementia and mental disorders were not identical, the country/territory was classified as dominated by the disorder with a higher burden. For example, if the mental disorders burden was High whilst dementia burden was Middle, it was classified as mental disorders-dominant group.

Finally, 204 countries and territories were divided into five concurrent (co-existence/co-occurrence) patterns (high co-existence/co-occurrence, middle co-existence/co-occurrence, low co-existence/co-occurrence, dementia-dominant, and mental disorders-dominant) based on the relative level of dementia and mental disorders referring to previous established method.[20]

### 2.4 Risk factors profile

Age, sex, and 11 modifiable risk factors of dementia were included.[3] The modifiable factors were based on previously established criteria[21] and covered early life (less education), midlife (hearing loss, physical inactivity [low, moderate, high], diabetes, smoking [ever, previous, current], hypertension, hyperlipidaemia, obesity, alcohol consumption [ever, previous, current], and traumatic brain injury [TBI]), and late-life (visual loss). Detailed definitions were shown in Supplementary Table 2.[3]

### 2.5 Statistical analysis

First, the prevalence and diagnostic sequence of dementia-mental disorder concurrence were summarized. Age- and sex-adjusted Cox regression was performed to examine the association between mental disorders and incident dementia. We then estimated the associations of multiple risk factors with incident dementia and incident mental disorders.

From the GBD database, to measure the temporal trend of dementia and mental disorders from 1990 to 2021, average annual percentage changes (AAPCs) were estimated using joinpoint regression. The joinpoint model assumes that its regression mean function is piecewise linear, with segments are continuously connected at unknown change-points, and is easier to interpret trend changes than non-linear models.[22] If AAPC value and 95% uncertainty interval (UI) were both >0 (or both <0), the corresponding rate was considered to be in an upward (or downward) trend. AAPC was calculated using the following equation (2), where *b_i_* is the slope coefficient for the i^th^ segment with i indexing the segments in the desired range of years, and *w_i_* is the length of each segment in the range of years.[19]

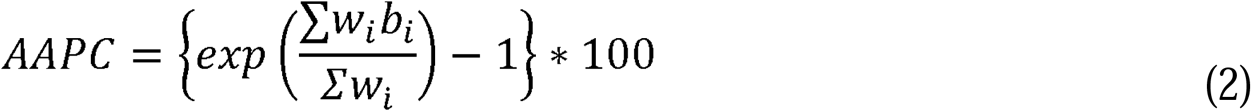

Then, we characterised the co-existence and co-occurrence burden of dementia with mental disorders from 1990 to 2021 on a global scale. Chi-square tests were used to compare the differences in co-existence and co-occurrence patterns among 204 countries and territories between 1990 and 2021.

We established a continuous-time multistate Markov model to estimate the pattern of transitions over the 31 years from 1990 to 2021. The state set contains five states: 1: high co-existence/occurrence, 2: middle co-existence/occurrence, 3: low co-existence/occurrence, 4: dementia-dominant, 5: mental disorders-dominant. Given a country or territory, we let *S_n_* denote its state in year *n*. The transition probability matrix (after time t) is a 5 × 5 matrix *P*(*t*) = (*P*(*t*)*_ij_*) whose entry in the *i*th row, *j*th column is *P*(*S_n_*_+*t*_ = *j*|S*_n_* = *i*). The transition density matrix 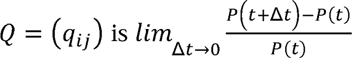 and we have

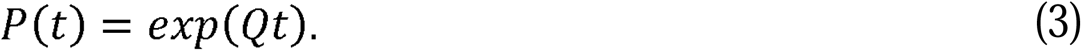

We used maximum likelihood estimation (MLE) to fit our model to the data. Since we had data over a series of years (1990, 1991, 1992,……, 2021), we considered all the transitions after one year. Given a county or territory, if S*_n_* = *i* and S*_n+_*_1_ = *j*, this would contribute to likelihood function

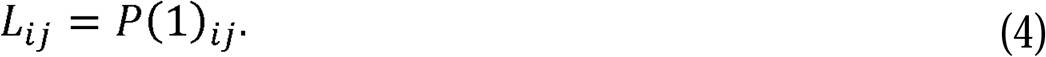

By equation (3) and (4), we have *L_ij_* = *exp*(*Q*)*_ij_*. The full likelihood function L(Q) is the product of all such terms *L_ij_* over all the transitions after one year and all the countries or territories. We took the logarithm of L(Q) and apply quasi-Newton method to optimize *log*(*L*(*Q*)). The matrix *Q* is determined by maximizer of *log*(*L*(*Q*)).

The average time spent in state *j* starting from state *i* between time *a* and time *b* is estimated by

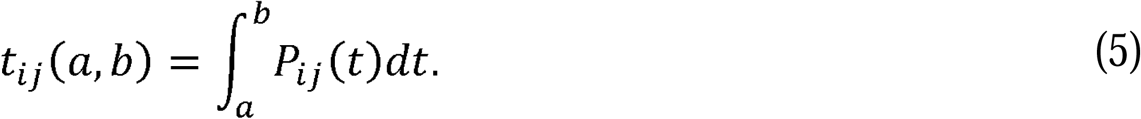

The total expected time spent in state *j* in the past 31 years 1990-2021 is estimated by

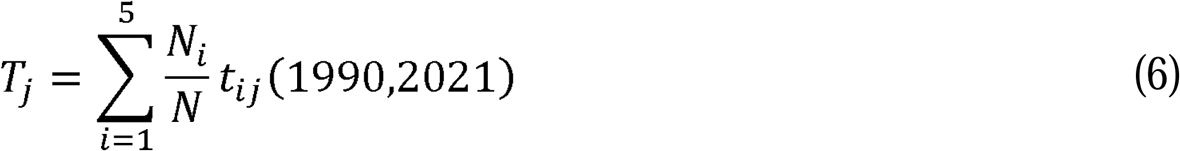

where *N_i_* is the number of countries or territories observed in state *i* in year 1990 and *N* is the total number of countries or territories of the data.

Then, to explore the regional distribution of co-existence and co-occurrence patterns, we summarized the number and proportion of countries among 21 territories. To further explain the results, we also summarized the proportion of pattern-specific countries with different SDI levels.

We further investigated the age (EOD vs LOD) and sex (male vs female) difference of concurrent burden of dementia with mental disorders. All statistical analyses were conducted using R (version 4.4.1) and Joinpoint (version 5.4.0).

## 3 Results

### 3.1 Individual-level concurrence of dementia with mental disorders

In UKB, 6649 (1.3%) dementia cases were identified among 502461 participants. Among those with specific mental disorders, the prevalence of concurrent dementia was higher in participants with BD (7.3%), followed by SCZ (7.1%), ED (5.7%), DD (5,0%), and ANX (4.2%) (Figure 1.A). Among cases, the proportion of concurrent diagnoses ranged from 9.7% to 36.4%, with most disorders (51.5%–71.5%) showing a predominance of mental disorder preceding dementia (Figure 1.B). Mental disorders, such as ANX, DD, SCZ, and BD, were associated with an increased risk of incident dementia (Figure 1.C). Specifically, overlapping risk factors between dementia and each mental disorder were as follows: diabetes and lower educational levels (schizophrenia); smoking, drinking, and obesity (eating disorders); and smoking, drinking, hypertension, and hyperlipidaemia, and obesity (depressive and anxiety disorders). Diabetes was also associated with a higher risk of incident depressive disorders (Figure 1.D).

**Figure 1.**
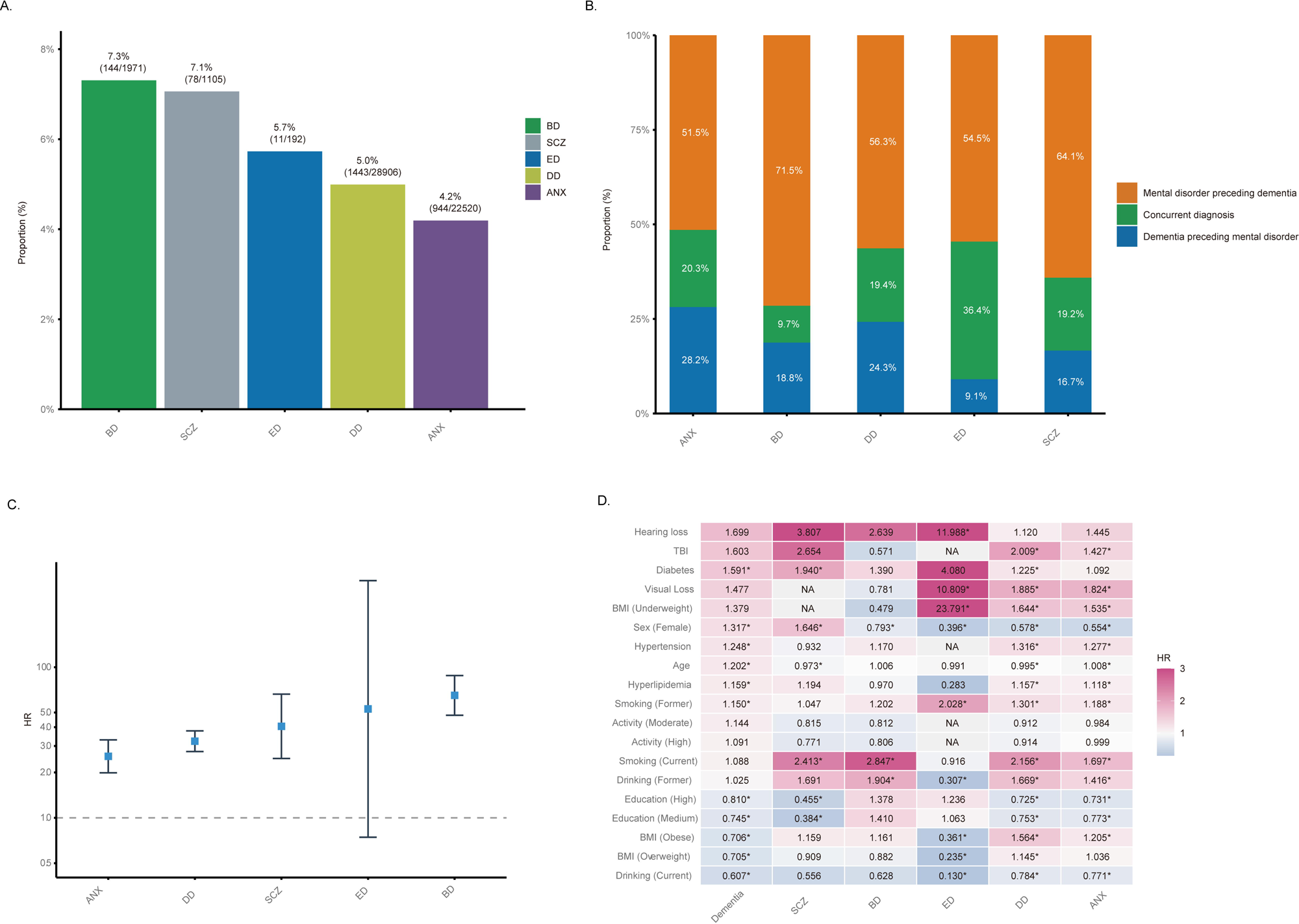
Individual-level concurrence of dementia with mental disorders. Note: A. Prevalence of comorbid dementia across mental disorders; B. Diagnostic precedence between dementia and mental disorders in comorbid cases; C. Associations between mental disorders and incident dementia; D. Risk factor profiles for dementia and mental disorders. Models in C were controlled for age and sex; SCZ: Schizophrenia; DD: Depressive Disorders; BD: Bipolar Disorders; ANX: Anxiety Disorders; ED: Eating Disorders.

### 3.2 Country-level concurrent burden of dementia with mental disorders

Globally, the prevalence (AAPC: 0.09; 95%UI: 0.06, 0.11) and incidence (AAPC: 0.06; 95%UI: 0.04, 0.08) of dementia slightly increased between 1990 and 2021, mainly in East Asia and high-income Asia Pacific regions. However, in the majority of regions (19 out of 21) the prevalence and incidence of dementia decreased over the past 31 years (Supplementary Table 3-4, Figure 2. A and D). On the contrary, the global and regional mental disorders burden, except for BD, ADHD, and IDID, including the prevalence (AAPC: 0.19; 95%UI: 0.16, 0.22) and incidence (AAPC: 0.38; 95%UI: 0.31, 0.47) significantly increased over the past 31 years (Supplementary Table 5-6, Figure 2. B and E).

**Figure 2.**
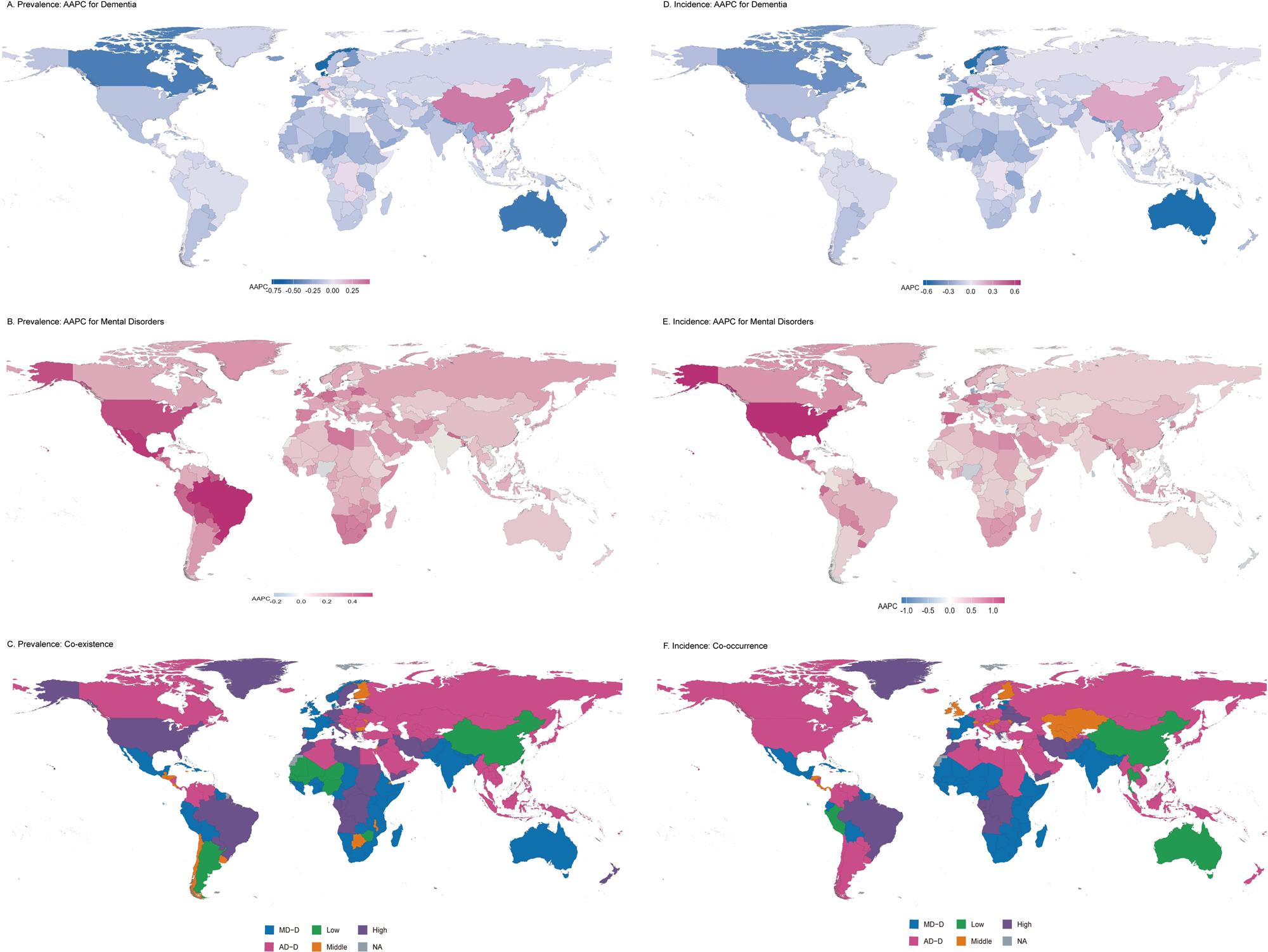
Global temporal trends in ASPR and ASIR and co-existence and co-occurrence patterns. Note: A. AAPC of ASPR for dementia, B. AAPC of ASPR for mental disorders; C. Co-existence; D. AAPC of ASIR for dementia, E. AAPC of ASIR for mental disorders; F. Co-occurrence. AAPC value were expressed using colors, with red for >0 and blue for <0. The darker color represented stronger magnitude. Low, Middle, or High co-existence/co-occurrence mean that the prevalence/incidence rates level of both dementia and mental disorders were at Low, Middle, or High levels, respectively, in a country or territory. Dementia-Dominant means that the dementia rate level was higher than that of mental disorders. Mental Disorders-Dominant means that the mental disorders rate level was higher than that of dementia. AAPC: Average Annual Percentage Change; ASPR: Age-Standardized Prevalence Rate; ASIR: Age-Standardized Incidence Rate; AD-D: Dementia-Dominant; MD-D: Mental Disorders-Dominant; Low: Low Co-existence/Occurrence; Middle: Middle Co-existence/Occurrence; High: High Co-existence/Occurrence; NA: Not Available.

We observed a common co-existence and co-occurrence burden between dementia and mental disorders (Supplementary Table 7, Figure 3. C and F). Dementia exhibited varying co-existence patterns with diverse mental disorder types (Figure 3). In 2021, the leading mental disorders with a high co-existence pattern with dementia were IDID (36 countries, 17.6% in 204 countries and territories), ADHD (32, 15.7%), ED (30, 14.7%), and ANX (29, 14.2%) (Figure 3. C).

**Figure 3.**
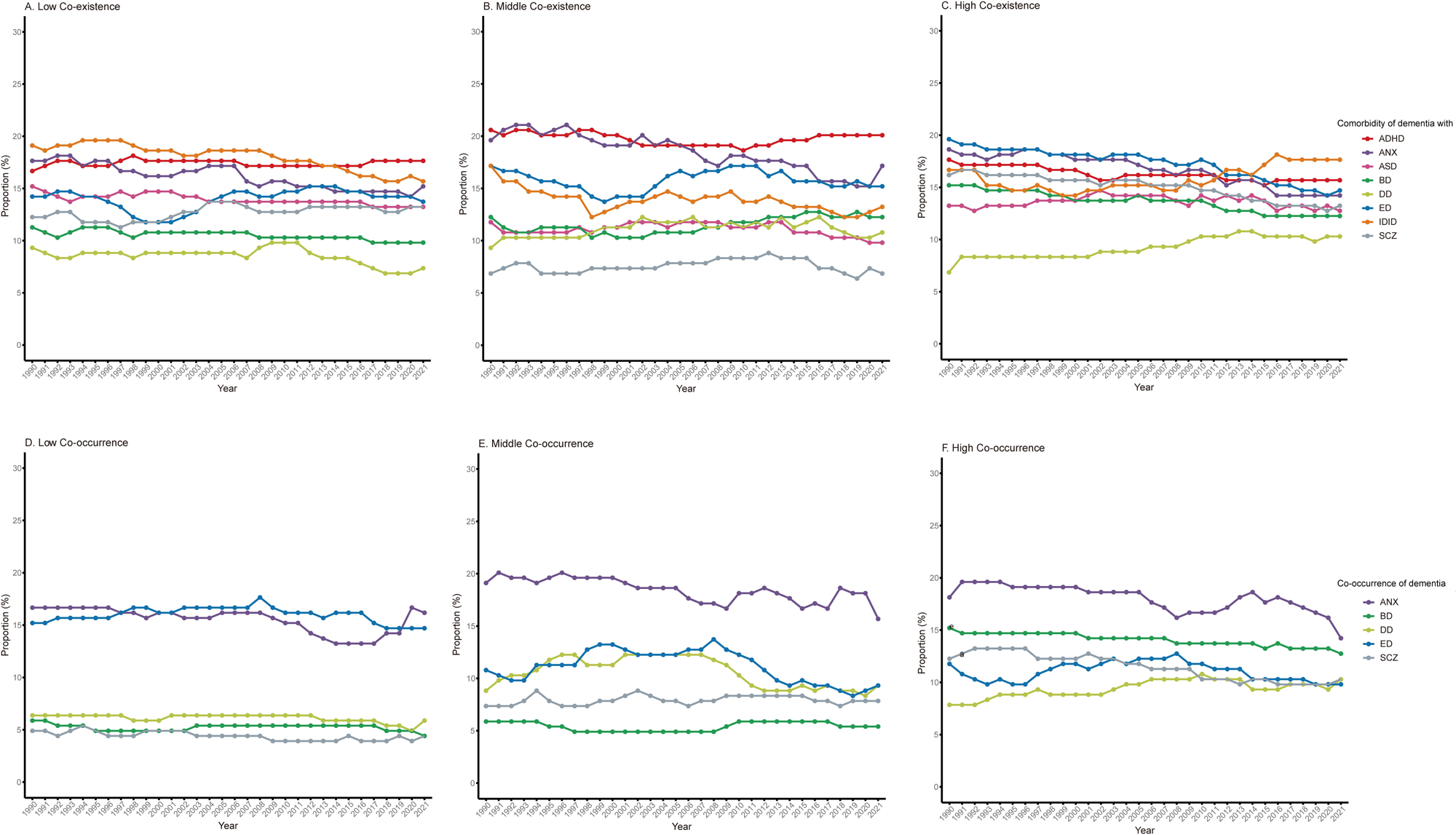
Proportions of co-existence and co-occurrence of dementia with eight mental disorders, 1990-2021. Note: Low, Middle, or High co-existence/co-occurrence mean that the prevalence/incidence rates level of both dementia and mental disorders were at Low, Middle, or High levels, respectively, in a country or territory. SCZ: Schizophrenia; DD: Depressive Disorders; BD: Bipolar Disorders; ANX: Anxiety Disorders; ED: Eating Disorders; ASD: Autism Spectrum Disorders; ADHD: Attention-Deficit/Hyperactivity Disorder; IDID: Idiopathic Developmental Intellectual Disability.

Similarly, the leading mental disorders with a high co-occurrence pattern with dementia were ANX (29, 14.2%) (Figure 3. F).

### 3.3 Long-term concurrent burden over the past 31 years

From 1990 to 2021, although the disease patterns fluctuated (Chi-square Ps<0.01), there was a high proportion of countries and territories maintaining concurrent patterns (Supplementary Table 8).

For 35 countries with high co-existence of dementia with overall mental disorders observed in 1990, 26 (74.3%) countries had a high co-existence pattern in 2021, and the expected total duration of remaining this pattern was 25.0 (80.6%) years over a period of 31 years (Supplementary Table 7).

The results showed a relatively high co-existence burden between dementia and mental disorders, especially neurodevelopmental disorders (e.g., IDID, ADHD), ANX, and ED (Figure 4). Over the past 31 years, the global total expected time in high co-existence of dementia with ED, IDID, ADHD, and ANX was 5.27 (17.0%), 5.16 (16.6%), 5.14 (16.6%), and 5.06 (16.3%) years, respectively, compared to 4.93 (15.9%) years for overall mental disorders (Figure 4. C). We also observed a common dementia-ANX co-occurrence pattern. The global total expected time in high co-occurrence between dementia and ANX was 5.13 (16.5%) years, compared to 2.99 (9.65%) years for overall mental disorders (Figure 4. F).

**Figure 4.**
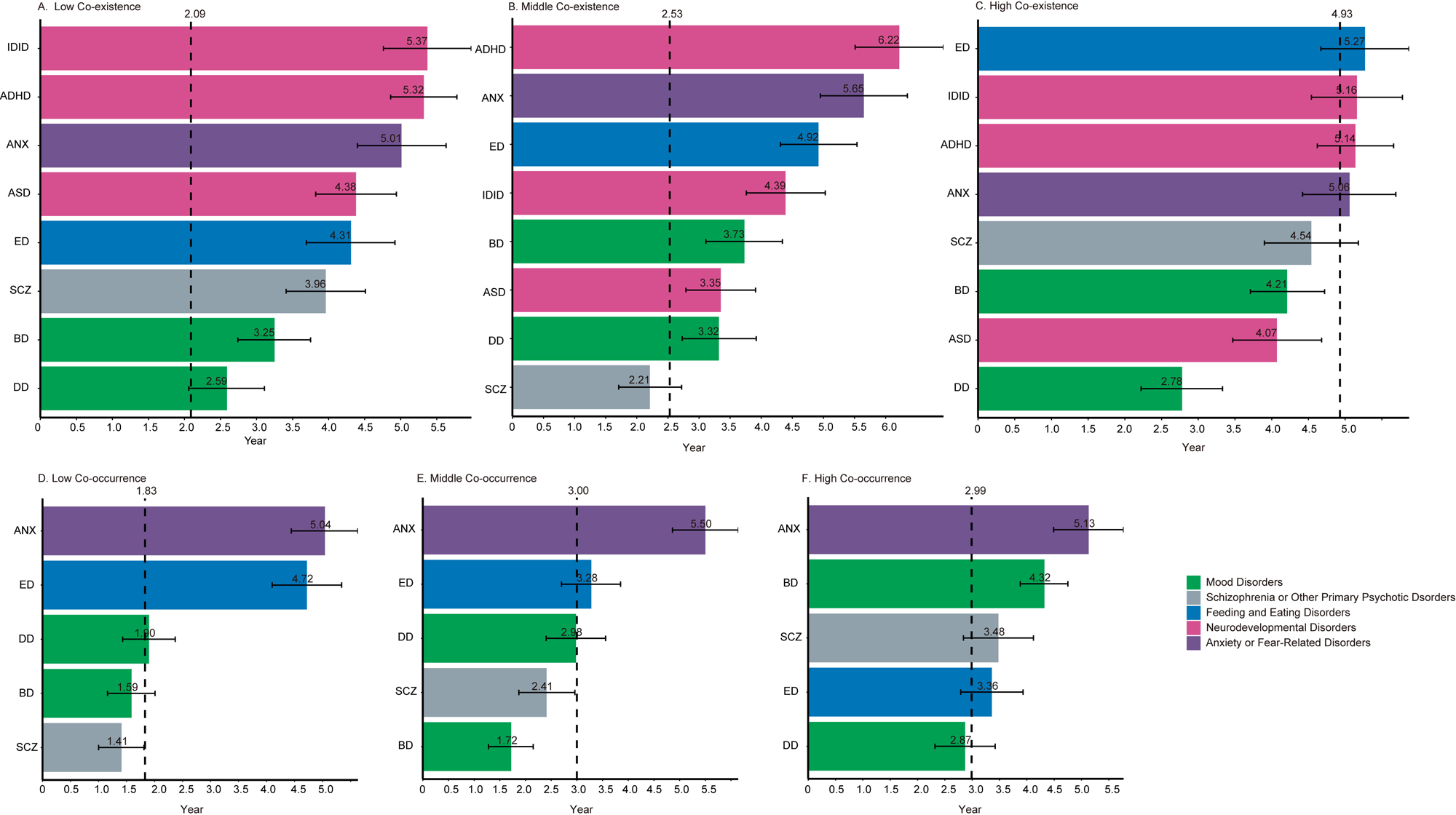
Total expected time in co-existence and co-occurrence of dementia with eight mental disorders, 1990–2021. Note: The vertical reference line is the total expected concurrent time of dementia with overall mental disorders. Low, Middle, or High co-existence/co-occurrence mean that the prevalence/incidence rate levels of both dementia and mental disorders were at Low, Middle, or High levels, respectively, in a country or territory. SCZ: Schizophrenia; DD: Depressive Disorders; BD: Bipolar Disorders; ANX: Anxiety Disorders; ED: Eating Disorders; ASD: Autism Spectrum Disorders; ADHD: Attention-Deficit/Hyperactivity Disorder; IDID: Idiopathic Developmental Intellectual Disability.

### 3.4 Spatial distribution of concurrent countries

Figure 5 shows the regional distribution of countries with consistent patterns of co-existence and occurrences in both 1990 and 2021.

**Figure 5.**
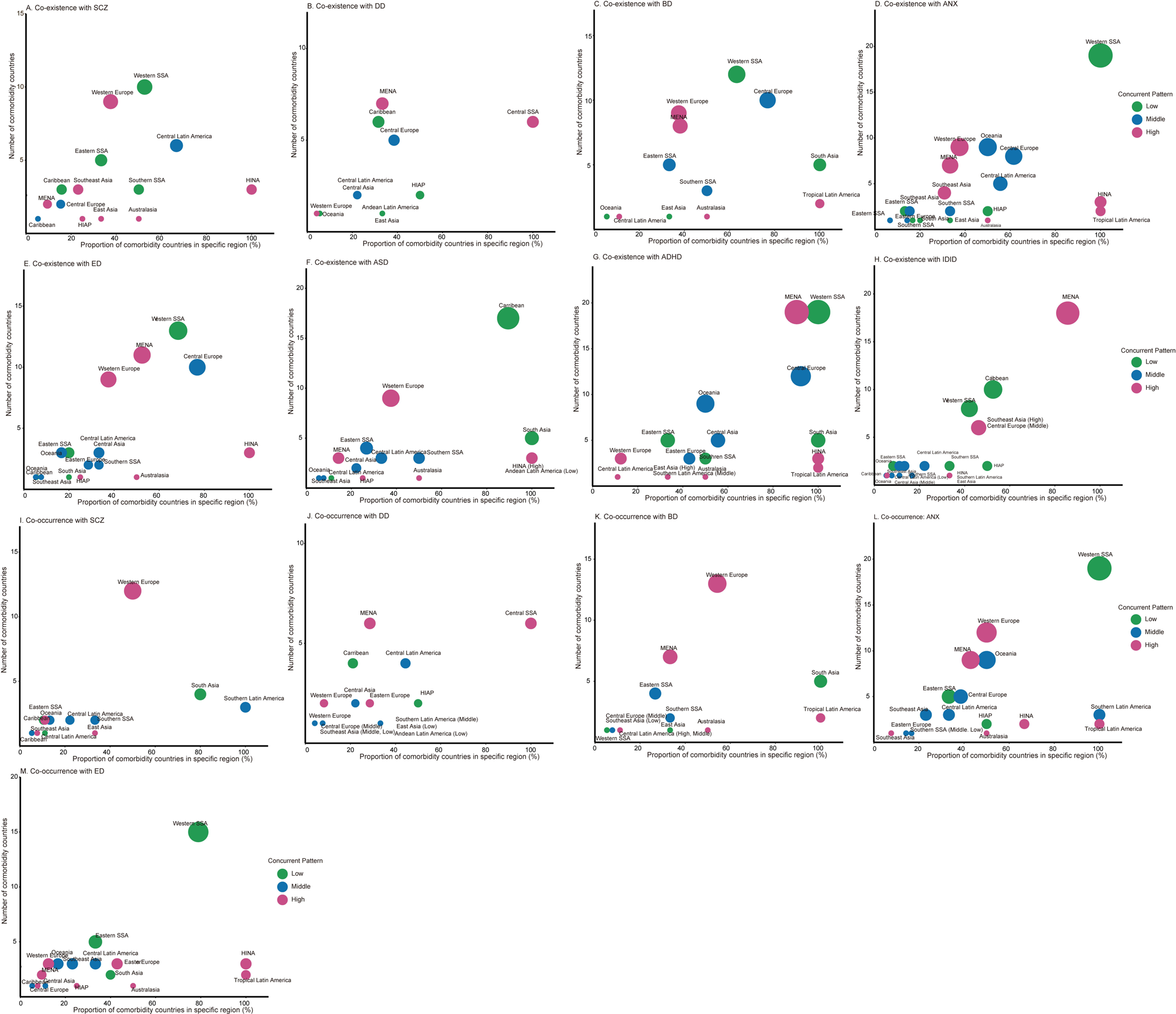
Number and regional proportion of countries with co-existence/co-occurrence in both 1990 and 2021. Note: The countries with concurrent (co-existence or co-occurrence) of dementia with eight mental disorders in both 1990 and 2021 were selected. Proportions of concurrent countries in specific region = Number of concurrent countries/Total number of countries in that region. Supplementary Table 9 showed the total number of countries in 21 regions. Low, Middle, or High co-existence/co-occurrence mean that the prevalence/incidence rates level of both dementia and mental disorders were at Low, Middle, or High levels, respectively, in a country or territory. SCZ: Schizophrenia; DD: Depressive Disorders; BD: Bipolar Disorders; ANX: Anxiety Disorders; ED: Eating Disorders; ASD: Autism Spectrum Disorders; ADHD: Attention-Deficit/Hyperactivity Disorder; IDID: Idiopathic Developmental Intellectual Disability; Low: Low Co-existence/Occurrence; Middle: Middle Co-existence/Occurrence; High: High Co-existence/Occurrence; SSA: Sub-Saharan Africa; MENA: Middle East and North Africa; HINA: High-income North America; HIAP: High-income Asia Pacific.

In both 1990 and 2021, most countries with high dementia-SCZ (9 countries, 45% in total, 37.5% in this region, Figure 5. A), dementia-BD (9, 42.9%, 37.5%, Figure 5. C), dementia-ANX (9, 34.6%, 37.5%, Figure 5. D), dementia-ED (9, 36.0%, 37.5%, Figure 5. E), and dementia-ASD (9, 52.9%, 37.5%, Figure 5. F) co-existence was located in Western Europe. All high-income North America countries (3 countries) had high dementia-SCZ, dementia-ANX, dementia-ED, dementia-ASD, dementia-ADHD co-existence in both 1990 and 2021 (Supplementary Table 9).

In 21 Middle East and North Africa, many countries showed high dementia-neurodevelopmental disorders co-existence, such as ADHD (19, 63.3%, 90.5%, Figure 5. G) and IDID (18, 69.2%, 85.7%, Figure 5. H).

In addition, all (2 countries) tropical Latin America countries evidenced high dementia-BD, dementia-ANX, dementia-ADHD co-existence. A total of 6 (23.1%, 46.2%, Figure 5. H) Southeast Asian countries had high dementia-IDID co-existence. Only 16 countries had high dementia-DD patterns, and 7 (50.0%, 33.3%, Figure 5. B) were predominantly located in Middle East and North Africa. All central sub-Saharan Africa countries had high co-existence and co-occurrence of dementia-DD burden.

Dementia had a low co-existence with SCZ, BP, ANX, ED, ADHD, IDID in western Sub-Saharan Africa. South Asia was covered by low co-existence of dementia-BD, dementia-ASD, and dementia-ADHD. Caribbean was covered by low dementia-DD, dementia-ASD, dementia-IDID co-existence. Andean Latin America was covered by low dementia-ASD co-existence. (Figure 5. A-H)

Similar to the co-existence patterns, a consistent regional distribution was found in the co-occurrence of dementia and mental disorders. (Figure 5. I-M)

For specific countries, we observed that New Zealand in Australasia, Western European countries (e.g., Iceland, Greece, Netherlands, Austria, Germany, Italy, Belgium, Cyprus, and Sweden), Middle East and North Africa countries (e.g., Iran, Kuwait, Lebanon, Palestine, Tunisia), high-income North America (e.g., Canada, United States, and Greenland) had a high co-existence of dementia with mental disorders.

### 3.5 Exploratory analyses

Among 40 countries with high SDI, a higher proportion of countries with high co-existence and occurrence of dementia with ED (co-existence: 42.5%, co-occurrence: 30.0%), ASD (42.5%), SCZ (40.0%, 25.0%), ADHD (27.5%), ANX (27.5%, 30.0%), and BD (25.0%, 30.0%) was observed (Supplementary Table 10, Figure S1. C).

Notably, similar results of co-existence and co-occurrence burden were found between EOD-mental disorders and LOD-mental disorders, as well as in both males and females (Supplementary Table 11-12). Over the past 31 years, neurodevelopmental disorders, such as IDID and ADHD, showed a high co-existence burden with both EOD and LOD.

## 4 Discussion

The study found a substantial concurrence of dementia and mental disorders at the individual level and further quantified temporal trends and spatial distribution of global concurrent burden between dementia and eight mental disorders at the country level, providing a comprehensive and nuanced understanding of dementia-mental disorders’ profiles. Individually, mental disorders predominantly preceded or coincided with dementia in concurrent cases, with evidence of causal links and shared risk factors between the two. Globally, we observed a common co-existence and co-occurrence burden of dementia with mental disorders, despite a decreasing prevalence and incidence of dementia over the course of three decades. There was a high co-existence and co-occurrence of dementia with ANX, ED, and SCZ in high SDI areas such as Western Europe and High-income North America regions. The Middle East and North Africa region had high co-existence of dementia with neurodevelopmental disorders, such as IDID and ADHD. Neurodevelopmental disorders have been highly concurrent with both EOD and LOD.

At the individual level, we found that participants with certain mental disorders had a higher prevalence of concurrent dementia, which may be partly explained by the associations between mental disorders and incident dementia. Our results were similar with previous studies linking mental disorders to dementia.[6] We further found that dementia shares lifestyle risk factors such as smoking and drinking with ED, DD, and ANX, and that metabolic diseases such as diabetes, hypertension, and hyperlipidemia may serve as additional shared risk factors between dementia and mental disorders, particularly SCZ, DD, and ANX.

Collectively, the shared risk factors—consistent with prior evidence—may underpin the concurrence between dementia and mental disorders at both individual and population levels.[3,7,23]

At the country level, over the past 31 years, global countries and territories on average experienced 5.27, 5.16, 5.14, and 5.06 years of high dementia co-existence with ED, IDID, ADHD, and ANX, respectively. Our results aligned with previous epidemiology studies that demonstrated eating disability,[24] neurodevelopmental disorders,[25] and ANX were commonly concurrent with dementia. Although there is a lack of data pertaining to the incidence of neurodevelopmental disorders, we observed the high co-existence of them with dementia in the population aged 40 and older. This is consistent with previous studies suggesting neurodevelopmental disorders shared genetic risk of developing dementia and usually persist from childhood into adulthood.[25] Also, epidemiological studies showed that participants including both children and adults with neurodevelopmental disorders, such as ADHD[26] had a higher risk of developing dementia.

The findings of this study reveal a globally significant regional specificity in the concurrent burden of dementia and mental disorders, with distinct countries and regions demonstrating unique, dominant patterns of multimorbidity. In high SDI regions, exemplified by Western Europe and High-income North America, numerous epidemiological studies have corroborated a declining trend in the incidence of dementia in recent years.[10,11] This favorable trajectory is largely attributed to advancements in several domains, including higher educational attainment, effective management of cardiovascular diseases and their associated risk factors, and the development of pharmacological interventions.[27] These promising trend of results provided further evidence in improving public health measures for dementia. We propose for middle-aged and young adult populations, countries should integrate modifiable risk factors including cardiocerebrovascular health, psychological stress, and lifestyle into routine health management protocols. In this process, decentralizing certain mental health services to general practitioners and non-physician healthcare providers could enhance the engagement and effectiveness of primary healthcare systems in health management.[28] In older age, large-scale screening for cognitive impairment and mental disorders co-existence should be conducted simultaneously, especially in regions with a high burden of cooccurring dementia and mental disorders (i.e. Western Europe and high-income North American countries). Personalized intervention protocols should be developed based on risk levels, encompassing cognitive training, lifestyle guidance, vascular risk factor control, and pharmacological interventions where necessary.

Additionally, our findings reveal a distinct burden pattern dominated by the concurrence of dementia with neurodevelopmental disorders in the Middle East and North Africa region, particularly IDID and ADHD. This observation aligns with and extends prior regional studies on ADHD, collectively highlighting the unique nature of this co-existence profile within this specific geographical context.[29] It is likely attributable to a confluence of systemic factors, including limited insurance coverage, lack of services, insufficient amenities, and the stigma associated with mental health issues.[30]. These findings underscore that the precision prevention and control of dementia and mental disorders necessitate a paradigm shift from reactive disease treatment models to proactive, integrated, and life-course health management strategies. Early screening and intervention for neurodevelopmental disorders should be a priority in the early stages of life.

The present study has several strengths. Firstly, we comprehensively estimated the individual and global burden of co-existence and co-occurrence of dementia and mental disorders from temporal and spatial perspective. Secondly, we estimated the co-existence and occurrence burden of dementia with eight mental disorders subtypes, covering early-onset and later-onset ages, sex, and regions, which offers a broader scope than prior single-disease and region-specific studies.[2,12] Thirdly, multi-state Markov models were used to quantify long-term global burden over 31 years, which obtained highly comparable results and provided novel evidence of dementia-mental disorders co-existence and co-occurrence. Several limitations should be acknowledged. Firstly, although we estimated potential associations between mental disorders and dementia and identified shared risk factors that may partly explain their concurrence, the underlying mechanisms are highly complex. These mechanisms may differ between individual- and country-level determinants, which warrants further investigation. Secondly, diagnostic variability across time and regional discrepancies in dementia and mental disorders may influence estimates, although harmonization approaches were applied to improve comparability across different diagnostic definitions.

Our study highlighted a substantial concurrence burden of dementia with mental disorders at both individual and global perspectives. To address the growing public health challenge of dementia and mental disorders, our findings highlight the importance of developing more targeted prevention and control strategies that specifically address the concurrence burden of these diseases.

## Declarations

### Ethics approval and consent to participate

The UKB study was approved by the North West Multi-Centre Research Ethics Committee as a Research Tissue Bank. All participants provided written informed consent.

### Consent for publication

Not applicable.

### Availability of data and materials

The data used during the current study are available from the corresponding author on reasonable request. The data from UK Biobank and Global Burden of Disease are openly available at [https://www.ukbiobank.ac.uk/] and [https://ghdx.healthdata.org/gbd-results-tool]. All code used to analyse the data and generate results will be uploaded to GitHub and made publicly available upon the publication of the article.

### Authors’ contributions

XX is the joint senior author. XX and HRZ conceived and designed the study. XX supervised the study. HRZ, XWL, and ZPX performed the statistical analysis. All authors contributed to the acquisition, analysis, or interpretation of data. HRZ, TP, YQL, ZPX, and XX visualized the results and drafted the manuscript. All authors read and approved the final manuscript. XX is the guarantor and attests that all listed authors meet authorship criteria and that no others meeting the criteria have been omitted.

## Acknowledgements

Not applicable.

## Additional files

Supplementary Material.docx.

## Abbreviations

AAPCs: average annual percentage changes
AD: Alzheimer’s Diseases
ADHD: attention-deficit/hyperactivity disorder
ANX: anxiety disorders
ASD: autism spectrum disorders
ASIR: age-standardized incidence rate
ASPR: age-standardized prevalence rate
BD: bipolar disorders
DD: depressive disorders
DSM: Diagnostic and Statistical Manual of Mental Disorders
ED: eating disorders
EOD: early-onset of dementia
GBD: Global Burden of Disease
ICD: International Classification of Diseases
IDID: idiopathic developmental intellectual disability
IQ: Intelligence Quotient
LOD: late-onset of dementia
MLE: maximum likelihood estimation
SDI: socio-demographic index
UI: uncertainty interval
UKB: UK Biobank
SCZ: schizophrenia

## Footnotes

① In this study, ‘concurrence’ was used as an umbrella term encompassing concurrence at the individual-level and prevalence-based ‘co-existence burden’ and incidence-based ‘co-occurrence burden’ at the country-level between dementia and mental disorders.

