## Supplemental File 1 for "Concurrence of Dementia and Mental Disorders Across Populational Epidemiology and Global Disease Burden Perspectives"

Supplementary Table 1: Definitions of Alzheimer’s disease and other dementias and mental disorders

Supplementary Table 2: Codes used in the UKB study to identify risk factors

Supplementary Table 3: AAPC of ASR for dementia and mental disorders among 204 countries

Supplementary Table 4: Global and regional AAPC of ASR for dementia

Supplementary Table 5: Global and regional AAPC of ASR for overall mental disorders.

Supplementary Table 6: Global and regional AAPC of ASR for eight mental disorders subtypes.

Supplementary Table 7: Co-existence patterns of dementia with overall mental disorders among 204 Countries from 1990 to 2021

Supplementary Table 8: The number of countries with co-existence and cooccurrence patterns in 1990 and 2021

Supplementary Table 9: The total number of countries in 21 regions.

Supplementary Table 10: SDI levels of 204 countries.

Supplementary Table 11: Total expected time spent in low, middle, and high co-existence and co-occurrence patterns of early-onset and later-onset of dementia with eight mental disorders over the past 31 years

Supplementary Table 12: Total expected time spent in low, middle, and high co-existence and co-occurrence patterns of dementia with eight mental disorders stratified by sex over the past 31 years

Supplementary Figure 1: Proportion of countries with co-existence and co-occurrence across five SDI levels.

Supplementary Table 1: Definitions of Alzheimer’s disease and other dementias and mental disorders

| **Cause** | **Definition** | **ICD-10** | **DSM-IV-TR** |
| --- | --- | --- | --- |
| Alzheimer’s disease and other dementias | Multiple cognitive deficits manifested by both memory impairment and one of the following: aphasia, apraxia, agnosia, disturbance in executive functioning; Must cause significant impairment in occupational functioning and represent a significant decline; Course is characterised by gradual onset and continuing cognitive decline; Cognitive deficits are not due to other psychiatric conditions; Deficits do not occur exclusively during the course of a delirium. | F00-F02.0, F02.8-F03.91, F06.2, G30-G31.1, G31.8-G32.89 | 290-290.9, 294.0-294.9, 331-331.2, 331.6-331.7,  331.82,  331.89-331.9 |
| Schizophrenia | Involves the experience of positive symptoms (e.g., delusions, hallucinations, thought disorder) and negative symptoms (e.g., flat affect, loss of interest, and emotional withdrawal). | F20 | 295.10-295.30, 295.60, 295.90 |
| Depressive disorders | Including major depressive disorder, defined as the presence of at least one depressive episode characterized by depressed mood or loss of interest/pleasure for most of every day, for at least two weeks, and dysthymia, defined as chronically depressed mood occurring for most of the day, more days than not, for at least two years (or at least one year in children and adolescents). | F32.0-9,  F33.0-9,  F34.1 | 296.21–24, 296.31–34,  300.4 |
| Bipolar disorders | Bipolar I is characterized by at least one manic episode, which can also alternate with a major depressive episode. Bipolar II is characterized by hypomanic episodes alternating with major depressive episodes. Cyclothymia is characterized by subsyndromal hypomanic and major depressive episode. Bipolar disorder not otherwise specified is characterized by clinically significant symptoms of bipolar disorder which do not meet criteria for the other diagnoses. We estimated burden for the entire spectrum of bipolar disorder simultaneously, rather than individually for each subtype of the disorder. At a minimum, epidemiological studies needed to report on bipolar I and bipolar II. | F30.0-F30.9, F31.0–F31.6, F31.8–F31.9, F34.0 | 296.0–296.7, 296.89, 301.13 |
| Anxiety disorders | Involves experiences of intense fear and distress, typically in combination with other physiological symptoms. Anxiety disorders were modelled as a single cause for “any” anxiety disorder to avoid the double-counting of individuals meeting criteria for more than one anxiety disorder. Epidemiological estimates reporting an outcome for “any” or “total” anxiety disorders were included if they reported on at least three anxiety disorders. | F40-42, F43.0, F43.1, F93.0-93.2, F93.8. | 300.0-300.3, 208.3, 309.21, 309.81 |
| Eating disorders | Including anorexia nervosa, characterized by refusal to maintain body weight at or above a minimally normal weight for age and height, intense fear of gaining weight, and disturbance in the way in which one’s body weight or shape is experienced, and bulimia nervosa, characterized by recurrent episodes of binge eating and inappropriate compensatory behavior to prevent weight gain. These episodes must occur, on average, at least twice a week for three months. | F50.0-50.1,  F50.2 | 307.1,  307.51 |
| Autism spectrum disorders | Characterized by pervasive impairment in several areas of development, including social interaction and communication skills, along with restricted and repetitive patterns of behaviors and/or interests. Symptoms must be present in the early developmental period, cause clinically significant impairment, and not be better explained by intellectual impairment or global developmental delay. | F84.0, F84.1, F84.2, F84.3, F84.4, F84.5, F84.8, F84.9 | 299.00, 299.80, 299.8, 299.8, 299.10 |
| Attention-deficit/hyperactivity disorder | Characterized by persistent inattention and/or hyperactivity-impulsivity. Diagnosis requires six or more symptoms of inattention or hyperactivity-impulsivity to have persisted for at least six months, in two or more settings; with at least some impairing symptoms being present prior to 7 years of age. | F90 | 314.0, 314.01 |
| Idiopathic developmental intellectual disability | Idiopathic developmental intellectual disability modelled as part of the intellectual disability impairment envelope in GBD 2019. Idiopathic developmental intellectual disability arises from any unknown source after the prevalence of all other sources of Intellectual Disability is accounted for. | In the GBD database, idiopathic intellectual disability was defined to include all levels of severity (based on IQ score): borderline (IQ score 70-85), mild (IQ score 50-69), moderate (IQ score 35-49), severe (IQ score 20-34), and profound (IQ score 0-19). | |

Note: Diagnostic codes are indicated for DSM-IV-TR and ICD-10, but corresponding diagnoses referencing other versions of DSM (DSM-III, DSM-III-R, DSM-IV, DSM-5, and DSM-5-TR) or ICD (ICD-9, ICD-11) were also accepted. DSM-IV-TR: Diagnostic and Statistical Manual of Mental Disorders Fourth Edition, Text Revision; ICD-10: International Classification of Diseases and Related Health Problems 10th Revision; IQ: Intelligence Quotient. In UKB, eating disorders were identified using F50 aligns with the UK Biobank’s standard data categorization.

Supplementary Table 2: Codes used in the UKB study to identify risk factors

| Disease | Algorithmically-derived or self-report | Illness code: ICD-10 |
| --- | --- | --- |
| Hearing loss |  | H90-91 |
| Diabetes | 2443, 6153, 6177 | E10, E100, E101, E102, E103, E104, E105, E106, E107, E108, E109, E11, E110, E111, E112, E113, E114, E115, E116, E117, E118, E119, E12, E121, E123, E125, E128, E129, E13, E130, E131, E132, E133, E134, E135, E136, E137, E138, E139, E140, E141, E142, E143, E144, E145, E146, E147, E148, E149 |
| Hyperlipidaemia | 6153, 6177 | E780, E781, E782, E783, E784, E785, E788, E789 |
| Hypertension | 6153, 6177 | I10 |
| Traumatic brain injury |  | S020, S0200, S0201, S021, S0210, S0211, S027, S0270, S0271, S028, S0280, S0281, S029, S0290, S0291, S06, S060, S0600, S0601, S061, S0610, S0611, S062, S0620, S0621, S063, S0630, S0631, S064, S0640, S0641, S065, S0650, S0651, S066, S0660, S0661, S067, S0670, S0671, S068, S0680, S0681, S069, S0690, S0691, S07, S09, S097, S098, S099 |
| Physical inactivity | 864, 874, 884, 894, 904, 914 |  |
| Visual loss |  | H53-54 |

Note: Abbreviation: ICD-10, the International Statistical Classification of Diseases and Related Health Problems, Tenth Revision; UKB: UK Biobank.

Supplementary Table 3: AAPC of ASR for dementia and mental disorders among 204 countries

| Region | Countries | Dementia | | Mental disorders | |
| --- | --- | --- | --- | --- | --- |
|  |  | Prevalence | Incidence | Prevalence | Incidence |
| East Asia | China | 0.75 (0.65, 0.83) | 0.68 (0.59, 0.75) | 0.07 (0.06, 0.08) | 0.38 (0.31, 0.44) |
| East Asia | Democratic People's Republic of Korea | -0.07 (-0.07, -0.06) | -0.07 (-0.07, -0.07) | 0.02 (0.02, 0.03) | -0.16 (-0.16, -0.15) |
| East Asia | Taiwan (Province of China) | 0.30 (0.28, 0.32) | 0.25 (0.23, 0.27) | 0.12 (0.12, 0.12) | 0.35 (0.33, 0.36) |
| Southeast Asia | Cambodia | -0.12 (-0.13, -0.12) | -0.11 (-0.11, -0.11) | 0.06 (0.03, 0.08) | 0.12 (0.07, 0.14) |
| Southeast Asia | Indonesia | -0.08 (-0.08, -0.08) | -0.07 (-0.08, -0.07) | 0.16 (0.16, 0.17) | 0.45 (0.43, 0.46) |
| Southeast Asia | Lao People's Democratic Republic | -0.09 (-0.09, -0.09) | -0.07 (-0.07, -0.07) | 0.03 (0.00, 0.04) | -0.01 (-0.05, 0.03) |
| Southeast Asia | Malaysia | -0.17 (-0.18, -0.14) | -0.13 (-0.14, -0.12) | 0.17 (0.14, 0.19) | 0.26 (0.22, 0.29) |
| Southeast Asia | Maldives | 0.10 (0.10, 0.10) | 0.10 (0.10, 0.10) | -0.02 (-0.03, -0.01) | -0.33 (-0.34, -0.30) |
| Southeast Asia | Myanmar | -0.26 (-0.27, -0.26) | -0.25 (-0.25, -0.25) | 0.11 (0.10, 0.12) | 0.51 (0.50, 0.52) |
| Southeast Asia | Philippines | -0.14 (-0.14, -0.14) | -0.14 (-0.14, -0.14) | 0.13 (0.10, 0.17) | -0.03 (-0.14, 0.05) |
| Southeast Asia | Sri Lanka | -0.13 (-0.13, -0.12) | -0.11 (-0.12, -0.11) | 0.08 (0.07, 0.09) | -0.23 (-0.25, -0.21) |
| Southeast Asia | Thailand | 0.02 (0.00, 0.03) | 0.03 (0.02, 0.04) | 0.13 (0.12, 0.14) | 0.64 (0.62, 0.65) |
| Southeast Asia | Timor-Leste | -0.22 (-0.22, -0.22) | -0.20 (-0.20, -0.19) | 0.07 (0.06, 0.07) | 0.05 (0.03, 0.07) |
| Southeast Asia | Viet Nam | -0.20 (-0.20, -0.19) | -0.18 (-0.19, -0.18) | -0.03 (-0.06, -0.00) | 0.11 (0.00, 0.17) |
| Oceania | Fiji | -0.07 (-0.07, -0.06) | -0.06 (-0.06, -0.06) | 0.17 (0.15, 0.18) | 0.38 (0.34, 0.40) |
| Oceania | Kiribati | 0.02 (0.01, 0.02) | 0.02 (0.02, 0.02) | 0.11 (0.09, 0.12) | 0.02 (-0.02, 0.05) |
| Oceania | Marshall Islands | -0.12 (-0.13, -0.12) | -0.11 (-0.11, -0.11) | 0.10 (0.08, 0.12) | 0.16 (0.11, 0.19) |
| Oceania | Micronesia (Federated States of) | 0.02 (0.02, 0.02) | 0.02 (0.02, 0.03) | 0.11 (0.08, 0.12) | 0.13 (0.09, 0.15) |
| Oceania | Papua New Guinea | -0.21 (-0.22, -0.21) | -0.20 (-0.20, -0.20) | 0.06 (0.03, 0.07) | 0.02 (-0.04, 0.05) |
| Oceania | Samoa | -0.11 (-0.11, -0.10) | -0.09 (-0.09, -0.09) | 0.06 (0.04, 0.07) | -0.02 (-0.05, -0.00) |
| Oceania | Solomon Islands | -0.01 (-0.01, -0.01) | 0.01 (0.00, 0.01) | 0.12 (0.10, 0.14) | 0.16 (0.11, 0.19) |
| Oceania | Tonga | -0.11 (-0.11, -0.10) | -0.10 (-0.10, -0.09) | 0.10 (0.08, 0.11) | 0.18 (0.14, 0.20) |
| Oceania | Vanuatu | -0.07 (-0.07, -0.07) | -0.06 (-0.07, -0.06) | 0.12 (0.11, 0.13) | 0.13 (0.09, 0.14) |
| Central Asia | Armenia | -0.01 (-0.02, -0.01) | -0.01 (-0.01, -0.01) | 0.36 (0.33, 0.40) | 0.78 (0.71, 0.87) |
| Central Asia | Azerbaijan | -0.10 (-0.10, -0.10) | -0.10 (-0.10, -0.10) | 0.23 (0.21, 0.26) | 0.48 (0.44, 0.53) |
| Central Asia | Georgia | 0.00 (0.00, 0.01) | 0.01 (0.01, 0.01) | 0.26 (0.26, 0.26) | 0.49 (0.48, 0.49) |
| Central Asia | Kazakhstan | -0.08 (-0.08, -0.07) | -0.07 (-0.08, -0.07) | 0.09 (0.08, 0.10) | 0.09 (0.07, 0.12) |
| Central Asia | Kyrgyzstan | 0.03 (0.02, 0.03) | 0.03 (0.03, 0.03) | 0.12 (0.09, 0.17) | 0.07 (0.00, 0.16) |
| Central Asia | Mongolia | 0.04 (0.04, 0.05) | 0.05 (0.04, 0.05) | 0.07 (0.04, 0.08) | 0.19 (0.13, 0.21) |
| Central Asia | Tajikistan | -0.21 (-0.22, -0.21) | -0.21 (-0.21, -0.20) | 0.16 (0.15, 0.17) | 0.13 (0.10, 0.16) |
| Central Asia | Turkmenistan | -0.16 (-0.16, -0.16) | -0.16 (-0.16, -0.16) | 0.10 (0.08, 0.13) | 0.09 (0.05, 0.15) |
| Central Asia | Uzbekistan | -0.01 (-0.02, -0.01) | -0.01 (-0.02, -0.01) | 0.05 (0.03, 0.09) | 0.07 (0.02, 0.14) |
| Central Europe | Albania | 0.01 (0.01, 0.02) | 0.02 (0.02, 0.02) | 0.34 (0.31, 0.39) | 0.82 (0.72, 0.92) |
| Central Europe | Bosnia and Herzegovina | 0.00 (0.00, 0.01) | 0.01 (0.00, 0.01) | 0.09 (0.08, 0.12) | 0.05 (0.02, 0.10) |
| Central Europe | Bulgaria | -0.07 (-0.07, -0.07) | -0.07 (-0.07, -0.07) | 0.29 (0.28, 0.29) | 0.31 (0.30, 0.32) |
| Central Europe | Croatia | -0.11 (-0.12, -0.11) | -0.10 (-0.11, -0.10) | 0.11 (0.08, 0.14) | -0.08 (-0.13, -0.01) |
| Central Europe | Czechia | -0.03 (-0.03, -0.02) | -0.02 (-0.02, -0.02) | 0.09 (0.07, 0.11) | -0.11 (-0.14, -0.06) |
| Central Europe | Hungary | -0.03 (-0.04, -0.03) | -0.03 (-0.03, -0.02) | 0.07 (0.06, 0.07) | -0.14 (-0.16, -0.12) |
| Central Europe | North Macedonia | -0.02 (-0.03, -0.02) | -0.02 (-0.02, -0.02) | 0.30 (0.27, 0.35) | 0.39 (0.33, 0.48) |
| Central Europe | Montenegro | -0.09 (-0.09, -0.08) | -0.08 (-0.09, -0.08) | 0.34 (0.32, 0.37) | 0.60 (0.56, 0.66) |
| Central Europe | Poland | -0.18 (-0.19, -0.18) | -0.17 (-0.18, -0.17) | 0.24 (0.22, 0.26) | 0.62 (0.57, 0.67) |
| Central Europe | Romania | -0.01 (-0.01, -0.00) | -0.01 (-0.01, -0.00) | 0.27 (0.25, 0.31) | 0.50 (0.43, 0.59) |
| Central Europe | Serbia | 0.01 (0.01, 0.01) | 0.01 (0.01, 0.02) | 0.17 (0.16, 0.20) | 0.13 (0.10, 0.19) |
| Central Europe | Slovakia | -0.05 (-0.06, -0.05) | -0.05 (-0.05, -0.05) | 0.18 (0.16, 0.19) | 0.19 (0.16, 0.21) |
| Central Europe | Slovenia | 0.01 (0.01, 0.02) | 0.01 (0.01, 0.02) | -0.00 (-0.04, 0.05) | -0.31 (-0.38, -0.22) |
| Eastern Europe | Belarus | -0.01 (-0.01, -0.00) | -0.00 (-0.01, -0.00) | 0.38 (0.38, 0.39) | 0.57 (0.56, 0.58) |
| Eastern Europe | Estonia | -0.04 (-0.05, -0.03) | -0.03 (-0.04, -0.03) | -0.01 (-0.04, 0.01) | -0.29 (-0.34, -0.26) |
| Eastern Europe | Latvia | 0.01 (0.00, 0.01) | 0.01 (0.01, 0.01) | 0.14 (0.09, 0.16) | -0.01 (-0.08, 0.03) |
| Eastern Europe | Lithuania | 0.00 (-0.01, 0.01) | 0.00 (-0.01, 0.01) | 0.32 (0.32, 0.33) | 0.46 (0.45, 0.47) |
| Eastern Europe | Republic of Moldova | 0.03 (0.03, 0.04) | 0.04 (0.04, 0.04) | 0.15 (0.12, 0.18) | 0.02 (-0.03, 0.09) |
| Eastern Europe | Russian Federation | -0.04 (-0.04, -0.04) | -0.03 (-0.03, -0.03) | 0.21 (0.20, 0.22) | 0.19 (0.17, 0.20) |
| Eastern Europe | Ukraine | -0.10 (-0.11, -0.10) | -0.09 (-0.10, -0.09) | 0.14 (0.13, 0.15) | 0.07 (0.05, 0.09) |
| High-income Asia Pacific | Brunei Darussalam | -0.01 (-0.02, -0.00) | -0.01 (-0.01, -0.00) | 0.12 (0.11, 0.13) | 0.33 (0.30, 0.36) |
| High-income Asia Pacific | Japan | 0.15 (0.12, 0.19) | 0.07 (0.04, 0.10) | 0.18 (0.14, 0.20) | 0.45 (0.42, 0.48) |
| High-income Asia Pacific | Republic of Korea | -0.04 (-0.05, -0.03) | -0.08 (-0.09, -0.07) | 0.16 (0.14, 0.18) | 0.64 (0.58, 0.69) |
| High-income Asia Pacific | Singapore | 0.11 (0.10, 0.12) | 0.12 (0.11, 0.12) | -0.21 (-0.22, -0.19) | -1.11 (-1.15, -1.07) |
| Australasia | Australia | -0.56 (-0.56, -0.55) | -0.58 (-0.58, -0.57) | 0.10 (0.07, 0.12) | 0.10 (0.05, 0.15) |
| Australasia | New Zealand | -0.20 (-0.20, -0.19) | -0.18 (-0.19, -0.18) | 0.07 (0.05, 0.08) | -0.16 (-0.20, -0.13) |
| Western Europe | Andorra | -0.21 (-0.22, -0.21) | -0.20 (-0.20, -0.19) | 0.27 (0.21, 0.35) | 0.47 (0.36, 0.62) |
| Western Europe | Austria | -0.22 (-0.22, -0.21) | -0.20 (-0.21, -0.20) | 0.14 (0.11, 0.18) | -0.09 (-0.18, -0.02) |
| Western Europe | Belgium | -0.41 (-0.41, -0.40) | -0.38 (-0.38, -0.38) | 0.34 (0.27, 0.43) | 0.88 (0.74, 1.06) |
| Western Europe | Cyprus | -0.14 (-0.14, -0.13) | -0.13 (-0.13, -0.12) | 0.20 (0.19, 0.22) | 0.33 (0.30, 0.37) |
| Western Europe | Denmark | -0.69 (-0.70, -0.68) | -0.64 (-0.66, -0.63) | -0.01 (-0.04, 0.02) | -0.51 (-0.56, -0.44) |
| Western Europe | Finland | -0.36 (-0.37, -0.36) | -0.36 (-0.36, -0.35) | 0.08 (0.03, 0.13) | 0.07 (-0.10, 0.20) |
| Western Europe | France | -0.17 (-0.18, -0.16) | -0.19 (-0.20, -0.19) | 0.19 (0.12, 0.26) | 0.17 (0.06, 0.30) |
| Western Europe | Germany | 0.05 (0.04, 0.05) | -0.07 (-0.08, -0.06) | 0.37 (0.34, 0.39) | 0.76 (0.71, 0.80) |
| Western Europe | Greece | -0.15 (-0.15, -0.15) | -0.13 (-0.14, -0.13) | 0.27 (0.23, 0.30) | 0.33 (0.24, 0.41) |
| Western Europe | Iceland | -0.34 (-0.35, -0.34) | -0.31 (-0.32, -0.31) | 0.06 (0.03, 0.10) | -0.01 (-0.07, 0.08) |
| Western Europe | Ireland | -0.30 (-0.30, -0.30) | -0.28 (-0.29, -0.28) | 0.32 (0.26, 0.38) | 0.70 (0.54, 0.85) |
| Western Europe | Israel | -0.20 (-0.20, -0.20) | -0.19 (-0.20, -0.19) | 0.15 (0.11, 0.19) | 0.13 (0.02, 0.22) |
| Western Europe | Italy | 0.30 (0.26, 0.32) | 0.47 (0.42, 0.51) | 0.17 (0.11, 0.25) | 0.33 (0.21, 0.46) |
| Western Europe | Luxembourg | -0.48 (-0.49, -0.47) | -0.46 (-0.47, -0.45) | 0.04 (-0.01, 0.10) | -0.28 (-0.36, -0.15) |
| Western Europe | Malta | -0.26 (-0.26, -0.25) | -0.24 (-0.25, -0.24) | 0.14 (0.11, 0.19) | 0.25 (0.18, 0.35) |
| Western Europe | Netherlands | -0.05 (-0.05, -0.04) | -0.15 (-0.16, -0.15) | 0.32 (0.27, 0.39) | 0.49 (0.40, 0.62) |
| Western Europe | Norway | -0.59 (-0.60, -0.58) | -0.57 (-0.58, -0.56) | 0.14 (0.12, 0.16) | 0.41 (0.38, 0.45) |
| Western Europe | Portugal | -0.07 (-0.07, -0.06) | -0.05 (-0.05, -0.05) | 0.28 (0.21, 0.34) | 0.30 (0.16, 0.41) |
| Western Europe | Spain | -0.54 (-0.61, -0.47) | -0.54 (-0.62, -0.46) | 0.31 (0.25, 0.36) | 0.90 (0.88, 0.92) |
| Western Europe | Sweden | -0.25 (-0.26, -0.24) | -0.27 (-0.28, -0.27) | 0.19 (0.15, 0.25) | 0.25 (0.15, 0.36) |
| Western Europe | Switzerland | -0.35 (-0.36, -0.33) | -0.30 (-0.31, -0.28) | 0.14 (0.12, 0.17) | 0.15 (0.03, 0.26) |
| Western Europe | United Kingdom | -0.19 (-0.19, -0.18) | -0.20 (-0.20, -0.19) | 0.34 (0.28, 0.40) | 0.31 (0.18, 0.45) |
| Southern Latin America | Argentina | -0.17 (-0.17, -0.16) | -0.16 (-0.16, -0.16) | 0.24 (0.18, 0.31) | 0.16 (0.06, 0.31) |
| Southern Latin America | Chile | -0.06 (-0.06, -0.05) | -0.06 (-0.06, -0.05) | 0.12 (0.03, 0.18) | -0.07 (-0.18, 0.08) |
| Southern Latin America | Uruguay | -0.17 (-0.18, -0.17) | -0.16 (-0.17, -0.15) | 0.36 (0.33, 0.37) | 0.85 (0.82, 0.87) |
| High-income North America | Canada | -0.36 (-0.38, -0.34) | -0.40 (-0.41, -0.38) | 0.18 (0.15, 0.24) | 0.56 (0.46, 0.68) |
| High-income North America | United States of America | -0.15 (-0.15, -0.14) | -0.18 (-0.18, -0.17) | 0.47 (0.41, 0.54) | 1.27 (1.14, 1.38) |
| Caribbean | Antigua and Barbuda | -0.08 (-0.08, -0.08) | -0.08 (-0.08, -0.08) | 0.24 (0.22, 0.26) | 0.45 (0.41, 0.51) |
| Caribbean | Bahamas | -0.07 (-0.07, -0.07) | -0.07 (-0.07, -0.06) | 0.29 (0.26, 0.34) | 0.53 (0.44, 0.64) |
| Caribbean | Barbados | -0.14 (-0.14, -0.13) | -0.13 (-0.14, -0.13) | 0.28 (0.27, 0.28) | 0.53 (0.52, 0.55) |
| Caribbean | Belize | -0.13 (-0.13, -0.12) | -0.13 (-0.13, -0.12) | 0.27 (0.21, 0.31) | 0.58 (0.45, 0.67) |
| Caribbean | Cuba | -0.01 (-0.06, 0.02) | -0.06 (-0.09, -0.03) | 0.09 (0.09, 0.10) | -0.09 (-0.12, -0.07) |
| Caribbean | Dominica | -0.10 (-0.10, -0.09) | -0.10 (-0.10, -0.09) | 0.23 (0.22, 0.24) | 0.43 (0.40, 0.47) |
| Caribbean | Dominican Republic | 0.00 (-0.04, 0.03) | -0.02 (-0.06, 0.00) | 0.28 (0.22, 0.36) | 0.63 (0.38, 0.78) |
| Caribbean | Grenada | -0.04 (-0.05, -0.04) | -0.05 (-0.05, -0.05) | 0.23 (0.22, 0.24) | 0.42 (0.40, 0.46) |
| Caribbean | Guyana | -0.03 (-0.04, -0.03) | -0.03 (-0.03, -0.03) | 0.42 (0.40, 0.43) | 0.82 (0.80, 0.85) |
| Caribbean | Haiti | -0.26 (-0.26, -0.25) | -0.25 (-0.25, -0.24) | 0.25 (0.23, 0.28) | 0.42 (0.37, 0.49) |
| Caribbean | Jamaica | -0.17 (-0.18, -0.17) | -0.17 (-0.18, -0.17) | 0.29 (0.28, 0.29) | 0.55 (0.54, 0.57) |
| Caribbean | Saint Lucia | -0.10 (-0.10, -0.10) | -0.10 (-0.10, -0.09) | 0.30 (0.29, 0.30) | 0.58 (0.57, 0.59) |
| Caribbean | Saint Vincent and the Grenadines | -0.17 (-0.17, -0.16) | -0.16 (-0.17, -0.16) | 0.26 (0.25, 0.27) | 0.59 (0.56, 0.62) |
| Caribbean | Suriname | -0.16 (-0.16, -0.16) | -0.15 (-0.15, -0.15) | 0.43 (0.42, 0.44) | 0.83 (0.81, 0.86) |
| Caribbean | Trinidad and Tobago | -0.08 (-0.08, -0.07) | -0.07 (-0.08, -0.07) | 0.32 (0.30, 0.33) | 0.53 (0.50, 0.54) |
| Andean Latin America | Bolivia (Plurinational State of) | -0.06 (-0.06, -0.05) | -0.05 (-0.05, -0.05) | 0.47 (0.36, 0.62) | 0.63 (0.45, 0.88) |
| Andean Latin America | Ecuador | -0.08 (-0.08, -0.08) | -0.08 (-0.08, -0.07) | 0.43 (0.32, 0.54) | 0.86 (0.66, 1.02) |
| Andean Latin America | Peru | -0.04 (-0.04, -0.04) | -0.03 (-0.04, -0.03) | 0.40 (0.31, 0.55) | 0.43 (0.26, 0.68) |
| Central Latin America | Colombia | -0.04 (-0.04, -0.04) | -0.04 (-0.05, -0.04) | 0.18 (0.13, 0.23) | 0.04 (-0.08, 0.12) |
| Central Latin America | Costa Rica | -0.08 (-0.08, -0.08) | -0.08 (-0.09, -0.08) | 0.34 (0.31, 0.39) | 0.65 (0.59, 0.73) |
| Central Latin America | El Salvador | 0.04 (0.03, 0.04) | 0.04 (0.03, 0.04) | 0.25 (0.19, 0.32) | 0.29 (0.19, 0.44) |
| Central Latin America | Guatemala | -0.03 (-0.04, -0.03) | -0.03 (-0.03, -0.03) | 0.26 (0.21, 0.33) | 0.37 (0.25, 0.48) |
| Central Latin America | Honduras | -0.06 (-0.06, -0.06) | -0.05 (-0.05, -0.04) | 0.43 (0.39, 0.48) | 0.83 (0.77, 0.94) |
| Central Latin America | Mexico | -0.29 (-0.30, -0.28) | -0.27 (-0.28, -0.27) | 0.53 (0.45, 0.61) | 0.93 (0.78, 1.08) |
| Central Latin America | Nicaragua | -0.03 (-0.03, -0.02) | -0.02 (-0.02, -0.02) | 0.31 (0.26, 0.38) | 0.55 (0.43, 0.66) |
| Central Latin America | Panama | -0.04 (-0.04, -0.04) | -0.05 (-0.05, -0.04) | 0.26 (0.18, 0.33) | 0.42 (0.27, 0.55) |
| Central Latin America | Venezuela (Bolivarian Republic of) | -0.07 (-0.07, -0.06) | -0.07 (-0.08, -0.07) | 0.15 (0.13, 0.18) | 0.23 (0.19, 0.29) |
| Tropical Latin America | Brazil | 0.00 (-0.02, 0.01) | -0.06 (-0.07, -0.05) | 0.56 (0.50, 0.60) | 0.33 (0.27, 0.39) |
| Tropical Latin America | Paraguay | -0.19 (-0.20, -0.19) | -0.20 (-0.20, -0.20) | 0.36 (0.35, 0.38) | 0.71 (0.67, 0.74) |
| North Africa and Middle East | Algeria | -0.14 (-0.15, -0.14) | -0.13 (-0.14, -0.13) | 0.11 (0.07, 0.15) | 0.23 (0.16, 0.32) |
| North Africa and Middle East | Bahrain | -0.10 (-0.10, -0.09) | -0.08 (-0.08, -0.07) | 0.07 (0.05, 0.11) | 0.02 (-0.03, 0.09) |
| North Africa and Middle East | Egypt | -0.12 (-0.13, -0.11) | -0.11 (-0.11, -0.10) | 0.19 (0.15, 0.23) | 0.64 (0.56, 0.74) |
| North Africa and Middle East | Iran (Islamic Republic of) | -0.12 (-0.12, -0.11) | -0.10 (-0.10, -0.09) | 0.23 (0.19, 0.28) | 0.36 (0.30, 0.41) |
| North Africa and Middle East | Iraq | -0.16 (-0.16, -0.15) | -0.14 (-0.15, -0.14) | 0.09 (0.03, 0.15) | 0.23 (0.10, 0.37) |
| North Africa and Middle East | Jordan | 0.01 (0.00, 0.02) | 0.03 (0.02, 0.04) | 0.02 (0.00, 0.04) | -0.10 (-0.13, -0.06) |
| North Africa and Middle East | Kuwait | -0.20 (-0.20, -0.19) | -0.15 (-0.15, -0.15) | 0.18 (0.14, 0.21) | 0.36 (0.30, 0.45) |
| North Africa and Middle East | Lebanon | -0.00 (-0.02, 0.01) | 0.01 (0.00, 0.02) | 0.43 (0.42, 0.44) | 0.89 (0.86, 0.92) |
| North Africa and Middle East | Libya | -0.18 (-0.18, -0.17) | -0.16 (-0.16, -0.15) | 0.34 (0.30, 0.39) | 0.54 (0.48, 0.64) |
| North Africa and Middle East | Morocco | -0.23 (-0.24, -0.23) | -0.22 (-0.22, -0.21) | 0.23 (0.16, 0.32) | 0.42 (0.30, 0.59) |
| North Africa and Middle East | Palestine | -0.13 (-0.14, -0.12) | -0.13 (-0.14, -0.12) | 0.16 (0.12, 0.20) | 0.34 (0.27, 0.42) |
| North Africa and Middle East | Oman | -0.26 (-0.26, -0.25) | -0.22 (-0.23, -0.21) | 0.25 (0.21, 0.30) | 0.47 (0.40, 0.56) |
| North Africa and Middle East | Qatar | -0.09 (-0.09, -0.08) | -0.07 (-0.08, -0.07) | 0.09 (0.07, 0.13) | 0.13 (0.08, 0.21) |
| North Africa and Middle East | Saudi Arabia | -0.21 (-0.22, -0.20) | -0.19 (-0.19, -0.18) | 0.24 (0.19, 0.31) | 0.56 (0.46, 0.70) |
| North Africa and Middle East | Syrian Arab Republic | -0.27 (-0.28, -0.27) | -0.23 (-0.24, -0.23) | 0.20 (0.18, 0.23) | 0.35 (0.30, 0.42) |
| North Africa and Middle East | Tunisia | -0.22 (-0.22, -0.22) | -0.20 (-0.20, -0.20) | 0.32 (0.31, 0.32) | 0.59 (0.57, 0.61) |
| North Africa and Middle East | Türkiye | -0.21 (-0.21, -0.20) | -0.19 (-0.19, -0.18) | 0.19 (0.17, 0.21) | 0.29 (0.24, 0.32) |
| North Africa and Middle East | United Arab Emirates | -0.34 (-0.34, -0.33) | -0.28 (-0.28, -0.27) | 0.14 (0.11, 0.18) | 0.19 (0.13, 0.27) |
| North Africa and Middle East | Yemen | -0.31 (-0.32, -0.31) | -0.26 (-0.27, -0.26) | 0.09 (0.07, 0.12) | 0.23 (0.15, 0.30) |
| North Africa and Middle East | Afghanistan | -0.07 (-0.07, -0.07) | -0.05 (-0.05, -0.05) | 0.29 (0.26, 0.33) | 0.46 (0.41, 0.52) |
| South Asia | Bangladesh | -0.13 (-0.13, -0.13) | -0.13 (-0.13, -0.12) | 0.24 (0.20, 0.30) | 0.54 (0.47, 0.64) |
| South Asia | Bhutan | -0.27 (-0.28, -0.27) | -0.26 (-0.26, -0.25) | -0.11 (-0.13, -0.08) | -0.12 (-0.16, -0.07) |
| South Asia | India | -0.02 (-0.03, 0.01) | -0.02 (-0.03, -0.01) | -0.01 (-0.04, 0.04) | 0.17 (0.03, 0.31) |
| South Asia | Nepal | -0.38 (-0.38, -0.38) | -0.37 (-0.37, -0.37) | 0.38 (0.32, 0.44) | 0.78 (0.66, 0.89) |
| South Asia | Pakistan | -0.23 (-0.23, -0.22) | -0.22 (-0.22, -0.22) | 0.19 (0.15, 0.23) | 0.10 (0.01, 0.17) |
| Central Sub-Saharan Africa | Angola | -0.09 (-0.10, -0.09) | -0.08 (-0.08, -0.08) | 0.16 (0.14, 0.20) | 0.21 (0.17, 0.27) |
| Central Sub-Saharan Africa | Central African Republic | -0.02 (-0.03, -0.01) | -0.05 (-0.05, -0.05) | 0.14 (0.10, 0.19) | 0.15 (0.08, 0.23) |
| Central Sub-Saharan Africa | Congo | -0.16 (-0.17, -0.16) | -0.11 (-0.11, -0.10) | 0.10 (0.07, 0.14) | 0.05 (-0.02, 0.12) |
| Central Sub-Saharan Africa | Democratic Republic of the Congo | 0.04 (0.04, 0.04) | 0.01 (0.01, 0.02) | 0.14 (0.10, 0.18) | 0.18 (0.13, 0.24) |
| Central Sub-Saharan Africa | Equatorial Guinea | -0.15 (-0.15, -0.14) | -0.12 (-0.13, -0.12) | 0.12 (0.08, 0.17) | 0.15 (0.09, 0.24) |
| Central Sub-Saharan Africa | Gabon | -0.08 (-0.08, -0.08) | -0.07 (-0.07, -0.07) | 0.17 (0.16, 0.18) | 0.19 (0.18, 0.21) |
| Eastern Sub-Saharan Africa | Burundi | -0.27 (-0.28, -0.27) | -0.26 (-0.26, -0.26) | -0.06 (-0.08, -0.04) | -0.35 (-0.39, -0.31) |
| Eastern Sub-Saharan Africa | Comoros | -0.16 (-0.16, -0.16) | -0.13 (-0.14, -0.13) | 0.16 (0.14, 0.18) | 0.17 (0.12, 0.21) |
| Eastern Sub-Saharan Africa | Djibouti | -0.18 (-0.18, -0.18) | -0.17 (-0.17, -0.16) | 0.18 (0.14, 0.21) | 0.30 (0.25, 0.36) |
| Eastern Sub-Saharan Africa | Eritrea | -0.21 (-0.21, -0.21) | -0.19 (-0.19, -0.19) | 0.08 (0.07, 0.08) | 0.08 (0.07, 0.08) |
| Eastern Sub-Saharan Africa | Ethiopia | -0.27 (-0.27, -0.27) | -0.24 (-0.24, -0.24) | 0.05 (0.03, 0.08) | 0.01 (-0.03, 0.05) |
| Eastern Sub-Saharan Africa | Kenya | -0.04 (-0.04, -0.04) | -0.06 (-0.06, -0.05) | 0.12 (0.08, 0.16) | 0.05 (0.00, 0.10) |
| Eastern Sub-Saharan Africa | Madagascar | -0.17 (-0.17, -0.17) | -0.17 (-0.17, -0.16) | 0.18 (0.16, 0.21) | 0.21 (0.17, 0.26) |
| Eastern Sub-Saharan Africa | Malawi | -0.01 (-0.02, -0.01) | -0.02 (-0.02, -0.02) | 0.16 (0.16, 0.16) | 0.14 (0.12, 0.15) |
| Southeast Asia | Mauritius | -0.03 (-0.03, -0.03) | -0.02 (-0.02, -0.02) | 0.09 (0.08, 0.11) | 0.08 (0.03, 0.13) |
| Eastern Sub-Saharan Africa | Mozambique | -0.10 (-0.11, -0.10) | -0.10 (-0.11, -0.10) | 0.28 (0.27, 0.28) | 0.52 (0.51, 0.52) |
| Eastern Sub-Saharan Africa | Rwanda | -0.02 (-0.02, -0.02) | -0.01 (-0.02, -0.01) | -0.02 (-0.02, -0.01) | -0.21 (-0.23, -0.20) |
| Southeast Asia | Seychelles | -0.12 (-0.12, -0.12) | -0.11 (-0.11, -0.11) | 0.17 (0.15, 0.18) | 0.20 (0.17, 0.23) |
| Eastern Sub-Saharan Africa | Somalia | -0.03 (-0.03, -0.03) | -0.03 (-0.03, -0.03) | 0.28 (0.28, 0.28) | 0.43 (0.42, 0.43) |
| Eastern Sub-Saharan Africa | United Republic of Tanzania | -0.27 (-0.27, -0.27) | -0.27 (-0.27, -0.27) | 0.13 (0.12, 0.14) | 0.13 (0.11, 0.15) |
| Eastern Sub-Saharan Africa | Uganda | -0.04 (-0.04, -0.04) | -0.04 (-0.05, -0.04) | 0.19 (0.17, 0.21) | 0.23 (0.20, 0.26) |
| Eastern Sub-Saharan Africa | Zambia | 0.01 (0.01, 0.02) | 0.01 (0.01, 0.01) | 0.25 (0.23, 0.26) | 0.34 (0.32, 0.36) |
| Southern Sub-Saharan Africa | Botswana | -0.14 (-0.14, -0.13) | -0.13 (-0.13, -0.13) | 0.32 (0.25, 0.35) | 0.56 (0.48, 0.60) |
| Southern Sub-Saharan Africa | Lesotho | -0.06 (-0.06, -0.05) | -0.05 (-0.05, -0.04) | 0.34 (0.31, 0.36) | 0.53 (0.50, 0.56) |
| Southern Sub-Saharan Africa | Namibia | -0.12 (-0.12, -0.12) | -0.11 (-0.12, -0.11) | 0.34 (0.31, 0.36) | 0.53 (0.48, 0.55) |
| Southern Sub-Saharan Africa | South Africa | -0.19 (-0.19, -0.19) | -0.19 (-0.19, -0.18) | 0.33 (0.29, 0.39) | 0.52 (0.44, 0.63) |
| Southern Sub-Saharan Africa | Eswatini | -0.14 (-0.14, -0.13) | -0.13 (-0.13, -0.12) | 0.51 (0.50, 0.51) | 0.92 (0.90, 0.95) |
| Southern Sub-Saharan Africa | Zimbabwe | -0.14 (-0.15, -0.14) | -0.12 (-0.12, -0.12) | 0.25 (0.24, 0.25) | 0.47 (0.46, 0.49) |
| Western Sub-Saharan Africa | Benin | -0.36 (-0.37, -0.35) | -0.36 (-0.37, -0.35) | 0.19 (0.18, 0.20) | 0.36 (0.34, 0.38) |
| Western Sub-Saharan Africa | Burkina Faso | -0.23 (-0.23, -0.22) | -0.21 (-0.22, -0.21) | 0.08 (0.06, 0.10) | 0.09 (0.05, 0.13) |
| Western Sub-Saharan Africa | Cameroon | -0.18 (-0.18, -0.18) | -0.18 (-0.18, -0.18) | 0.19 (0.17, 0.20) | 0.33 (0.31, 0.36) |
| Western Sub-Saharan Africa | Cabo Verde | -0.18 (-0.18, -0.17) | -0.17 (-0.17, -0.17) | 0.37 (0.34, 0.42) | 0.70 (0.65, 0.79) |
| Western Sub-Saharan Africa | Chad | -0.32 (-0.32, -0.32) | -0.30 (-0.31, -0.30) | 0.13 (0.10, 0.17) | 0.22 (0.16, 0.27) |
| Western Sub-Saharan Africa | Côte d'Ivoire | -0.13 (-0.13, -0.13) | -0.12 (-0.12, -0.11) | 0.11 (0.09, 0.12) | 0.14 (0.11, 0.16) |
| Western Sub-Saharan Africa | Gambia | -0.26 (-0.26, -0.26) | -0.25 (-0.25, -0.25) | 0.21 (0.18, 0.24) | 0.33 (0.29, 0.38) |
| Western Sub-Saharan Africa | Ghana | -0.08 (-0.08, -0.07) | -0.08 (-0.08, -0.07) | 0.19 (0.16, 0.22) | 0.32 (0.27, 0.37) |
| Western Sub-Saharan Africa | Guinea | -0.20 (-0.20, -0.20) | -0.19 (-0.19, -0.19) | 0.23 (0.21, 0.26) | 0.43 (0.39, 0.48) |
| Western Sub-Saharan Africa | Guinea-Bissau | -0.10 (-0.10, -0.09) | -0.09 (-0.09, -0.08) | 0.23 (0.20, 0.26) | 0.40 (0.36, 0.46) |
| Western Sub-Saharan Africa | Liberia | -0.12 (-0.12, -0.11) | -0.11 (-0.12, -0.11) | 0.19 (0.16, 0.22) | 0.28 (0.24, 0.34) |
| Western Sub-Saharan Africa | Mali | -0.15 (-0.16, -0.15) | -0.14 (-0.14, -0.13) | 0.09 (0.07, 0.13) | 0.09 (0.01, 0.16) |
| Western Sub-Saharan Africa | Mauritania | -0.25 (-0.26, -0.25) | -0.24 (-0.24, -0.24) | 0.09 (0.06, 0.12) | 0.13 (0.09, 0.19) |
| Western Sub-Saharan Africa | Niger | -0.22 (-0.22, -0.22) | -0.19 (-0.20, -0.19) | 0.12 (0.11, 0.13) | 0.21 (0.17, 0.25) |
| Western Sub-Saharan Africa | Nigeria | -0.32 (-0.33, -0.32) | -0.30 (-0.31, -0.29) | -0.05 (-0.07, -0.02) | -0.20 (-0.26, -0.15) |
| Western Sub-Saharan Africa | Sao Tome and Principe | -0.24 (-0.25, -0.24) | -0.22 (-0.22, -0.21) | 0.13 (0.11, 0.15) | 0.24 (0.18, 0.29) |
| Western Sub-Saharan Africa | Senegal | -0.24 (-0.24, -0.23) | -0.23 (-0.23, -0.22) | 0.24 (0.21, 0.27) | 0.40 (0.34, 0.45) |
| Western Sub-Saharan Africa | Sierra Leone | -0.19 (-0.20, -0.19) | -0.18 (-0.18, -0.18) | 0.18 (0.14, 0.21) | 0.36 (0.32, 0.41) |
| Western Sub-Saharan Africa | Togo | -0.12 (-0.13, -0.11) | -0.12 (-0.12, -0.11) | 0.18 (0.15, 0.20) | 0.32 (0.29, 0.38) |
| Oceania | American Samoa | -0.07 (-0.07, -0.07) | -0.06 (-0.06, -0.06) | 0.12 (0.09, 0.13) | 0.18 (0.14, 0.21) |
| Caribbean | Bermuda | -0.05 (-0.06, -0.05) | -0.05 (-0.05, -0.05) | 0.15 (0.14, 0.17) | 0.08 (0.05, 0.12) |
| Oceania | Cook Islands | -0.07 (-0.07, -0.06) | -0.06 (-0.06, -0.06) | 0.15 (0.13, 0.16) | 0.27 (0.23, 0.29) |
| High-income North America | Greenland | -0.08 (-0.09, -0.07) | -0.07 (-0.07, -0.06) | 0.25 (0.21, 0.32) | 0.40 (0.31, 0.52) |
| Oceania | Guam | -0.01 (-0.02, -0.01) | -0.02 (-0.02, -0.02) | 0.23 (0.22, 0.26) | 0.45 (0.42, 0.49) |
| Western Europe | Monaco | -0.28 (-0.28, -0.27) | -0.26 (-0.26, -0.26) | 0.26 (0.24, 0.29) | 0.47 (0.43, 0.52) |
| Oceania | Nauru | 0.10 (0.09, 0.10) | 0.09 (0.09, 0.09) | 0.19 (0.16, 0.20) | 0.33 (0.29, 0.35) |
| Oceania | Niue | -0.10 (-0.10, -0.10) | -0.09 (-0.09, -0.09) | 0.11 (0.09, 0.12) | 0.23 (0.19, 0.25) |
| Oceania | Northern Mariana Islands | -0.05 (-0.06, -0.05) | -0.05 (-0.06, -0.04) | 0.27 (0.25, 0.28) | 0.67 (0.64, 0.69) |
| Oceania | Palau | -0.12 (-0.12, -0.11) | -0.10 (-0.10, -0.09) | 0.09 (0.07, 0.10) | 0.18 (0.14, 0.21) |
| Caribbean | Puerto Rico | -0.07 (-0.07, -0.07) | -0.07 (-0.08, -0.07) | 0.21 (0.17, 0.27) | 0.31 (0.23, 0.42) |
| Caribbean | Saint Kitts and Nevis | -0.04 (-0.04, -0.04) | -0.03 (-0.04, -0.03) | 0.22 (0.19, 0.25) | 0.39 (0.34, 0.47) |
| Western Europe | San Marino | -0.40 (-0.40, -0.39) | -0.37 (-0.37, -0.36) | 0.34 (0.28, 0.43) | 0.62 (0.51, 0.78) |
| Oceania | Tokelau | -0.07 (-0.07, -0.06) | -0.06 (-0.06, -0.05) | 0.10 (0.08, 0.11) | 0.19 (0.15, 0.21) |
| Oceania | Tuvalu | -0.09 (-0.09, -0.09) | -0.07 (-0.08, -0.07) | 0.08 (0.05, 0.09) | 0.17 (0.12, 0.19) |
| Caribbean | United States Virgin Islands | -0.10 (-0.10, -0.10) | -0.10 (-0.11, -0.10) | 0.27 (0.25, 0.30) | 0.53 (0.49, 0.59) |
| Eastern Sub-Saharan Africa | South Sudan | -0.24 (-0.24, -0.23) | -0.20 (-0.21, -0.19) | 0.21 (0.18, 0.25) | 0.30 (0.24, 0.37) |
| North Africa and Middle East | Sudan | -0.25 (-0.26, -0.25) | -0.22 (-0.23, -0.22) | 0.10 (0.05, 0.15) | 0.43 (0.34, 0.56) |

Supplementary Table 4: Global and regional AAPC of ASR for dementia

| **Region** | **Prevalence** | | | **Incidence** | | |
| --- | --- | --- | --- | --- | --- | --- |
|  | **ASR in 1990** | **ASR in 2021** | **AAPC (95%UI)** | **ASR in 1990** | **ASR in 2021** | **AAPC (95%UI)** |
| Global | 672.22 | 694.01 | 0.09 (0.06, 0.11) | 116.97 | 119.76 | 0.06 (0.04, 0.08) |
| East Asia | 697.26 | 887.95 | 0.73 (0.64, 0.81) | 120.29 | 149.61 | 0.65 (0.57, 0.71) |
| Southeast Asia | 675.02 | 644.38 | -0.15 (-0.15, -0.15) | 114.85 | 110.07 | -0.14 (-0.14, -0.13) |
| Oceania | 676.80 | 644.83 | -0.16 (-0.16, -0.15) | 117.17 | 112.05 | -0.14 (-0.15, -0.14) |
| Central Asia | 638.07 | 626.81 | -0.05 (-0.06, -0.05) | 111.72 | 109.82 | -0.05 (-0.06, -0.05) |
| Central Europe | 657.16 | 641.22 | -0.08 (-0.08, -0.08) | 115.01 | 112.42 | -0.07 (-0.07, -0.07) |
| Eastern Europe | 669.20 | 658.68 | -0.05 (-0.05, -0.05) | 117.31 | 115.66 | -0.04 (-0.05, -0.04) |
| High-income Asia Pacific | 658.06 | 684.81 | 0.15 (0.13, 0.19) | 116.94 | 118.62 | 0.07 (0.04, 0.10) |
| Australasia | 706.96 | 604.41 | -0.50 (-0.51, -0.49) | 123.79 | 105.44 | -0.51 (-0.52, -0.51) |
| Western Europe | 691.03 | 670.36 | -0.09 (-0.11, -0.08) | 122.45 | 118.56 | -0.10 (-0.11, -0.09) |
| Southern Latin America | 622.39 | 595.26 | -0.14 (-0.14, -0.14) | 111.77 | 107.09 | -0.13 (-0.14, -0.13) |
| High-income North America | 815.90 | 775.11 | -0.17 (-0.17, -0.16) | 139.57 | 131.39 | -0.20 (-0.20, -0.19) |
| Caribbean | 557.24 | 550.22 | -0.06 (-0.09, -0.04) | 97.60 | 95.60 | -0.08 (-0.10, -0.07) |
| Andean Latin America | 450.85 | 444.09 | -0.05 (-0.05, -0.05) | 80.60 | 79.55 | -0.04 (-0.05, -0.04) |
| Central Latin America | 625.86 | 596.48 | -0.15 (-0.16, -0.15) | 111.25 | 106.12 | -0.15 (-0.16, -0.15) |
| Tropical Latin America | 758.68 | 759.83 | -0.00 (-0.02, 0.01) | 129.14 | 126.83 | -0.06 (-0.07, -0.05) |
| North Africa and Middle East | 812.51 | 772.66 | -0.16 (-0.17, -0.16) | 138.06 | 132.19 | -0.14 (-0.14, -0.13) |
| South Asia | 446.47 | 437.07 | -0.06 (-0.07, -0.04) | 80.57 | 79.00 | -0.06 (-0.07, -0.06) |
| Central Sub-Saharan Africa | 752.86 | 750.54 | -0.01 (-0.01, -0.01) | 126.90 | 126.14 | -0.02 (-0.02, -0.02) |
| Eastern Sub-Saharan Africa | 618.54 | 588.72 | -0.16 (-0.16, -0.16) | 107.12 | 102.41 | -0.14 (-0.15, -0.14) |
| Southern Sub-Saharan Africa | 639.88 | 606.69 | -0.17 (-0.17, -0.17) | 112.76 | 107.13 | -0.16 (-0.17, -0.16) |
| Western Sub-Saharan Africa | 436.85 | 406.02 | -0.24 (-0.24, -0.24) | 78.42 | 73.18 | -0.22 (-0.23, -0.22) |

Note: AAPC: Average Annual Percentage Change; ASR: Age-Standardized Rate.

Supplementary Table 5: Global and regional AAPC of ASR for overall mental disorders.

| **Region** | **Prevalence** | | | **Incidence** | | |
| --- | --- | --- | --- | --- | --- | --- |
|  | **ASR in 1990** | **ASR in 2021** | **AAPC (95%UI)** | **ASR in 1990** | **ASR in 2021** | **AAPC (95%UI)** |
| Global | 12884.70 | 13554.28 | 0.19 (0.16, 0.24) | 4738.46 | 5459.50 | 0.48 (0.40, 0.61) |
| East Asia | 11338.87 | 11399.07 | 0.01 (-0.02, 0.02) | 3537.47 | 3343.70 | -0.17 (-0.22, -0.12) |
| Southeast Asia | 11093.86 | 11484.56 | 0.11 (0.11, 0.11) | 3167.74 | 3720.07 | 0.53 (0.52, 0.54) |
| Oceania | 11288.62 | 11726.42 | 0.11 (0.08, 0.13) | 3775.38 | 3981.87 | 0.14 (0.08, 0.17) |
| Central Asia | 10507.24 | 11097.81 | 0.21 (0.18, 0.24) | 4448.82 | 4994.23 | 0.43 (0.36, 0.51) |
| Central Europe | 11024.53 | 12031.98 | 0.30 (0.28, 0.33) | 3801.47 | 4362.91 | 0.48 (0.44, 0.55) |
| Eastern Europe | 11656.32 | 13057.16 | 0.37 (0.36, 0.38) | 5101.54 | 6012.77 | 0.54 (0.52, 0.55) |
| High-income Asia Pacific | 10647.10 | 11429.55 | 0.23 (0.22, 0.24) | 3275.55 | 3948.27 | 0.64 (0.61, 0.68) |
| Australasia | 17886.22 | 18828.43 | 0.18 (0.16, 0.20) | 6766.62 | 7325.33 | 0.28 (0.23, 0.36) |
| Western Europe | 15151.49 | 16750.75 | 0.39 (0.33, 0.48) | 6120.27 | 7218.52 | 0.72 (0.60, 0.90) |
| Southern Latin America | 13783.94 | 15178.17 | 0.39 (0.30, 0.49) | 4980.12 | 5774.98 | 0.57 (0.42, 0.79) |
| High-income North America | 14962.40 | 17809.06 | 0.64 (0.55, 0.72) | 5204.91 | 8145.38 | 1.60 (1.43, 1.83) |
| Caribbean | 14638.94 | 15673.38 | 0.24 (0.21, 0.29) | 5803.28 | 6307.97 | 0.33 (0.26, 0.42) |
| Andean Latin America | 13798.73 | 15898.12 | 0.58 (0.44, 0.76) | 4244.05 | 5347.34 | 0.82 (0.52, 1.06) |
| Central Latin America | 11765.95 | 13577.02 | 0.53 (0.44, 0.62) | 4458.30 | 5849.00 | 0.98 (0.84, 1.19) |
| Tropical Latin America | 15370.81 | 18385.63 | 0.63 (0.58, 0.68) | 6084.25 | 7097.67 | 0.59 (0.48, 0.70) |
| North Africa and Middle East | 15344.99 | 16365.24 | 0.27 (0.22, 0.35) | 6334.70 | 7336.05 | 0.57 (0.45, 0.72) |
| South Asia | 14524.32 | 14214.83 | -0.04 (-0.08, 0.02) | 5731.60 | 6133.77 | 0.31 (0.16, 0.46) |
| Central Sub-Saharan Africa | 13556.71 | 14276.77 | 0.21 (0.16, 0.28) | 8274.33 | 8706.47 | 0.23 (0.15, 0.33) |
| Eastern Sub-Saharan Africa | 13106.03 | 13796.54 | 0.18 (0.17, 0.21) | 7018.40 | 7528.81 | 0.26 (0.23, 0.30) |
| Southern Sub-Saharan Africa | 11879.22 | 13539.90 | 0.47 (0.44, 0.53) | 5701.09 | 7052.59 | 0.81 (0.73, 0.91) |
| Western Sub-Saharan Africa | 11332.10 | 11554.29 | 0.09 (0.06, 0.13) | 5617.47 | 5658.50 | 0.09 (0.03, 0.16) |

Note: AAPC: Average Annual Percentage Change; ASR: Age-Standardized Rate.

Supplementary Table 6: Global and regional AAPC of ASR for eight mental disorders subtypes.

| **Region** | **Mental Disorders** | | | | | | | |
| --- | --- | --- | --- | --- | --- | --- | --- | --- |
|  | **SCZ** | **DD** | **BD** | **ANX** | **ED** | **ASD** | **ADHD** | **IDID** |
| ASPR | | | | | | | | |
| Global | 0.06 (0.06, 0.06) | 0.28 (0.23, 0.33) | -0.12 (-0.12, -0.12) | 0.43 (0.38, 0.48) | 0.40 (0.39, 0.41) | 0.05 (0.05, 0.05) | -0.12 (-0.13, -0.12) | -0.81 (-0.82, -0.80) |
| East Asia | 0.11 (0.10, 0.12) | 0.19 (0.16, 0.21) | -0.01 (-0.01, -0.01) | 0.05 (0.01, 0.08) | 2.02 (2.00, 2.04) | 0.29 (0.28, 0.29) | -0.30 (-0.32, -0.29) | -1.22 (-1.23, -1.20) |
| Southeast Asia | 0.12 (0.09, 0.14) | 0.15 (0.14, 0.15) | -0.00 (-0.00, -0.00) | 0.49 (0.48, 0.49) | 1.31 (1.30, 1.32) | 0.18 (0.17, 0.18) | -0.09 (-0.09, -0.09) | -2.38 (-2.41, -2.36) |
| Oceania | -0.01 (-0.02, 0.00) | 0.05 (0.02, 0.06) | 0.00 (0.00, 0.00) | 0.22 (0.18, 0.24) | 0.21 (0.20, 0.21) | 0.00 (-0.00, 0.00) | -0.02 (-0.02, -0.02) | 0.01 (-0.01, 0.02) |
| Central Asia | 0.04 (0.04, 0.05) | 0.09 (0.07, 0.13) | -0.00 (-0.00, -0.00) | 0.41 (0.37, 0.46) | 0.45 (0.42, 0.48) | 0.09 (0.09, 0.09) | 0.00 (0.00, 0.00) | -0.56 (-0.59, -0.53) |
| Central Europe | 0.09 (0.09, 0.09) | 0.09 (0.07, 0.12) | -0.00 (-0.00, -0.00) | 0.56 (0.53, 0.61) | 0.90 (0.88, 0.91) | 0.19 (0.18, 0.19) | 0.02 (0.02, 0.02) | -1.52 (-1.54, -1.51) |
| Eastern Europe | 0.15 (0.14, 0.16) | 0.05 (0.04, 0.06) | -0.00 (-0.00, -0.00) | 0.59 (0.58, 0.60) | 0.22 (0.19, 0.24) | 0.16 (0.15, 0.16) | 0.01 (0.01, 0.01) | -0.26 (-0.29, -0.22) |
| High-income Asia Pacific | -0.09 (-0.10, -0.08) | 0.28 (0.26, 0.30) | -0.05 (-0.05, -0.05) | 0.35 (0.28, 0.39) | 0.72 (0.71, 0.73) | 0.33 (0.32, 0.33) | 0.05 (0.04, 0.05) | -5.12 (-5.20, -5.04) |
| Australasia | 0.01 (0.01, 0.01) | 0.05 (-0.01, 0.09) | 0.01 (0.01, 0.01) | 0.17 (0.13, 0.21) | 0.85 (0.83, 0.88) | 0.13 (0.12, 0.13) | -0.01 (-0.01, -0.00) | 1.84 (1.76, 1.91) |
| Western Europe | -0.03 (-0.03, -0.02) | 0.33 (0.25, 0.42) | 0.04 (0.04, 0.04) | 0.49 (0.42, 0.60) | 0.59 (0.58, 0.60) | 0.19 (0.18, 0.19) | 0.24 (0.24, 0.25) | -0.88 (-0.92, -0.87) |
| Southern Latin America | 0.01 (0.01, 0.02) | 0.12 (0.06, 0.20) | -0.06 (-0.06, -0.06) | 0.61 (0.48, 0.73) | 0.62 (0.60, 0.63) | 0.14 (0.14, 0.14) | -0.02 (-0.02, -0.01) | -0.83 (-0.85, -0.81) |
| High-income North America | -0.06 (-0.07, -0.06) | 0.78 (0.70, 0.90) | -0.02 (-0.02, -0.02) | 0.57 (0.45, 0.69) | -0.02 (-0.03, -0.01) | 0.13 (0.13, 0.14) | 0.32 (0.30, 0.34) | -0.47 (-0.50, -0.46) |
| Caribbean | 0.01 (0.01, 0.02) | 0.17 (0.12, 0.22) | -0.01 (-0.01, -0.01) | 0.49 (0.46, 0.54) | 0.22 (0.21, 0.23) | 0.02 (0.02, 0.02) | -0.00 (-0.00, -0.00) | 0.06 (0.05, 0.07) |
| Andean Latin America | 0.05 (0.05, 0.05) | 0.40 (0.27, 0.54) | 0.00 (0.00, 0.00) | 0.87 (0.70, 1.11) | 0.88 (0.87, 0.89) | 0.18 (0.18, 0.18) | -0.01 (-0.01, -0.01) | -1.12 (-1.14, -1.11) |
| Central Latin America | 0.02 (0.02, 0.02) | 0.48 (0.40, 0.57) | 0.01 (0.01, 0.01) | 0.80 (0.70, 0.94) | 0.31 (0.30, 0.32) | 0.10 (0.10, 0.11) | -0.05 (-0.05, -0.04) | -0.68 (-0.71, -0.67) |
| Tropical Latin America | 0.05 (0.04, 0.05) | 0.24 (0.18, 0.29) | 0.00 (0.00, 0.00) | 1.20 (1.11, 1.28) | 0.56 (0.55, 0.57) | 0.08 (0.08, 0.09) | 0.00 (-0.00, 0.01) | -0.79 (-0.80, -0.78) |
| North Africa and Middle East | 0.05 (0.05, 0.05) | 0.27 (0.21, 0.34) | 0.00 (0.00, 0.00) | 0.51 (0.44, 0.60) | 0.71 (0.70, 0.71) | 0.19 (0.19, 0.19) | -0.04 (-0.04, -0.03) | -1.15 (-1.16, -1.14) |
| South Asia | 0.12 (0.11, 0.13) | 0.16 (0.07, 0.25) | -0.01 (-0.01, -0.01) | 0.51 (0.42, 0.60) | 1.66 (1.65, 1.67) | 0.09 (0.08, 0.09) | 0.01 (0.01, 0.01) | -1.15 (-1.16, -1.14) |
| Central Sub-Saharan Africa | -0.04 (-0.05, -0.04) | 0.13 (0.09, 0.18) | -0.00 (-0.00, -0.00) | 0.30 (0.24, 0.39) | -0.15 (-0.17, -0.14) | 0.15 (0.14, 0.15) | 0.06 (0.06, 0.06) | 1.78 (1.75, 1.82) |
| Eastern Sub-Saharan Africa | 0.05 (0.04, 0.05) | 0.09 (0.07, 0.11) | -0.00 (-0.00, -0.00) | 0.40 (0.36, 0.43) | 0.64 (0.63, 0.65) | 0.12 (0.12, 0.12) | -0.01 (-0.01, -0.01) | -0.08 (-0.10, -0.06) |
| Southern Sub-Saharan Africa | 0.02 (0.02, 0.03) | 0.38 (0.34, 0.45) | 0.00 (0.00, 0.00) | 0.64 (0.58, 0.71) | 0.23 (0.22, 0.24) | 0.07 (0.07, 0.08) | 0.00 (-0.00, 0.00) | -0.14 (-0.15, -0.13) |
| Western Sub-Saharan Africa | 0.09 (0.09, 0.09) | 0.04 (0.01, 0.07) | 0.00 (0.00, 0.00) | 0.21 (0.15, 0.27) | 0.59 (0.58, 0.60) | 0.01 (0.01, 0.02) | -0.13 (-0.13, -0.13) | 0.28 (0.26, 0.30) |
| ASIR | | | | | | | | |
| Global | -0.08 (-0.09, -0.08) | 0.38 (0.30, 0.47) | -0.07 (-0.08, -0.07) | 0.46 (0.40, 0.52) | 0.34 (0.34, 0.35) | NA | NA | NA |
| East Asia | -0.26 (-0.27, -0.25) | 0.41 (0.33, 0.48) | -0.01 (-0.01, -0.01) | 0.06 (0.03, 0.08) | 1.02 (1.01, 1.03) | NA | NA | NA |
| Southeast Asia | -0.14 (-0.16, -0.13) | 0.31 (0.30, 0.32) | -0.00 (-0.00, -0.00) | 0.49 (0.47, 0.50) | 0.58 (0.57, 0.58) | NA | NA | NA |
| Oceania | -0.07 (-0.09, -0.06) | 0.09 (0.04, 0.12) | 0.00 (0.00, 0.00) | 0.23 (0.19, 0.25) | 0.07 (0.06, 0.08) | NA | NA | NA |
| Central Asia | -0.02 (-0.03, -0.02) | 0.14 (0.11, 0.19) | 0.00 (0.00, 0.00) | 0.46 (0.42, 0.51) | 0.16 (0.13, 0.17) | NA | NA | NA |
| Central Europe | -0.03 (-0.04, -0.02) | 0.16 (0.13, 0.21) | 0.00 (0.00, 0.00) | 0.63 (0.60, 0.68) | 0.46 (0.45, 0.46) | NA | NA | NA |
| Eastern Europe | 0.00 (0.00, 0.01) | 0.07 (0.06, 0.09) | -0.00 (-0.00, -0.00) | 0.67 (0.66, 0.69) | 0.00 (-0.01, 0.02) | NA | NA | NA |
| High-income Asia Pacific | 0.02 (0.00, 0.03) | 0.43 (0.38, 0.48) | 0.03 (0.02, 0.03) | 0.29 (0.25, 0.32) | 0.34 (0.33, 0.35) | NA | NA | NA |
| Australasia | 0.06 (0.05, 0.08) | 0.06 (-0.03, 0.12) | 0.12 (0.11, 0.12) | 0.09 (0.04, 0.13) | 0.08 (0.05, 0.11) | NA | NA | NA |
| Western Europe | 0.01 (0.00, 0.01) | 0.43 (0.31, 0.55) | 0.11 (0.11, 0.12) | 0.55 (0.43, 0.63) | 0.18 (0.18, 0.19) | NA | NA | NA |
| Southern Latin America | -0.03 (-0.04, -0.02) | 0.17 (0.08, 0.31) | 0.04 (0.04, 0.04) | 0.61 (0.49, 0.75) | 0.13 (0.12, 0.14) | NA | NA | NA |
| High-income North America | 0.08 (0.07, 0.09) | 1.28 (1.13, 1.40) | -0.03 (-0.05, -0.01) | 0.57 (0.48, 0.67) | 0.44 (0.42, 0.45) | NA | NA | NA |
| Caribbean | 0.03 (0.02, 0.03) | 0.23 (0.17, 0.30) | 0.01 (0.01, 0.01) | 0.46 (0.43, 0.51) | 0.09 (0.08, 0.09) | NA | NA | NA |
| Andean Latin America | -0.06 (-0.07, -0.06) | 0.58 (0.39, 0.78) | -0.00 (-0.00, -0.00) | 0.89 (0.72, 1.14) | 0.33 (0.32, 0.34) | NA | NA | NA |
| Central Latin America | -0.02 (-0.04, -0.01) | 0.62 (0.52, 0.73) | 0.00 (0.00, 0.01) | 0.63 (0.49, 0.74) | 0.14 (0.14, 0.15) | NA | NA | NA |
| Tropical Latin America | -0.05 (-0.06, -0.05) | 0.29 (0.21, 0.36) | -0.00 (-0.00, -0.00) | 0.80 (0.74, 0.87) | 0.23 (0.22, 0.24) | NA | NA | NA |
| North Africa and Middle East | -0.04 (-0.04, -0.04) | 0.37 (0.28, 0.45) | -0.00 (-0.00, -0.00) | 0.47 (0.40, 0.56) | 0.28 (0.27, 0.29) | NA | NA | NA |
| South Asia | -0.05 (-0.06, -0.04) | 0.20 (0.07, 0.33) | -0.01 (-0.01, -0.01) | 0.51 (0.40, 0.63) | 0.75 (0.74, 0.76) | NA | NA | NA |
| Central Sub-Saharan Africa | 0.02 (0.01, 0.02) | 0.17 (0.13, 0.23) | -0.00 (-0.00, -0.00) | 0.31 (0.24, 0.40) | -0.14 (-0.15, -0.14) | NA | NA | NA |
| Eastern Sub-Saharan Africa | -0.10 (-0.11, -0.10) | 0.12 (0.09, 0.16) | 0.00 (-0.00, 0.00) | 0.38 (0.35, 0.41) | 0.27 (0.26, 0.27) | NA | NA | NA |
| Southern Sub-Saharan Africa | -0.03 (-0.04, -0.03) | 0.52 (0.45, 0.61) | 0.01 (0.01, 0.01) | 0.63 (0.57, 0.70) | 0.08 (0.08, 0.09) | NA | NA | NA |
| Western Sub-Saharan Africa | -0.01 (-0.01, -0.00) | 0.05 (0.00, 0.10) | 0.00 (0.00, 0.00) | 0.20 (0.14, 0.26) | 0.30 (0.29, 0.31) | NA | NA | NA |

Note: AAPC: Average Annual Percentage Change; ASPR: Age-Standardized Prevalence Rate; ASIR: Age-Standardized Incidence Rate; NA: Not Available.

Supplementary Table 7: Co-existence patterns of dementia with overall mental disorders among 204 Countries from 1990 to 2021

| Countries | 1990 | 1991 | 1992 | 1993 | 1994 | 1995 | 1996 | 1997 | 1998 | 1999 | 2000 | 2001 | 2002 | 2003 | 2004 | 2005 | 2006 | 2007 | 2008 | 2009 | 2010 | 2011 | 2012 | 2013 | 2014 | 2015 | 2016 | 2017 | 2018 | 2019 | 2020 | 2021 |
| --- | --- | --- | --- | --- | --- | --- | --- | --- | --- | --- | --- | --- | --- | --- | --- | --- | --- | --- | --- | --- | --- | --- | --- | --- | --- | --- | --- | --- | --- | --- | --- | --- |
| China | 4 | 4 | 4 | 4 | 4 | 4 | 4 | 4 | 4 | 4 | 4 | 4 | 4 | 4 | 4 | 4 | 4 | 4 | 4 | 4 | 4 | 4 | 4 | 4 | 4 | 4 | 4 | 4 | 4 | 4 | 4 | 4 |
| Democratic People's Republic of Korea | 4 | 4 | 4 | 4 | 4 | 4 | 4 | 4 | 4 | 4 | 4 | 4 | 4 | 4 | 4 | 4 | 4 | 4 | 4 | 4 | 4 | 4 | 4 | 4 | 4 | 4 | 4 | 4 | 4 | 4 | 4 | 4 |
| Taiwan | 3 | 3 | 3 | 3 | 3 | 3 | 3 | 3 | 3 | 3 | 3 | 3 | 3 | 3 | 3 | 3 | 3 | 3 | 3 | 3 | 3 | 3 | 3 | 3 | 3 | 3 | 3 | 3 | 3 | 3 | 3 | 3 |
| Cambodia | 4 | 4 | 4 | 4 | 4 | 4 | 4 | 4 | 4 | 4 | 4 | 4 | 4 | 4 | 4 | 4 | 4 | 4 | 4 | 4 | 4 | 4 | 4 | 4 | 4 | 4 | 4 | 4 | 4 | 4 | 4 | 4 |
| Indonesia | 4 | 4 | 4 | 4 | 4 | 4 | 4 | 4 | 4 | 4 | 4 | 4 | 4 | 4 | 4 | 4 | 4 | 4 | 4 | 4 | 4 | 4 | 4 | 4 | 4 | 4 | 4 | 4 | 4 | 4 | 4 | 4 |
| Lao People's Democratic Republic | 4 | 2 | 2 | 2 | 2 | 2 | 2 | 2 | 2 | 4 | 4 | 4 | 4 | 4 | 4 | 4 | 4 | 4 | 4 | 4 | 4 | 4 | 4 | 4 | 4 | 4 | 4 | 4 | 4 | 4 | 4 | 4 |
| Malaysia | 4 | 4 | 4 | 4 | 4 | 4 | 4 | 4 | 4 | 4 | 4 | 4 | 4 | 4 | 4 | 4 | 4 | 4 | 4 | 4 | 4 | 4 | 4 | 4 | 4 | 4 | 4 | 4 | 4 | 4 | 4 | 4 |
| Maldives | 4 | 4 | 4 | 4 | 4 | 4 | 4 | 4 | 4 | 4 | 4 | 4 | 4 | 4 | 4 | 4 | 4 | 4 | 4 | 4 | 4 | 4 | 4 | 4 | 4 | 4 | 4 | 4 | 4 | 4 | 4 | 4 |
| Myanmar | 4 | 4 | 4 | 4 | 4 | 4 | 4 | 4 | 4 | 4 | 4 | 4 | 4 | 4 | 4 | 4 | 4 | 4 | 4 | 4 | 4 | 4 | 4 | 4 | 4 | 4 | 4 | 4 | 4 | 4 | 4 | 4 |
| Philippines | 4 | 4 | 4 | 4 | 4 | 4 | 4 | 4 | 4 | 4 | 4 | 4 | 4 | 4 | 4 | 4 | 4 | 4 | 4 | 4 | 4 | 4 | 4 | 4 | 4 | 4 | 4 | 4 | 4 | 4 | 4 | 4 |
| Sri Lanka | 4 | 4 | 4 | 4 | 4 | 4 | 4 | 4 | 4 | 4 | 4 | 4 | 4 | 4 | 4 | 4 | 4 | 4 | 4 | 4 | 4 | 4 | 4 | 4 | 4 | 4 | 4 | 4 | 4 | 4 | 4 | 4 |
| Thailand | 3 | 3 | 3 | 3 | 3 | 3 | 3 | 3 | 3 | 3 | 3 | 3 | 3 | 3 | 4 | 4 | 4 | 4 | 4 | 4 | 4 | 4 | 4 | 4 | 4 | 4 | 4 | 4 | 4 | 4 | 4 | 4 |
| Timor-Leste | 4 | 4 | 4 | 4 | 4 | 4 | 4 | 4 | 4 | 4 | 4 | 4 | 4 | 4 | 4 | 4 | 4 | 4 | 4 | 4 | 4 | 4 | 4 | 4 | 4 | 4 | 4 | 4 | 4 | 4 | 4 | 4 |
| Viet Nam | 4 | 4 | 4 | 4 | 4 | 4 | 4 | 4 | 4 | 4 | 4 | 4 | 4 | 4 | 4 | 4 | 4 | 4 | 4 | 4 | 4 | 4 | 4 | 4 | 4 | 4 | 4 | 4 | 4 | 4 | 4 | 4 |
| Fiji | 4 | 4 | 4 | 4 | 4 | 4 | 4 | 4 | 4 | 4 | 4 | 4 | 4 | 4 | 4 | 4 | 4 | 4 | 4 | 4 | 4 | 4 | 4 | 4 | 4 | 4 | 4 | 4 | 4 | 4 | 4 | 4 |
| Kiribati | 4 | 4 | 4 | 4 | 4 | 4 | 4 | 4 | 4 | 4 | 4 | 4 | 4 | 4 | 4 | 4 | 4 | 4 | 4 | 4 | 4 | 4 | 4 | 4 | 4 | 4 | 4 | 4 | 4 | 4 | 4 | 4 |
| Marshall Islands | 3 | 3 | 4 | 4 | 3 | 3 | 3 | 3 | 4 | 3 | 3 | 3 | 3 | 3 | 3 | 3 | 3 | 3 | 3 | 3 | 3 | 3 | 3 | 3 | 3 | 3 | 3 | 3 | 3 | 3 | 4 | 3 |
| Micronesia (Federated States of) | 4 | 4 | 4 | 4 | 4 | 4 | 4 | 4 | 4 | 4 | 4 | 4 | 4 | 4 | 4 | 4 | 4 | 4 | 4 | 4 | 4 | 4 | 4 | 4 | 4 | 4 | 4 | 4 | 4 | 4 | 4 | 4 |
| Papua New Guinea | 4 | 4 | 4 | 4 | 4 | 4 | 4 | 4 | 4 | 4 | 4 | 4 | 4 | 4 | 4 | 4 | 4 | 4 | 4 | 4 | 4 | 4 | 4 | 4 | 4 | 4 | 4 | 4 | 4 | 4 | 4 | 4 |
| Samoa | 4 | 4 | 4 | 4 | 4 | 4 | 4 | 4 | 4 | 4 | 4 | 4 | 4 | 4 | 4 | 4 | 4 | 4 | 4 | 4 | 4 | 4 | 4 | 4 | 4 | 4 | 4 | 4 | 4 | 4 | 4 | 4 |
| Solomon Islands | 4 | 4 | 4 | 4 | 4 | 4 | 4 | 4 | 4 | 4 | 4 | 4 | 4 | 4 | 4 | 4 | 4 | 4 | 4 | 4 | 4 | 4 | 4 | 4 | 4 | 4 | 4 | 4 | 4 | 4 | 4 | 4 |
| Tonga | 4 | 4 | 4 | 4 | 4 | 4 | 4 | 4 | 4 | 4 | 4 | 4 | 4 | 4 | 4 | 4 | 4 | 4 | 4 | 4 | 4 | 4 | 4 | 4 | 4 | 4 | 4 | 4 | 4 | 4 | 4 | 4 |
| Vanuatu | 3 | 4 | 4 | 4 | 4 | 4 | 4 | 4 | 4 | 4 | 4 | 4 | 4 | 4 | 4 | 4 | 4 | 4 | 4 | 4 | 4 | 4 | 4 | 4 | 4 | 4 | 4 | 4 | 4 | 4 | 4 | 4 |
| Armenia | 4 | 4 | 4 | 4 | 4 | 4 | 4 | 4 | 4 | 4 | 4 | 4 | 4 | 4 | 4 | 4 | 4 | 4 | 4 | 4 | 4 | 4 | 4 | 4 | 4 | 4 | 4 | 4 | 4 | 4 | 4 | 4 |
| Azerbaijan | 4 | 4 | 4 | 4 | 4 | 4 | 4 | 4 | 4 | 4 | 4 | 4 | 4 | 4 | 4 | 4 | 4 | 4 | 4 | 4 | 4 | 4 | 4 | 4 | 4 | 4 | 4 | 4 | 4 | 4 | 4 | 4 |
| Georgia | 4 | 4 | 4 | 4 | 4 | 4 | 4 | 4 | 4 | 4 | 4 | 4 | 4 | 4 | 4 | 4 | 4 | 4 | 4 | 4 | 4 | 4 | 4 | 4 | 4 | 4 | 4 | 4 | 4 | 4 | 4 | 4 |
| Kazakhstan | 4 | 4 | 4 | 4 | 4 | 4 | 4 | 4 | 4 | 4 | 4 | 4 | 4 | 4 | 4 | 4 | 4 | 4 | 4 | 4 | 4 | 4 | 4 | 4 | 4 | 4 | 4 | 4 | 4 | 4 | 4 | 4 |
| Kyrgyzstan | 4 | 4 | 4 | 4 | 4 | 4 | 4 | 4 | 4 | 4 | 4 | 4 | 4 | 4 | 4 | 4 | 4 | 4 | 4 | 4 | 4 | 4 | 4 | 4 | 4 | 4 | 4 | 4 | 4 | 4 | 4 | 4 |
| Mongolia | 4 | 4 | 4 | 4 | 4 | 4 | 4 | 4 | 4 | 4 | 4 | 4 | 4 | 4 | 4 | 4 | 4 | 4 | 4 | 4 | 4 | 4 | 4 | 4 | 4 | 4 | 4 | 4 | 4 | 4 | 4 | 4 |
| Tajikistan | 4 | 4 | 4 | 4 | 4 | 4 | 4 | 4 | 4 | 4 | 4 | 4 | 4 | 4 | 4 | 4 | 4 | 4 | 4 | 4 | 4 | 4 | 4 | 4 | 4 | 4 | 4 | 4 | 4 | 4 | 4 | 4 |
| Turkmenistan | 4 | 4 | 4 | 4 | 4 | 4 | 4 | 4 | 4 | 4 | 4 | 4 | 4 | 4 | 4 | 4 | 4 | 4 | 4 | 4 | 4 | 4 | 4 | 4 | 4 | 4 | 4 | 4 | 4 | 4 | 4 | 4 |
| Uzbekistan | 3 | 3 | 3 | 3 | 3 | 3 | 3 | 4 | 4 | 4 | 4 | 4 | 4 | 4 | 3 | 3 | 3 | 3 | 4 | 4 | 4 | 4 | 4 | 4 | 4 | 4 | 4 | 4 | 4 | 4 | 4 | 4 |
| Albania | 4 | 4 | 4 | 4 | 4 | 4 | 4 | 4 | 4 | 4 | 4 | 4 | 4 | 4 | 4 | 4 | 4 | 4 | 4 | 4 | 4 | 4 | 4 | 4 | 4 | 4 | 4 | 4 | 4 | 4 | 4 | 4 |
| Bosnia and Herzegovina | 2 | 2 | 4 | 4 | 4 | 4 | 4 | 4 | 4 | 4 | 4 | 4 | 4 | 4 | 4 | 4 | 4 | 4 | 4 | 4 | 4 | 4 | 4 | 4 | 4 | 4 | 4 | 4 | 4 | 4 | 4 | 4 |
| Bulgaria | 4 | 4 | 4 | 4 | 4 | 4 | 4 | 4 | 4 | 4 | 4 | 4 | 4 | 4 | 4 | 4 | 4 | 4 | 4 | 4 | 4 | 4 | 4 | 4 | 4 | 4 | 4 | 4 | 4 | 4 | 4 | 2 |
| Croatia | 2 | 2 | 2 | 2 | 2 | 2 | 2 | 2 | 2 | 2 | 4 | 4 | 4 | 4 | 4 | 4 | 4 | 4 | 4 | 4 | 4 | 4 | 4 | 4 | 4 | 4 | 4 | 4 | 4 | 4 | 4 | 4 |
| Czechia | 4 | 4 | 4 | 4 | 4 | 4 | 4 | 4 | 4 | 4 | 4 | 4 | 4 | 4 | 4 | 4 | 4 | 4 | 4 | 4 | 4 | 4 | 4 | 4 | 4 | 4 | 4 | 4 | 4 | 4 | 4 | 4 |
| Hungary | 2 | 2 | 2 | 2 | 2 | 2 | 2 | 4 | 4 | 4 | 4 | 4 | 4 | 4 | 4 | 4 | 4 | 4 | 4 | 4 | 4 | 4 | 4 | 4 | 4 | 4 | 4 | 4 | 4 | 4 | 4 | 4 |
| North Macedonia | 4 | 4 | 4 | 4 | 4 | 4 | 4 | 4 | 4 | 4 | 4 | 4 | 4 | 4 | 4 | 4 | 4 | 4 | 4 | 4 | 4 | 4 | 4 | 4 | 4 | 4 | 4 | 4 | 4 | 4 | 4 | 4 |
| Montenegro | 4 | 4 | 4 | 4 | 4 | 4 | 4 | 4 | 4 | 4 | 4 | 4 | 4 | 4 | 4 | 4 | 4 | 4 | 4 | 4 | 4 | 4 | 4 | 4 | 4 | 4 | 4 | 4 | 4 | 4 | 4 | 2 |
| Poland | 4 | 4 | 4 | 4 | 4 | 4 | 4 | 4 | 4 | 4 | 4 | 4 | 4 | 4 | 4 | 4 | 4 | 4 | 4 | 4 | 4 | 4 | 4 | 4 | 4 | 4 | 4 | 4 | 4 | 4 | 4 | 4 |
| Romania | 4 | 4 | 4 | 4 | 4 | 4 | 4 | 4 | 4 | 4 | 4 | 4 | 4 | 4 | 4 | 4 | 4 | 4 | 4 | 4 | 4 | 4 | 4 | 4 | 4 | 4 | 4 | 4 | 4 | 4 | 4 | 4 |
| Serbia | 4 | 4 | 4 | 4 | 4 | 4 | 4 | 4 | 4 | 4 | 4 | 4 | 4 | 4 | 4 | 4 | 4 | 4 | 4 | 4 | 4 | 4 | 4 | 4 | 4 | 4 | 4 | 4 | 4 | 4 | 4 | 4 |
| Slovakia | 4 | 4 | 4 | 4 | 4 | 4 | 4 | 4 | 4 | 4 | 4 | 4 | 4 | 4 | 4 | 4 | 4 | 4 | 4 | 4 | 4 | 4 | 4 | 4 | 4 | 4 | 4 | 4 | 4 | 4 | 4 | 4 |
| Slovenia | 2 | 2 | 2 | 2 | 2 | 2 | 2 | 2 | 2 | 2 | 2 | 2 | 2 | 2 | 2 | 2 | 2 | 2 | 2 | 2 | 2 | 2 | 2 | 4 | 4 | 4 | 4 | 4 | 4 | 4 | 4 | 4 |
| Belarus | 2 | 2 | 2 | 2 | 2 | 2 | 2 | 2 | 2 | 2 | 2 | 2 | 2 | 2 | 2 | 2 | 2 | 2 | 2 | 2 | 4 | 4 | 4 | 4 | 4 | 4 | 4 | 4 | 4 | 4 | 4 | 1 |
| Estonia | 5 | 5 | 5 | 5 | 5 | 5 | 5 | 5 | 5 | 5 | 5 | 5 | 1 | 1 | 1 | 1 | 4 | 4 | 4 | 4 | 4 | 4 | 4 | 4 | 4 | 4 | 2 | 2 | 2 | 2 | 2 | 2 |
| Latvia | 5 | 5 | 5 | 5 | 5 | 5 | 5 | 5 | 5 | 5 | 5 | 5 | 2 | 2 | 2 | 2 | 4 | 4 | 4 | 4 | 4 | 4 | 4 | 4 | 4 | 4 | 4 | 4 | 4 | 4 | 4 | 1 |
| Lithuania | 5 | 5 | 5 | 5 | 5 | 5 | 5 | 5 | 5 | 5 | 5 | 5 | 5 | 5 | 5 | 5 | 5 | 5 | 5 | 5 | 5 | 5 | 5 | 5 | 5 | 5 | 5 | 5 | 5 | 5 | 5 | 5 |
| Republic of Moldova | 2 | 2 | 2 | 2 | 2 | 2 | 2 | 2 | 2 | 2 | 2 | 2 | 2 | 2 | 2 | 2 | 2 | 2 | 2 | 2 | 2 | 2 | 2 | 2 | 2 | 2 | 2 | 2 | 2 | 2 | 2 | 2 |
| Russian Federation | 4 | 4 | 4 | 4 | 4 | 4 | 4 | 4 | 4 | 4 | 4 | 4 | 4 | 4 | 4 | 4 | 4 | 4 | 4 | 4 | 4 | 4 | 4 | 4 | 4 | 4 | 4 | 4 | 4 | 4 | 4 | 4 |
| Ukraine | 2 | 4 | 2 | 1 | 1 | 1 | 1 | 1 | 1 | 1 | 4 | 4 | 4 | 4 | 4 | 4 | 4 | 4 | 4 | 4 | 4 | 4 | 4 | 4 | 4 | 4 | 4 | 4 | 4 | 2 | 4 | 4 |
| Brunei Darussalam | 3 | 3 | 3 | 3 | 3 | 3 | 3 | 3 | 3 | 3 | 3 | 3 | 3 | 3 | 3 | 3 | 3 | 3 | 3 | 3 | 3 | 3 | 3 | 3 | 3 | 3 | 3 | 3 | 3 | 3 | 3 | 3 |
| Japan | 4 | 4 | 4 | 4 | 4 | 4 | 4 | 4 | 4 | 4 | 4 | 4 | 4 | 4 | 4 | 4 | 4 | 4 | 4 | 4 | 4 | 4 | 4 | 4 | 4 | 4 | 4 | 4 | 4 | 4 | 4 | 4 |
| Republic of Korea | 4 | 4 | 4 | 4 | 4 | 4 | 4 | 4 | 4 | 4 | 4 | 4 | 4 | 4 | 4 | 4 | 4 | 4 | 4 | 4 | 4 | 4 | 4 | 4 | 4 | 4 | 4 | 4 | 4 | 4 | 4 | 4 |
| Singapore | 3 | 3 | 3 | 3 | 3 | 3 | 3 | 3 | 3 | 3 | 3 | 3 | 3 | 3 | 3 | 3 | 3 | 3 | 3 | 3 | 3 | 3 | 3 | 3 | 3 | 3 | 3 | 3 | 3 | 3 | 3 | 3 |
| Australia | 1 | 1 | 1 | 1 | 1 | 1 | 1 | 1 | 5 | 5 | 5 | 5 | 5 | 5 | 5 | 5 | 5 | 5 | 5 | 5 | 5 | 5 | 5 | 5 | 5 | 5 | 5 | 5 | 5 | 5 | 5 | 5 |
| New Zealand | 1 | 1 | 1 | 1 | 1 | 1 | 1 | 1 | 1 | 1 | 1 | 1 | 1 | 1 | 1 | 1 | 1 | 1 | 1 | 1 | 1 | 1 | 1 | 1 | 1 | 1 | 1 | 1 | 1 | 1 | 1 | 1 |
| Andorra | 4 | 4 | 4 | 4 | 4 | 4 | 4 | 4 | 4 | 4 | 4 | 4 | 4 | 4 | 4 | 4 | 4 | 4 | 4 | 4 | 4 | 4 | 4 | 4 | 4 | 4 | 2 | 2 | 2 | 2 | 5 | 2 |
| Austria | 4 | 4 | 4 | 4 | 4 | 4 | 4 | 4 | 4 | 4 | 4 | 4 | 4 | 4 | 4 | 4 | 4 | 4 | 4 | 4 | 4 | 4 | 4 | 4 | 4 | 4 | 4 | 4 | 4 | 4 | 4 | 4 |
| Belgium | 4 | 4 | 4 | 4 | 4 | 4 | 4 | 4 | 4 | 4 | 4 | 4 | 4 | 4 | 4 | 4 | 4 | 4 | 4 | 4 | 4 | 4 | 4 | 4 | 4 | 4 | 4 | 4 | 4 | 4 | 4 | 4 |
| Cyprus | 4 | 4 | 4 | 4 | 4 | 4 | 4 | 4 | 4 | 4 | 4 | 4 | 4 | 4 | 4 | 4 | 4 | 4 | 4 | 4 | 4 | 4 | 4 | 4 | 4 | 4 | 4 | 4 | 4 | 4 | 4 | 4 |
| Denmark | 5 | 5 | 5 | 5 | 5 | 5 | 5 | 5 | 5 | 5 | 5 | 5 | 5 | 5 | 5 | 5 | 5 | 5 | 5 | 5 | 5 | 5 | 5 | 5 | 5 | 5 | 5 | 5 | 5 | 5 | 5 | 5 |
| Finland | 4 | 4 | 4 | 4 | 4 | 4 | 4 | 4 | 4 | 4 | 4 | 4 | 2 | 2 | 2 | 4 | 2 | 2 | 2 | 2 | 2 | 2 | 2 | 2 | 2 | 2 | 2 | 2 | 2 | 2 | 2 | 2 |
| France | 5 | 5 | 5 | 5 | 5 | 5 | 5 | 5 | 5 | 5 | 5 | 5 | 5 | 5 | 5 | 5 | 5 | 5 | 5 | 5 | 5 | 5 | 5 | 5 | 5 | 5 | 5 | 5 | 5 | 5 | 5 | 5 |
| Germany | 4 | 4 | 4 | 4 | 4 | 4 | 4 | 4 | 4 | 4 | 4 | 4 | 4 | 4 | 4 | 4 | 4 | 4 | 4 | 1 | 1 | 1 | 1 | 1 | 1 | 1 | 1 | 1 | 1 | 1 | 1 | 1 |
| Greece | 1 | 1 | 1 | 1 | 1 | 1 | 1 | 1 | 1 | 1 | 1 | 1 | 1 | 1 | 1 | 1 | 1 | 1 | 1 | 1 | 1 | 1 | 1 | 1 | 1 | 1 | 1 | 1 | 1 | 1 | 1 | 1 |
| Iceland | 4 | 4 | 4 | 4 | 4 | 4 | 4 | 4 | 4 | 4 | 4 | 4 | 4 | 4 | 4 | 4 | 4 | 4 | 4 | 4 | 4 | 4 | 4 | 4 | 4 | 4 | 4 | 4 | 4 | 4 | 4 | 4 |
| Ireland | 1 | 1 | 1 | 1 | 1 | 1 | 1 | 1 | 1 | 1 | 1 | 1 | 1 | 1 | 1 | 1 | 1 | 1 | 1 | 1 | 1 | 1 | 5 | 5 | 5 | 5 | 5 | 5 | 5 | 5 | 5 | 5 |
| Israel | 4 | 4 | 4 | 4 | 2 | 2 | 2 | 2 | 2 | 2 | 2 | 2 | 2 | 2 | 2 | 2 | 2 | 2 | 2 | 2 | 2 | 2 | 2 | 2 | 2 | 2 | 2 | 2 | 2 | 2 | 2 | 2 |
| Italy | 1 | 1 | 1 | 1 | 1 | 1 | 1 | 1 | 1 | 1 | 1 | 1 | 1 | 1 | 4 | 4 | 4 | 1 | 1 | 1 | 1 | 1 | 1 | 1 | 1 | 1 | 1 | 1 | 1 | 1 | 1 | 1 |
| Luxembourg | 5 | 5 | 5 | 5 | 5 | 5 | 5 | 5 | 5 | 5 | 5 | 5 | 5 | 5 | 5 | 5 | 5 | 5 | 5 | 5 | 5 | 5 | 5 | 5 | 5 | 5 | 5 | 5 | 5 | 5 | 5 | 5 |
| Malta | 4 | 4 | 4 | 4 | 4 | 4 | 4 | 4 | 4 | 4 | 2 | 2 | 2 | 2 | 2 | 2 | 2 | 2 | 2 | 2 | 2 | 2 | 2 | 2 | 2 | 2 | 2 | 2 | 2 | 2 | 2 | 2 |
| Netherlands | 1 | 1 | 1 | 1 | 1 | 1 | 1 | 1 | 1 | 1 | 1 | 1 | 1 | 1 | 1 | 1 | 1 | 1 | 1 | 1 | 1 | 1 | 1 | 1 | 1 | 1 | 1 | 1 | 1 | 1 | 1 | 1 |
| Norway | 1 | 1 | 1 | 1 | 1 | 1 | 1 | 1 | 1 | 1 | 1 | 1 | 1 | 1 | 1 | 1 | 1 | 1 | 1 | 1 | 1 | 1 | 5 | 5 | 5 | 5 | 5 | 5 | 5 | 5 | 5 | 5 |
| Portugal | 5 | 1 | 1 | 5 | 1 | 1 | 1 | 1 | 1 | 1 | 1 | 1 | 1 | 1 | 1 | 1 | 1 | 1 | 1 | 1 | 1 | 1 | 1 | 1 | 1 | 1 | 1 | 1 | 1 | 1 | 1 | 1 |
| Spain | 1 | 5 | 5 | 5 | 5 | 5 | 5 | 5 | 5 | 5 | 5 | 5 | 5 | 5 | 5 | 5 | 5 | 5 | 5 | 5 | 5 | 5 | 5 | 5 | 5 | 5 | 5 | 5 | 5 | 5 | 5 | 5 |
| Sweden | 1 | 1 | 1 | 1 | 1 | 1 | 1 | 1 | 1 | 1 | 1 | 1 | 1 | 1 | 1 | 1 | 1 | 1 | 1 | 1 | 1 | 1 | 1 | 1 | 1 | 1 | 1 | 1 | 1 | 1 | 1 | 1 |
| Switzerland | 1 | 1 | 1 | 1 | 1 | 1 | 1 | 1 | 1 | 1 | 1 | 1 | 1 | 1 | 1 | 1 | 1 | 1 | 1 | 1 | 1 | 1 | 1 | 1 | 1 | 5 | 5 | 5 | 5 | 5 | 5 | 5 |
| United Kingdom | 2 | 2 | 2 | 2 | 2 | 2 | 2 | 5 | 5 | 5 | 5 | 5 | 2 | 2 | 2 | 2 | 2 | 2 | 2 | 2 | 2 | 2 | 2 | 2 | 2 | 2 | 2 | 5 | 5 | 5 | 5 | 5 |
| Argentina | 4 | 4 | 4 | 4 | 4 | 4 | 4 | 4 | 4 | 4 | 4 | 4 | 4 | 4 | 4 | 4 | 4 | 4 | 3 | 3 | 3 | 3 | 4 | 4 | 4 | 3 | 3 | 3 | 3 | 3 | 3 | 3 |
| Chile | 5 | 5 | 5 | 5 | 5 | 5 | 5 | 5 | 5 | 5 | 5 | 5 | 5 | 5 | 5 | 5 | 5 | 5 | 5 | 2 | 2 | 2 | 2 | 2 | 2 | 2 | 2 | 2 | 2 | 2 | 5 | 2 |
| Uruguay | 4 | 4 | 4 | 4 | 4 | 4 | 4 | 4 | 4 | 2 | 2 | 2 | 2 | 2 | 2 | 2 | 2 | 2 | 2 | 5 | 5 | 5 | 5 | 5 | 5 | 2 | 2 | 2 | 2 | 2 | 2 | 2 |
| Canada | 4 | 4 | 4 | 4 | 4 | 4 | 4 | 4 | 4 | 4 | 4 | 4 | 4 | 4 | 4 | 4 | 4 | 4 | 4 | 4 | 4 | 4 | 4 | 4 | 4 | 4 | 4 | 4 | 4 | 4 | 4 | 4 |
| United States of America | 1 | 1 | 1 | 1 | 1 | 1 | 1 | 1 | 1 | 1 | 1 | 1 | 1 | 1 | 1 | 1 | 1 | 1 | 1 | 1 | 1 | 1 | 1 | 1 | 1 | 1 | 1 | 1 | 1 | 1 | 1 | 1 |
| Antigua and Barbuda | 5 | 5 | 5 | 5 | 5 | 5 | 5 | 5 | 5 | 5 | 5 | 5 | 5 | 5 | 5 | 5 | 5 | 5 | 5 | 5 | 5 | 5 | 5 | 5 | 5 | 5 | 5 | 5 | 5 | 5 | 5 | 5 |
| Bahamas | 5 | 5 | 5 | 5 | 5 | 5 | 5 | 5 | 5 | 5 | 5 | 5 | 5 | 5 | 5 | 5 | 5 | 5 | 5 | 5 | 5 | 5 | 5 | 5 | 5 | 5 | 5 | 5 | 5 | 5 | 5 | 5 |
| Barbados | 5 | 5 | 5 | 5 | 5 | 5 | 5 | 5 | 5 | 5 | 5 | 5 | 5 | 5 | 5 | 5 | 5 | 5 | 5 | 5 | 5 | 5 | 5 | 5 | 5 | 5 | 5 | 5 | 5 | 5 | 5 | 5 |
| Belize | 5 | 5 | 5 | 5 | 5 | 5 | 5 | 5 | 5 | 5 | 5 | 5 | 5 | 5 | 5 | 5 | 5 | 5 | 5 | 5 | 5 | 5 | 5 | 5 | 5 | 5 | 5 | 5 | 5 | 5 | 5 | 5 |
| Cuba | 5 | 5 | 5 | 5 | 5 | 5 | 5 | 5 | 5 | 5 | 5 | 5 | 5 | 5 | 5 | 5 | 5 | 5 | 5 | 5 | 5 | 5 | 5 | 5 | 5 | 5 | 5 | 5 | 5 | 5 | 5 | 5 |
| Dominica | 5 | 5 | 5 | 5 | 5 | 5 | 5 | 5 | 5 | 5 | 5 | 5 | 5 | 5 | 5 | 5 | 5 | 5 | 5 | 5 | 5 | 5 | 5 | 5 | 5 | 5 | 5 | 5 | 5 | 5 | 5 | 5 |
| Dominican Republic | 5 | 5 | 5 | 5 | 5 | 5 | 5 | 5 | 5 | 5 | 5 | 5 | 5 | 5 | 5 | 5 | 5 | 5 | 5 | 5 | 5 | 5 | 5 | 5 | 5 | 5 | 5 | 5 | 5 | 5 | 5 | 5 |
| Grenada | 5 | 5 | 5 | 5 | 5 | 5 | 5 | 5 | 5 | 5 | 5 | 5 | 5 | 5 | 5 | 5 | 5 | 5 | 5 | 5 | 5 | 5 | 5 | 5 | 5 | 5 | 5 | 5 | 5 | 5 | 5 | 5 |
| Guyana | 5 | 5 | 5 | 5 | 5 | 5 | 5 | 5 | 5 | 5 | 5 | 5 | 5 | 5 | 5 | 5 | 5 | 5 | 5 | 5 | 5 | 5 | 5 | 5 | 5 | 5 | 5 | 5 | 5 | 5 | 5 | 5 |
| Haiti | 5 | 5 | 5 | 5 | 5 | 5 | 5 | 5 | 5 | 5 | 5 | 5 | 5 | 5 | 5 | 5 | 5 | 5 | 5 | 5 | 5 | 5 | 5 | 5 | 5 | 5 | 5 | 5 | 5 | 5 | 5 | 5 |
| Jamaica | 2 | 2 | 2 | 2 | 2 | 2 | 2 | 2 | 2 | 2 | 2 | 2 | 2 | 2 | 2 | 2 | 2 | 2 | 2 | 2 | 2 | 2 | 2 | 2 | 2 | 2 | 2 | 2 | 2 | 2 | 2 | 2 |
| Saint Lucia | 5 | 5 | 5 | 5 | 5 | 5 | 5 | 5 | 5 | 5 | 5 | 5 | 5 | 5 | 5 | 5 | 5 | 5 | 5 | 5 | 5 | 5 | 5 | 5 | 5 | 5 | 5 | 5 | 5 | 5 | 5 | 5 |
| Saint Vincent and the Grenadines | 5 | 5 | 5 | 5 | 5 | 5 | 5 | 5 | 5 | 5 | 5 | 5 | 5 | 5 | 5 | 5 | 5 | 5 | 5 | 5 | 5 | 5 | 5 | 5 | 5 | 5 | 5 | 5 | 5 | 5 | 5 | 5 |
| Suriname | 5 | 5 | 5 | 5 | 5 | 5 | 5 | 5 | 5 | 5 | 5 | 5 | 5 | 5 | 5 | 5 | 5 | 5 | 5 | 5 | 5 | 5 | 5 | 5 | 5 | 5 | 5 | 5 | 5 | 5 | 5 | 5 |
| Trinidad and Tobago | 5 | 5 | 5 | 5 | 5 | 5 | 5 | 5 | 5 | 5 | 5 | 5 | 5 | 5 | 5 | 5 | 5 | 5 | 5 | 5 | 5 | 5 | 5 | 5 | 5 | 5 | 5 | 5 | 5 | 5 | 5 | 5 |
| Bolivia (Plurinational State of) | 5 | 5 | 5 | 5 | 5 | 5 | 5 | 5 | 5 | 5 | 5 | 5 | 5 | 5 | 5 | 5 | 5 | 5 | 5 | 5 | 5 | 5 | 5 | 5 | 5 | 5 | 5 | 5 | 5 | 5 | 5 | 5 |
| Ecuador | 5 | 5 | 5 | 5 | 5 | 5 | 5 | 5 | 5 | 5 | 5 | 5 | 5 | 5 | 5 | 5 | 5 | 5 | 5 | 5 | 5 | 5 | 5 | 5 | 5 | 5 | 5 | 5 | 5 | 5 | 5 | 5 |
| Peru | 5 | 5 | 5 | 5 | 5 | 5 | 5 | 5 | 5 | 5 | 5 | 5 | 5 | 5 | 5 | 5 | 5 | 5 | 5 | 5 | 5 | 5 | 5 | 5 | 5 | 5 | 5 | 5 | 5 | 5 | 5 | 5 |
| Colombia | 4 | 4 | 4 | 4 | 4 | 4 | 4 | 4 | 4 | 4 | 4 | 4 | 4 | 4 | 4 | 2 | 4 | 4 | 4 | 4 | 4 | 4 | 4 | 4 | 4 | 4 | 4 | 4 | 4 | 4 | 4 | 4 |
| Costa Rica | 4 | 4 | 4 | 4 | 4 | 4 | 4 | 4 | 4 | 4 | 2 | 2 | 2 | 2 | 2 | 2 | 2 | 2 | 2 | 2 | 2 | 2 | 2 | 2 | 2 | 2 | 2 | 2 | 2 | 2 | 2 | 2 |
| El Salvador | 2 | 2 | 2 | 2 | 2 | 2 | 2 | 2 | 2 | 2 | 2 | 2 | 2 | 2 | 2 | 2 | 2 | 2 | 2 | 2 | 2 | 2 | 2 | 2 | 2 | 2 | 2 | 2 | 2 | 2 | 2 | 2 |
| Guatemala | 2 | 2 | 2 | 2 | 2 | 2 | 2 | 5 | 5 | 5 | 5 | 5 | 5 | 5 | 5 | 5 | 5 | 5 | 5 | 5 | 5 | 5 | 5 | 2 | 2 | 2 | 2 | 2 | 2 | 2 | 2 | 2 |
| Honduras | 4 | 4 | 4 | 4 | 4 | 4 | 4 | 2 | 2 | 2 | 2 | 2 | 2 | 2 | 2 | 2 | 2 | 2 | 2 | 2 | 2 | 2 | 2 | 2 | 2 | 2 | 2 | 2 | 2 | 2 | 2 | 2 |
| Mexico | 3 | 3 | 3 | 3 | 3 | 3 | 3 | 3 | 3 | 3 | 3 | 3 | 3 | 3 | 3 | 3 | 3 | 3 | 5 | 5 | 5 | 5 | 5 | 5 | 5 | 5 | 5 | 5 | 5 | 5 | 5 | 5 |
| Nicaragua | 4 | 4 | 2 | 2 | 2 | 2 | 2 | 2 | 2 | 2 | 2 | 2 | 2 | 2 | 2 | 2 | 2 | 2 | 2 | 2 | 2 | 2 | 2 | 2 | 2 | 2 | 4 | 4 | 4 | 4 | 4 | 4 |
| Panama | 4 | 4 | 4 | 4 | 4 | 4 | 4 | 4 | 4 | 4 | 4 | 4 | 4 | 4 | 4 | 4 | 4 | 4 | 4 | 4 | 4 | 4 | 4 | 4 | 4 | 4 | 4 | 4 | 4 | 4 | 2 | 4 |
| Venezuela (Bolivarian Republic of) | 4 | 4 | 4 | 4 | 4 | 4 | 4 | 4 | 4 | 4 | 4 | 4 | 4 | 4 | 4 | 4 | 4 | 4 | 4 | 4 | 4 | 4 | 4 | 4 | 4 | 4 | 4 | 4 | 4 | 4 | 4 | 4 |
| Brazil | 1 | 1 | 1 | 1 | 1 | 1 | 1 | 1 | 1 | 1 | 1 | 1 | 1 | 1 | 1 | 1 | 1 | 1 | 1 | 1 | 1 | 1 | 1 | 1 | 1 | 1 | 1 | 1 | 1 | 1 | 1 | 1 |
| Paraguay | 1 | 1 | 1 | 1 | 1 | 1 | 1 | 1 | 1 | 1 | 1 | 1 | 1 | 1 | 1 | 1 | 1 | 1 | 1 | 1 | 1 | 1 | 1 | 1 | 1 | 1 | 1 | 1 | 1 | 1 | 1 | 1 |
| Algeria | 1 | 1 | 1 | 1 | 1 | 1 | 1 | 1 | 1 | 1 | 1 | 1 | 1 | 1 | 1 | 1 | 1 | 1 | 1 | 4 | 4 | 4 | 4 | 1 | 1 | 1 | 1 | 1 | 1 | 1 | 4 | 4 |
| Bahrain | 1 | 1 | 1 | 1 | 1 | 1 | 1 | 1 | 1 | 1 | 1 | 1 | 1 | 1 | 1 | 1 | 1 | 1 | 1 | 1 | 1 | 1 | 1 | 1 | 1 | 1 | 1 | 1 | 1 | 1 | 1 | 1 |
| Egypt | 4 | 4 | 4 | 4 | 4 | 4 | 4 | 4 | 4 | 4 | 4 | 4 | 4 | 4 | 4 | 4 | 4 | 4 | 4 | 4 | 4 | 4 | 4 | 4 | 4 | 4 | 4 | 4 | 4 | 4 | 4 | 4 |
| Iran (Islamic Republic of) | 1 | 1 | 1 | 1 | 1 | 1 | 1 | 1 | 1 | 1 | 1 | 1 | 1 | 1 | 1 | 1 | 1 | 1 | 1 | 1 | 1 | 1 | 1 | 1 | 1 | 1 | 1 | 1 | 1 | 1 | 1 | 1 |
| Iraq | 4 | 4 | 4 | 4 | 4 | 4 | 4 | 4 | 4 | 4 | 4 | 4 | 4 | 4 | 4 | 4 | 4 | 4 | 4 | 4 | 4 | 4 | 4 | 4 | 4 | 4 | 4 | 4 | 4 | 4 | 1 | 4 |
| Jordan | 1 | 1 | 1 | 1 | 1 | 1 | 1 | 1 | 1 | 1 | 1 | 1 | 1 | 1 | 1 | 1 | 1 | 1 | 1 | 1 | 1 | 1 | 1 | 1 | 1 | 1 | 1 | 1 | 1 | 1 | 4 | 4 |
| Kuwait | 4 | 4 | 4 | 4 | 4 | 4 | 4 | 4 | 4 | 4 | 4 | 4 | 4 | 4 | 4 | 4 | 4 | 4 | 4 | 4 | 4 | 4 | 4 | 4 | 4 | 4 | 4 | 4 | 4 | 4 | 4 | 4 |
| Lebanon | 1 | 1 | 1 | 1 | 1 | 1 | 1 | 1 | 1 | 1 | 1 | 1 | 1 | 1 | 1 | 1 | 1 | 1 | 1 | 1 | 1 | 1 | 1 | 1 | 1 | 1 | 1 | 1 | 1 | 1 | 1 | 1 |
| Libya | 1 | 1 | 1 | 1 | 1 | 1 | 1 | 1 | 1 | 1 | 1 | 1 | 1 | 1 | 1 | 1 | 1 | 1 | 1 | 1 | 1 | 1 | 1 | 1 | 1 | 1 | 1 | 1 | 1 | 1 | 1 | 1 |
| Morocco | 1 | 1 | 1 | 1 | 1 | 1 | 1 | 1 | 1 | 1 | 1 | 1 | 1 | 1 | 1 | 1 | 1 | 1 | 1 | 1 | 1 | 1 | 1 | 1 | 1 | 1 | 1 | 1 | 1 | 1 | 1 | 1 |
| Palestine | 1 | 1 | 1 | 1 | 1 | 1 | 1 | 1 | 1 | 1 | 1 | 1 | 1 | 1 | 1 | 1 | 1 | 1 | 1 | 1 | 1 | 1 | 1 | 1 | 1 | 1 | 1 | 1 | 1 | 1 | 1 | 1 |
| Oman | 4 | 4 | 4 | 4 | 4 | 4 | 4 | 4 | 4 | 4 | 4 | 4 | 4 | 4 | 4 | 4 | 4 | 4 | 4 | 4 | 4 | 4 | 4 | 4 | 4 | 4 | 4 | 4 | 4 | 4 | 4 | 4 |
| Qatar | 4 | 4 | 4 | 4 | 4 | 4 | 4 | 4 | 4 | 4 | 4 | 4 | 4 | 4 | 4 | 4 | 4 | 4 | 4 | 4 | 4 | 4 | 4 | 4 | 4 | 4 | 4 | 4 | 4 | 4 | 4 | 4 |
| Saudi Arabia | 4 | 4 | 4 | 4 | 4 | 4 | 4 | 4 | 4 | 4 | 4 | 4 | 4 | 4 | 4 | 4 | 4 | 4 | 4 | 4 | 4 | 4 | 4 | 4 | 4 | 4 | 4 | 4 | 4 | 4 | 4 | 4 |
| Syrian Arab Republic | 1 | 1 | 1 | 1 | 1 | 1 | 1 | 1 | 1 | 1 | 1 | 1 | 1 | 1 | 1 | 1 | 1 | 1 | 1 | 1 | 1 | 1 | 1 | 1 | 1 | 1 | 1 | 1 | 1 | 1 | 1 | 1 |
| Tunisia | 1 | 1 | 1 | 1 | 1 | 1 | 1 | 1 | 1 | 1 | 1 | 1 | 1 | 1 | 1 | 1 | 1 | 1 | 1 | 1 | 1 | 1 | 1 | 1 | 1 | 1 | 1 | 1 | 1 | 1 | 1 | 1 |
| Türkiye | 4 | 4 | 4 | 4 | 4 | 4 | 4 | 4 | 4 | 4 | 4 | 4 | 4 | 4 | 4 | 4 | 4 | 4 | 4 | 4 | 4 | 4 | 4 | 4 | 4 | 4 | 4 | 4 | 4 | 4 | 4 | 4 |
| United Arab Emirates | 4 | 4 | 4 | 4 | 4 | 4 | 4 | 4 | 4 | 4 | 4 | 4 | 4 | 4 | 4 | 4 | 4 | 4 | 4 | 4 | 4 | 4 | 4 | 4 | 4 | 4 | 4 | 4 | 4 | 4 | 4 | 4 |
| Yemen | 1 | 1 | 1 | 1 | 1 | 1 | 1 | 1 | 1 | 1 | 1 | 1 | 1 | 1 | 1 | 1 | 1 | 1 | 1 | 1 | 1 | 1 | 1 | 1 | 1 | 1 | 1 | 1 | 1 | 1 | 1 | 1 |
| Afghanistan | 1 | 1 | 1 | 1 | 1 | 1 | 1 | 1 | 1 | 1 | 1 | 1 | 1 | 1 | 1 | 1 | 1 | 1 | 1 | 1 | 1 | 1 | 1 | 1 | 1 | 1 | 1 | 1 | 1 | 1 | 1 | 1 |
| Bangladesh | 5 | 5 | 5 | 5 | 5 | 5 | 5 | 5 | 5 | 5 | 5 | 5 | 5 | 5 | 5 | 5 | 5 | 5 | 5 | 5 | 5 | 5 | 5 | 5 | 5 | 5 | 5 | 5 | 5 | 5 | 5 | 5 |
| Bhutan | 5 | 5 | 5 | 5 | 5 | 5 | 5 | 5 | 5 | 5 | 5 | 5 | 5 | 5 | 5 | 5 | 5 | 5 | 5 | 5 | 5 | 5 | 5 | 5 | 5 | 5 | 5 | 5 | 5 | 5 | 5 | 5 |
| India | 5 | 5 | 5 | 5 | 5 | 5 | 5 | 5 | 5 | 5 | 5 | 5 | 5 | 5 | 5 | 5 | 5 | 5 | 5 | 5 | 5 | 5 | 5 | 5 | 5 | 5 | 5 | 5 | 5 | 5 | 5 | 5 |
| Nepal | 5 | 5 | 5 | 5 | 5 | 5 | 5 | 5 | 5 | 5 | 5 | 5 | 5 | 5 | 5 | 5 | 5 | 5 | 5 | 5 | 5 | 5 | 5 | 5 | 5 | 5 | 5 | 5 | 5 | 5 | 5 | 5 |
| Pakistan | 5 | 5 | 5 | 5 | 5 | 5 | 5 | 5 | 5 | 5 | 5 | 5 | 5 | 5 | 5 | 5 | 5 | 5 | 5 | 5 | 5 | 5 | 5 | 5 | 5 | 5 | 5 | 5 | 5 | 5 | 5 | 5 |
| Angola | 1 | 1 | 1 | 1 | 1 | 1 | 1 | 1 | 1 | 1 | 1 | 1 | 1 | 1 | 1 | 1 | 1 | 1 | 1 | 1 | 1 | 1 | 1 | 1 | 1 | 1 | 1 | 1 | 1 | 1 | 1 | 1 |
| Central African Republic | 1 | 1 | 1 | 1 | 1 | 1 | 1 | 1 | 1 | 1 | 1 | 1 | 1 | 1 | 1 | 1 | 1 | 1 | 1 | 1 | 1 | 1 | 1 | 1 | 1 | 1 | 1 | 1 | 1 | 1 | 1 | 1 |
| Congo | 1 | 1 | 1 | 1 | 1 | 1 | 1 | 1 | 1 | 1 | 1 | 1 | 1 | 1 | 1 | 1 | 1 | 1 | 1 | 1 | 1 | 1 | 1 | 1 | 1 | 1 | 1 | 1 | 1 | 1 | 1 | 1 |
| Democratic Republic of the Congo | 1 | 1 | 1 | 1 | 1 | 1 | 1 | 1 | 1 | 1 | 1 | 1 | 1 | 1 | 1 | 1 | 1 | 1 | 1 | 1 | 1 | 1 | 1 | 1 | 1 | 1 | 1 | 1 | 1 | 1 | 1 | 1 |
| Equatorial Guinea | 1 | 1 | 1 | 1 | 1 | 1 | 1 | 1 | 1 | 1 | 1 | 1 | 1 | 1 | 1 | 1 | 1 | 1 | 1 | 1 | 1 | 1 | 1 | 1 | 1 | 1 | 1 | 1 | 1 | 1 | 1 | 1 |
| Gabon | 1 | 1 | 1 | 1 | 1 | 1 | 1 | 1 | 1 | 1 | 1 | 1 | 1 | 1 | 1 | 1 | 1 | 1 | 1 | 1 | 1 | 1 | 1 | 1 | 1 | 1 | 1 | 1 | 1 | 1 | 1 | 1 |
| Burundi | 5 | 5 | 5 | 5 | 5 | 5 | 5 | 5 | 5 | 5 | 5 | 5 | 5 | 5 | 5 | 5 | 5 | 5 | 5 | 5 | 5 | 5 | 5 | 5 | 5 | 5 | 5 | 5 | 5 | 5 | 5 | 5 |
| Comoros | 5 | 5 | 5 | 5 | 5 | 5 | 5 | 5 | 5 | 5 | 5 | 5 | 5 | 5 | 5 | 5 | 5 | 5 | 5 | 5 | 5 | 5 | 5 | 5 | 5 | 5 | 5 | 5 | 5 | 5 | 5 | 5 |
| Djibouti | 2 | 2 | 2 | 2 | 2 | 2 | 2 | 2 | 2 | 2 | 2 | 2 | 2 | 2 | 5 | 2 | 2 | 2 | 2 | 2 | 2 | 2 | 2 | 2 | 2 | 2 | 2 | 2 | 2 | 2 | 2 | 2 |
| Eritrea | 5 | 5 | 5 | 5 | 5 | 5 | 5 | 5 | 5 | 5 | 5 | 5 | 5 | 5 | 5 | 5 | 5 | 5 | 5 | 5 | 5 | 5 | 5 | 5 | 5 | 5 | 5 | 5 | 5 | 5 | 5 | 5 |
| Ethiopia | 5 | 5 | 5 | 5 | 5 | 5 | 5 | 5 | 5 | 5 | 5 | 5 | 5 | 5 | 5 | 5 | 5 | 5 | 5 | 5 | 5 | 5 | 5 | 5 | 5 | 5 | 5 | 5 | 5 | 5 | 5 | 5 |
| Kenya | 5 | 5 | 5 | 5 | 5 | 5 | 5 | 5 | 5 | 5 | 5 | 5 | 5 | 5 | 5 | 5 | 5 | 5 | 5 | 5 | 5 | 5 | 5 | 5 | 5 | 5 | 5 | 5 | 5 | 5 | 5 | 5 |
| Madagascar | 5 | 5 | 5 | 5 | 5 | 5 | 5 | 5 | 5 | 5 | 5 | 5 | 5 | 5 | 5 | 5 | 5 | 5 | 5 | 5 | 5 | 5 | 5 | 5 | 5 | 5 | 5 | 5 | 5 | 5 | 5 | 5 |
| Malawi | 5 | 5 | 5 | 5 | 5 | 5 | 5 | 5 | 5 | 2 | 2 | 2 | 5 | 5 | 5 | 5 | 5 | 2 | 2 | 2 | 2 | 2 | 2 | 2 | 2 | 2 | 2 | 2 | 2 | 2 | 2 | 2 |
| Mauritius | 2 | 2 | 2 | 2 | 2 | 2 | 2 | 2 | 2 | 2 | 2 | 2 | 2 | 2 | 2 | 2 | 2 | 2 | 2 | 2 | 2 | 2 | 4 | 4 | 4 | 4 | 4 | 4 | 4 | 4 | 4 | 4 |
| Mozambique | 5 | 5 | 5 | 5 | 5 | 5 | 5 | 5 | 5 | 5 | 5 | 5 | 5 | 5 | 5 | 5 | 5 | 5 | 5 | 5 | 5 | 5 | 5 | 5 | 5 | 5 | 5 | 5 | 5 | 5 | 5 | 5 |
| Rwanda | 5 | 5 | 5 | 5 | 5 | 5 | 5 | 5 | 5 | 5 | 5 | 5 | 5 | 5 | 5 | 5 | 5 | 5 | 5 | 5 | 5 | 5 | 5 | 5 | 5 | 5 | 5 | 5 | 5 | 5 | 5 | 5 |
| Seychelles | 4 | 4 | 4 | 4 | 4 | 4 | 4 | 4 | 4 | 4 | 4 | 4 | 4 | 4 | 4 | 4 | 4 | 4 | 4 | 4 | 4 | 4 | 4 | 4 | 4 | 4 | 4 | 4 | 4 | 4 | 4 | 4 |
| Somalia | 5 | 5 | 5 | 5 | 5 | 5 | 5 | 5 | 5 | 5 | 5 | 5 | 5 | 5 | 5 | 5 | 5 | 5 | 5 | 5 | 5 | 5 | 5 | 5 | 5 | 5 | 5 | 5 | 5 | 5 | 5 | 5 |
| United Republic of Tanzania | 5 | 5 | 5 | 5 | 5 | 5 | 5 | 5 | 5 | 5 | 5 | 5 | 5 | 5 | 5 | 5 | 5 | 5 | 5 | 5 | 5 | 5 | 5 | 5 | 5 | 5 | 5 | 5 | 5 | 5 | 5 | 5 |
| Uganda | 5 | 5 | 5 | 5 | 5 | 5 | 5 | 5 | 5 | 5 | 5 | 5 | 5 | 5 | 5 | 5 | 5 | 5 | 5 | 5 | 5 | 5 | 5 | 5 | 5 | 5 | 5 | 5 | 5 | 5 | 5 | 5 |
| Zambia | 5 | 5 | 5 | 5 | 5 | 5 | 5 | 5 | 5 | 5 | 5 | 5 | 5 | 5 | 5 | 5 | 5 | 5 | 5 | 5 | 5 | 5 | 5 | 5 | 5 | 5 | 5 | 5 | 5 | 5 | 5 | 5 |
| Botswana | 2 | 2 | 2 | 2 | 2 | 2 | 2 | 2 | 2 | 2 | 2 | 2 | 2 | 2 | 2 | 2 | 2 | 2 | 2 | 2 | 2 | 2 | 2 | 2 | 2 | 2 | 2 | 2 | 2 | 2 | 2 | 2 |
| Lesotho | 5 | 5 | 5 | 5 | 5 | 5 | 5 | 5 | 5 | 5 | 5 | 5 | 5 | 5 | 5 | 5 | 5 | 5 | 5 | 5 | 5 | 5 | 5 | 5 | 5 | 5 | 5 | 5 | 5 | 5 | 5 | 5 |
| Namibia | 3 | 3 | 3 | 3 | 3 | 3 | 5 | 5 | 5 | 5 | 5 | 5 | 5 | 5 | 5 | 5 | 5 | 5 | 5 | 5 | 5 | 5 | 5 | 5 | 5 | 5 | 5 | 5 | 5 | 5 | 3 | 5 |
| South Africa | 2 | 2 | 2 | 2 | 2 | 2 | 2 | 2 | 2 | 2 | 2 | 2 | 2 | 2 | 2 | 2 | 2 | 2 | 2 | 2 | 2 | 2 | 2 | 2 | 2 | 2 | 2 | 2 | 2 | 2 | 2 | 5 |
| Eswatini | 5 | 5 | 5 | 5 | 5 | 5 | 5 | 5 | 5 | 5 | 5 | 5 | 5 | 5 | 5 | 5 | 5 | 5 | 5 | 5 | 5 | 5 | 5 | 5 | 5 | 5 | 5 | 5 | 5 | 5 | 5 | 5 |
| Zimbabwe | 3 | 3 | 3 | 3 | 3 | 3 | 3 | 3 | 3 | 3 | 3 | 3 | 3 | 3 | 3 | 3 | 3 | 3 | 3 | 3 | 3 | 3 | 3 | 3 | 3 | 3 | 3 | 3 | 3 | 3 | 3 | 3 |
| Benin | 5 | 5 | 5 | 5 | 5 | 5 | 5 | 5 | 5 | 5 | 5 | 5 | 5 | 5 | 5 | 5 | 5 | 5 | 5 | 5 | 5 | 5 | 5 | 5 | 5 | 5 | 5 | 5 | 5 | 5 | 5 | 5 |
| Burkina Faso | 5 | 5 | 5 | 5 | 5 | 5 | 5 | 5 | 5 | 5 | 5 | 5 | 5 | 5 | 5 | 5 | 5 | 5 | 5 | 5 | 5 | 5 | 5 | 5 | 5 | 5 | 5 | 5 | 5 | 5 | 5 | 5 |
| Cameroon | 5 | 5 | 5 | 5 | 5 | 5 | 5 | 5 | 5 | 5 | 5 | 5 | 5 | 5 | 5 | 5 | 5 | 5 | 5 | 5 | 5 | 5 | 5 | 5 | 5 | 5 | 5 | 5 | 5 | 5 | 5 | 5 |
| Cabo Verde | 5 | 5 | 5 | 5 | 5 | 5 | 5 | 5 | 5 | 5 | 5 | 5 | 5 | 5 | 5 | 5 | 5 | 5 | 5 | 5 | 5 | 5 | 5 | 5 | 5 | 5 | 5 | 5 | 5 | 5 | 5 | 5 |
| Chad | 5 | 5 | 5 | 5 | 5 | 5 | 5 | 5 | 5 | 5 | 5 | 5 | 5 | 5 | 5 | 5 | 5 | 5 | 5 | 5 | 5 | 5 | 5 | 5 | 5 | 5 | 5 | 5 | 5 | 5 | 5 | 5 |
| Côte d'Ivoire | 3 | 3 | 3 | 3 | 3 | 3 | 3 | 3 | 3 | 3 | 3 | 3 | 3 | 3 | 3 | 3 | 3 | 3 | 3 | 3 | 3 | 3 | 3 | 5 | 5 | 5 | 5 | 5 | 5 | 5 | 3 | 3 |
| Gambia | 5 | 5 | 5 | 5 | 5 | 5 | 5 | 5 | 5 | 5 | 5 | 5 | 5 | 5 | 5 | 5 | 5 | 5 | 5 | 5 | 5 | 5 | 5 | 5 | 5 | 5 | 5 | 5 | 5 | 5 | 5 | 5 |
| Ghana | 5 | 5 | 5 | 5 | 5 | 5 | 5 | 5 | 5 | 5 | 5 | 5 | 5 | 5 | 5 | 5 | 5 | 5 | 5 | 5 | 5 | 5 | 5 | 5 | 5 | 5 | 5 | 5 | 5 | 5 | 5 | 5 |
| Guinea | 5 | 5 | 5 | 5 | 5 | 5 | 5 | 5 | 5 | 5 | 5 | 5 | 5 | 5 | 5 | 5 | 5 | 5 | 5 | 5 | 5 | 5 | 5 | 5 | 5 | 5 | 5 | 5 | 5 | 5 | 5 | 5 |
| Guinea-Bissau | 5 | 5 | 5 | 5 | 5 | 5 | 5 | 5 | 5 | 5 | 5 | 5 | 5 | 5 | 5 | 5 | 5 | 5 | 5 | 5 | 5 | 5 | 5 | 5 | 5 | 5 | 5 | 5 | 5 | 5 | 5 | 5 |
| Liberia | 5 | 5 | 5 | 5 | 5 | 5 | 5 | 5 | 5 | 5 | 5 | 5 | 5 | 5 | 5 | 5 | 5 | 5 | 5 | 5 | 5 | 5 | 5 | 5 | 5 | 5 | 5 | 5 | 5 | 5 | 5 | 5 |
| Mali | 3 | 3 | 3 | 3 | 3 | 3 | 3 | 3 | 3 | 3 | 3 | 3 | 3 | 3 | 3 | 3 | 3 | 3 | 3 | 3 | 3 | 3 | 3 | 3 | 3 | 3 | 3 | 3 | 3 | 3 | 3 | 3 |
| Mauritania | 3 | 3 | 3 | 3 | 3 | 3 | 3 | 3 | 3 | 3 | 3 | 3 | 3 | 3 | 3 | 3 | 3 | 3 | 3 | 3 | 3 | 3 | 3 | 3 | 3 | 3 | 3 | 3 | 3 | 3 | 3 | 3 |
| Niger | 3 | 3 | 3 | 3 | 3 | 3 | 3 | 3 | 3 | 5 | 5 | 5 | 5 | 5 | 5 | 5 | 5 | 5 | 5 | 5 | 5 | 5 | 5 | 5 | 5 | 5 | 5 | 5 | 5 | 5 | 5 | 3 |
| Nigeria | 5 | 5 | 5 | 5 | 5 | 5 | 5 | 5 | 5 | 5 | 5 | 5 | 5 | 5 | 5 | 5 | 5 | 5 | 5 | 5 | 5 | 3 | 3 | 3 | 3 | 3 | 3 | 3 | 3 | 3 | 3 | 3 |
| Sao Tome and Principe | 3 | 3 | 3 | 3 | 3 | 3 | 3 | 3 | 3 | 3 | 3 | 3 | 3 | 3 | 3 | 3 | 3 | 3 | 3 | 3 | 3 | 3 | 3 | 3 | 3 | 3 | 3 | 3 | 3 | 3 | 3 | 3 |
| Senegal | 3 | 3 | 3 | 3 | 3 | 3 | 3 | 3 | 3 | 3 | 3 | 3 | 3 | 3 | 3 | 3 | 3 | 3 | 3 | 3 | 3 | 5 | 5 | 5 | 5 | 5 | 5 | 5 | 5 | 5 | 5 | 3 |
| Sierra Leone | 5 | 5 | 5 | 5 | 5 | 5 | 5 | 5 | 5 | 5 | 5 | 5 | 5 | 5 | 5 | 5 | 5 | 5 | 5 | 5 | 5 | 5 | 5 | 5 | 5 | 5 | 5 | 5 | 5 | 5 | 5 | 5 |
| Togo | 5 | 5 | 5 | 5 | 5 | 5 | 5 | 5 | 5 | 5 | 5 | 5 | 5 | 5 | 5 | 5 | 5 | 5 | 5 | 5 | 5 | 5 | 5 | 5 | 5 | 5 | 5 | 5 | 5 | 5 | 5 | 5 |
| American Samoa | 4 | 4 | 4 | 4 | 4 | 4 | 4 | 4 | 4 | 4 | 4 | 4 | 4 | 4 | 4 | 4 | 4 | 4 | 4 | 4 | 4 | 4 | 4 | 4 | 4 | 4 | 4 | 4 | 4 | 4 | 4 | 4 |
| Bermuda | 5 | 5 | 5 | 5 | 5 | 5 | 5 | 5 | 5 | 5 | 5 | 5 | 5 | 5 | 5 | 5 | 5 | 5 | 5 | 5 | 5 | 5 | 5 | 5 | 5 | 5 | 5 | 5 | 5 | 5 | 5 | 5 |
| Cook Islands | 4 | 4 | 4 | 4 | 4 | 4 | 4 | 4 | 4 | 4 | 4 | 4 | 4 | 4 | 4 | 4 | 4 | 4 | 4 | 4 | 4 | 4 | 4 | 4 | 4 | 4 | 4 | 4 | 4 | 4 | 4 | 4 |
| Greenland | 1 | 1 | 1 | 1 | 1 | 1 | 1 | 1 | 1 | 1 | 1 | 1 | 1 | 1 | 1 | 1 | 1 | 1 | 1 | 1 | 1 | 1 | 1 | 1 | 1 | 1 | 1 | 1 | 1 | 1 | 1 | 1 |
| Guam | 4 | 4 | 4 | 4 | 4 | 4 | 4 | 4 | 4 | 4 | 4 | 4 | 4 | 4 | 4 | 4 | 4 | 4 | 4 | 4 | 4 | 4 | 4 | 4 | 4 | 4 | 4 | 4 | 4 | 4 | 4 | 4 |
| Monaco | 1 | 1 | 1 | 1 | 1 | 1 | 1 | 1 | 1 | 1 | 1 | 1 | 1 | 1 | 1 | 1 | 5 | 5 | 5 | 5 | 5 | 5 | 5 | 5 | 5 | 5 | 5 | 5 | 5 | 5 | 5 | 5 |
| Nauru | 4 | 4 | 4 | 4 | 4 | 4 | 4 | 4 | 4 | 4 | 4 | 4 | 4 | 4 | 4 | 4 | 4 | 4 | 4 | 4 | 4 | 4 | 4 | 4 | 4 | 4 | 4 | 4 | 4 | 4 | 4 | 4 |
| Niue | 4 | 4 | 4 | 4 | 4 | 4 | 4 | 4 | 4 | 4 | 4 | 4 | 4 | 4 | 4 | 4 | 4 | 4 | 4 | 4 | 4 | 4 | 4 | 4 | 4 | 4 | 4 | 4 | 4 | 4 | 4 | 4 |
| Northern Mariana Islands | 4 | 4 | 4 | 4 | 4 | 4 | 4 | 4 | 4 | 4 | 4 | 4 | 4 | 4 | 4 | 4 | 4 | 4 | 4 | 4 | 4 | 4 | 4 | 4 | 4 | 4 | 4 | 4 | 4 | 4 | 4 | 4 |
| Palau | 3 | 3 | 3 | 3 | 3 | 3 | 3 | 3 | 3 | 3 | 3 | 3 | 3 | 3 | 3 | 3 | 3 | 3 | 3 | 3 | 3 | 3 | 3 | 3 | 3 | 3 | 3 | 4 | 4 | 4 | 3 | 4 |
| Puerto Rico | 5 | 5 | 5 | 5 | 5 | 5 | 5 | 5 | 5 | 5 | 5 | 5 | 5 | 5 | 5 | 5 | 5 | 5 | 5 | 5 | 5 | 5 | 5 | 5 | 5 | 5 | 5 | 5 | 5 | 5 | 5 | 5 |
| Saint Kitts and Nevis | 5 | 5 | 5 | 5 | 5 | 5 | 5 | 5 | 5 | 5 | 5 | 5 | 5 | 5 | 5 | 5 | 5 | 5 | 5 | 5 | 5 | 5 | 5 | 5 | 5 | 5 | 5 | 5 | 5 | 5 | 5 | 5 |
| San Marino | 1 | 1 | 1 | 1 | 1 | 1 | 1 | 1 | 1 | 1 | 1 | 1 | 1 | 1 | 1 | 1 | 1 | 1 | 1 | 1 | 1 | 5 | 5 | 5 | 5 | 5 | 5 | 5 | 5 | 5 | 5 | 5 |
| Tokelau | 4 | 4 | 4 | 4 | 4 | 4 | 4 | 4 | 4 | 4 | 4 | 4 | 4 | 4 | 4 | 4 | 4 | 4 | 4 | 4 | 4 | 4 | 4 | 4 | 4 | 4 | 4 | 4 | 4 | 4 | 4 | 4 |
| Tuvalu | 4 | 4 | 4 | 4 | 4 | 4 | 4 | 4 | 4 | 4 | 4 | 4 | 4 | 4 | 4 | 4 | 4 | 4 | 4 | 4 | 4 | 4 | 4 | 4 | 4 | 4 | 4 | 4 | 4 | 4 | 4 | 4 |
| United States Virgin Islands | 5 | 5 | 5 | 5 | 5 | 5 | 5 | 5 | 5 | 5 | 5 | 5 | 5 | 5 | 5 | 5 | 5 | 5 | 5 | 5 | 5 | 5 | 5 | 5 | 5 | 5 | 5 | 5 | 5 | 5 | 5 | 5 |
| South Sudan | 5 | 5 | 5 | 5 | 5 | 5 | 5 | 5 | 5 | 5 | 5 | 5 | 5 | 5 | 5 | 5 | 5 | 5 | 5 | 5 | 5 | 5 | 5 | 5 | 5 | 5 | 5 | 5 | 5 | 5 | 5 | 5 |
| Sudan | 1 | 1 | 1 | 1 | 1 | 1 | 1 | 1 | 1 | 1 | 1 | 1 | 1 | 1 | 1 | 1 | 1 | 1 | 1 | 1 | 1 | 1 | 1 | 1 | 1 | 1 | 1 | 1 | 1 | 1 | 1 | 1 |

Note: 1: High Co-existence; 2: Middle Co-existence, 3: Low Co-existence, 4: Dementia-Dominant, 5: Mental Disorders-Dominant.

Supplementary Table 8: The number of countries with co-existence and cooccurrence patterns in 1990 and 2021

| Concurrent Pattern in 1990 | Co-existence pattern in 2021 | | | P | Cooccurrence pattern in 2021 | | | P |
| --- | --- | --- | --- | --- | --- | --- | --- | --- |
|  | Low | Middle | High |  | Low | Middle | High |  |
| Schizophrenia |  |  |  | <0.001 |  |  |  | <0.001 |
| Low | 21 (77.8%) | 0 (0%) | 0 (0%) |  | 7 (77.8%) | 0 (0%) | 0 (0%) |  |
| Middle | 0 (0%) | 9 (64.3%) | 0 (0%) |  | 0 (0%) | 10 (62.5%) | 0 (0%) |  |
| High | 0 (0%) | 0 (0%) | 20 (74.1%) |  | 0 (0%) | 0 (0%) | 16 (76.2%) |  |
| Depressive Disorders |  |  |  | <0.001 |  |  |  | <0.001 |
| Low | 11 (73.3%) | 0 (0%) | 0 (0%) |  | 9 (75.0%) | 0 (0%) | 0 (0%) |  |
| Middle | 0 (0%) | 9 (40.9%) | 0 (0%) |  | 0 (0%) | 10 (52.6%) | 0 (0%) |  |
| High | 0 (0%) | 0 (0%) | 14 (66.7%) |  | 0 (0%) | 0 (0%) | 16 (76.2%) |  |
| Bipolar Disorders |  |  |  | <0.001 |  |  |  | <0.001 |
| Low | 19 (95.0%) | 0 (0%) | 0 (0%) |  | 8 (88.9%) | 0 (0%) | 0 (0%) |  |
| Middle | 0 (0%) | 19 (76.0%) | 0 (0%) |  | 0 (0%) | 8 (72.7%) | 0 (0%) |  |
| High | 0 (0%) | 0 (0%) | 21 (84.0%) |  | 0 (0%) | 0 (0%) | 24 (92.3%) |  |
| Anxiety Disorders |  |  |  | <0.001 |  |  |  | <0.001 |
| Low | 26 (83.9%) | 1 (2.9%) | 0 (0%) |  | 27 (81.8%) | 0 (0%) | 0 (0%) |  |
| Middle | 1 (3.2%) | 28 (80.0%) | 0 (0%) |  | 1 (3.0%) | 25 (78.1%) | 1 (3.4%) |  |
| High | 0 (0%) | 0 (0%) | 26 (89.7%) |  | 0 (0%) | 0 (0%) | 27 (93.1%) |  |
| Eating Disorders |  |  |  | <0.001 |  |  |  | <0.001 |
| Low | 18 (64.3%) | 2 (6.5%) | 0 (0%) |  | 22 (73.3%) | 0 (0%) | 0 (0%) |  |
| Middle | 0 (0%) | 25 (80.6%) | 0 (0%) |  | 0 (0%) | 11 (57.9%) | 0 (0%) |  |
| High | 0 (0%) | 0 (0%) | 25 (83.3%) |  | 0 (0%) | 1 (5.3%) | 16 (80.0%) |  |
| ASD |  |  |  | <0.001 |  |  |  |  |
| Low | 27 (100%) | 0 (0%) | 0 (0%) |  | NA | NA | NA |  |
| Middle | 0 (0%) | 14 (70.0%) | 0 (0%) |  | NA | NA | NA |  |
| High | 0 (0%) | 0 (0%) | 17 (65.4%) |  | NA | NA | NA |  |
| ADHD |  |  |  | <0.001 |  |  |  |  |
| Low | 32 (88.9%) | 0 (0%) | 0 (0%) |  | NA | NA | NA |  |
| Middle | 0 (0%) | 30 (73.2%) | 0 (0%) |  | NA | NA | NA |  |
| High | 0 (0%) | 0 (0%) | 30 (93.8%) |  | NA | NA | NA |  |
| IDID |  |  |  | <0.001 |  |  |  |  |
| Low | 28 (87.5%) | 0 (0%) | 0 (0%) |  | NA | NA | NA |  |
| Middle | 0 (0%) | 17 (63.0%) | 3 (8.3%) |  | NA | NA | NA |  |
| High | 0 (0%) | 2 (7.4%) | 26 (72.2%) |  | NA | NA | NA |  |

Note: The table only shows the number and percentage of concurrent (co-existence or cooccurrence) countries,where values for dementia-dominant and mental disorders-dominant are not shown. Low, middle, and high co-existence/co-occurrence mean that the prevalence/incidence rates of both dementia and mental disorders are at low, middle, and high levels, respectively, in a country or territory. Low: Low Co-existence/Occurrence; Middle: Middle Co-existence/Occurrence; High: High Co-existence/Occurrence; NA: Not Available.

Supplementary Table 9. The total number of countries in 21 regions.

| **Region** | **The total number of countries** |
| --- | --- |
| Australasia | 2 |
| Tropical Latin America | 2 |
| East Asia | 3 |
| Southern Latin America | 3 |
| High-income North America | 3 |
| Andean Latin America | 3 |
| High-income Asia Pacific | 4 |
| South Asia | 5 |
| Central Sub-Saharan Africa | 6 |
| Southern Sub-Saharan Africa | 6 |
| Eastern Europe | 7 |
| Central Asia | 9 |
| Central Latin America | 9 |
| Southeast Asia | 13 |
| Central Europe | 13 |
| Eastern Sub-Saharan Africa | 15 |
| Oceania | 18 |
| Caribbean | 19 |
| Western Sub-Saharan Africa | 19 |
| North Africa and Middle East | 21 |
| Western Europe | 24 |
| Total | 204 |

Supplementary Table 10. SDI levels of 204 countries.

| Region Name | Location_name | SDI Index value in 2021 | SDI Level |
| --- | --- | --- | --- |
| East Asia | China | 0.72 | High middle |
| East Asia | Democratic People's Republic of Korea | 0.57 | Low middle |
| East Asia | Taiwan | 0.88 | High |
| Southeast Asia | Cambodia | 0.47 | Low middle |
| Southeast Asia | Indonesia | 0.66 | Middle |
| Southeast Asia | Lao People's Democratic Republic | 0.49 | Low middle |
| Southeast Asia | Malaysia | 0.74 | High middle |
| Southeast Asia | Maldives | 0.66 | Middle |
| Southeast Asia | Myanmar | 0.53 | Low middle |
| Southeast Asia | Philippines | 0.65 | Middle |
| Southeast Asia | Sri Lanka | 0.70 | Middle |
| Southeast Asia | Thailand | 0.68 | Middle |
| Southeast Asia | Timor-Leste | 0.45 | Low |
| Southeast Asia | Viet Nam | 0.62 | Middle |
| Oceania | Fiji | 0.67 | Middle |
| Oceania | Kiribati | 0.53 | Low middle |
| Oceania | Marshall Islands | 0.57 | Low middle |
| Oceania | Micronesia (Federated States of) | 0.59 | Low middle |
| Oceania | Papua New Guinea | 0.42 | Low |
| Oceania | Samoa | 0.59 | Low middle |
| Oceania | Solomon Islands | 0.43 | Low |
| Oceania | Tonga | 0.63 | Middle |
| Oceania | Vanuatu | 0.47 | Low middle |
| Central Asia | Armenia | 0.70 | Middle |
| Central Asia | Azerbaijan | 0.70 | Middle |
| Central Asia | Georgia | 0.73 | High middle |
| Central Asia | Kazakhstan | 0.72 | High middle |
| Central Asia | Kyrgyzstan | 0.61 | Low middle |
| Central Asia | Mongolia | 0.62 | Low middle |
| Central Asia | Tajikistan | 0.54 | Low middle |
| Central Asia | Turkmenistan | 0.68 | Middle |
| Central Asia | Uzbekistan | 0.66 | Middle |
| Central Europe | Albania | 0.71 | Middle |
| Central Europe | Bosnia and Herzegovina | 0.72 | High middle |
| Central Europe | Bulgaria | 0.76 | High middle |
| Central Europe | Croatia | 0.80 | High middle |
| Central Europe | Czechia | 0.83 | High |
| Central Europe | Hungary | 0.79 | High middle |
| Central Europe | North Macedonia | 0.75 | High middle |
| Central Europe | Montenegro | 0.80 | High middle |
| Central Europe | Poland | 0.81 | High |
| Central Europe | Romania | 0.77 | High middle |
| Central Europe | Serbia | 0.79 | High middle |
| Central Europe | Slovakia | 0.81 | High middle |
| Central Europe | Slovenia | 0.84 | High |
| Eastern Europe | Belarus | 0.78 | High middle |
| Eastern Europe | Estonia | 0.85 | High |
| Eastern Europe | Latvia | 0.83 | High |
| Eastern Europe | Lithuania | 0.86 | High |
| Eastern Europe | Republic of Moldova | 0.73 | High middle |
| Eastern Europe | Russian Federation | 0.81 | High middle |
| Eastern Europe | Ukraine | 0.76 | High middle |
| High-income Asia Pacific | Brunei Darussalam | 0.81 | High middle |
| High-income Asia Pacific | Japan | 0.87 | High |
| High-income Asia Pacific | Republic of Korea | 0.89 | High |
| High-income Asia Pacific | Singapore | 0.86 | High |
| Australasia | Australia | 0.84 | High |
| Australasia | New Zealand | 0.85 | High |
| Western Europe | Andorra | 0.87 | High |
| Western Europe | Austria | 0.85 | High |
| Western Europe | Belgium | 0.85 | High |
| Western Europe | Cyprus | 0.84 | High |
| Western Europe | Denmark | 0.90 | High |
| Western Europe | Finland | 0.86 | High |
| Western Europe | France | 0.84 | High |
| Western Europe | Germany | 0.90 | High |
| Western Europe | Greece | 0.79 | High middle |
| Western Europe | Iceland | 0.87 | High |
| Western Europe | Ireland | 0.87 | High |
| Western Europe | Israel | 0.81 | High middle |
| Western Europe | Italy | 0.81 | High middle |
| Western Europe | Luxembourg | 0.88 | High |
| Western Europe | Malta | 0.80 | High middle |
| Western Europe | Netherlands | 0.89 | High |
| Western Europe | Norway | 0.92 | High |
| Western Europe | Portugal | 0.75 | High middle |
| Western Europe | Spain | 0.77 | High middle |
| Western Europe | Sweden | 0.89 | High |
| Western Europe | Switzerland | 0.93 | High |
| Western Europe | United Kingdom | 0.86 | High |
| Southern Latin America | Argentina | 0.73 | High middle |
| Southern Latin America | Chile | 0.77 | High middle |
| Southern Latin America | Uruguay | 0.72 | High middle |
| High-income North America | Canada | 0.87 | High |
| High-income North America | United States of America | 0.86 | High |
| Caribbean | Antigua and Barbuda | 0.75 | High middle |
| Caribbean | Bahamas | 0.81 | High middle |
| Caribbean | Barbados | 0.75 | High middle |
| Caribbean | Belize | 0.61 | Low middle |
| Caribbean | Cuba | 0.67 | Middle |
| Caribbean | Dominica | 0.75 | High middle |
| Caribbean | Dominican Republic | 0.62 | Middle |
| Caribbean | Grenada | 0.67 | Middle |
| Caribbean | Guyana | 0.65 | Middle |
| Caribbean | Haiti | 0.45 | Low |
| Caribbean | Jamaica | 0.68 | Middle |
| Caribbean | Saint Lucia | 0.67 | Middle |
| Caribbean | Saint Vincent and the Grenadines | 0.64 | Middle |
| Caribbean | Suriname | 0.64 | Middle |
| Caribbean | Trinidad and Tobago | 0.77 | High middle |
| Andean Latin America | Bolivia (Plurinational State of) | 0.60 | Low middle |
| Andean Latin America | Ecuador | 0.67 | Middle |
| Andean Latin America | Peru | 0.66 | Middle |
| Central Latin America | Colombia | 0.66 | Middle |
| Central Latin America | Costa Rica | 0.70 | Middle |
| Central Latin America | El Salvador | 0.57 | Low middle |
| Central Latin America | Guatemala | 0.54 | Low middle |
| Central Latin America | Honduras | 0.51 | Low middle |
| Central Latin America | Mexico | 0.66 | Middle |
| Central Latin America | Nicaragua | 0.52 | Low middle |
| Central Latin America | Panama | 0.71 | Middle |
| Central Latin America | Venezuela (Bolivarian Republic of) | 0.60 | Low middle |
| Tropical Latin America | Brazil | 0.65 | Middle |
| Tropical Latin America | Paraguay | 0.65 | Middle |
| North Africa and Middle East | Algeria | 0.66 | Middle |
| North Africa and Middle East | Bahrain | 0.75 | High middle |
| North Africa and Middle East | Egypt | 0.60 | Low middle |
| North Africa and Middle East | Iran (Islamic Republic of) | 0.70 | Middle |
| North Africa and Middle East | Iraq | 0.66 | Middle |
| North Africa and Middle East | Jordan | 0.73 | High middle |
| North Africa and Middle East | Kuwait | 0.85 | High |
| North Africa and Middle East | Lebanon | 0.74 | High middle |
| North Africa and Middle East | Libya | 0.74 | High middle |
| North Africa and Middle East | Morocco | 0.56 | Low middle |
| North Africa and Middle East | Palestine | 0.63 | Middle |
| North Africa and Middle East | Oman | 0.77 | High middle |
| North Africa and Middle East | Qatar | 0.85 | High |
| North Africa and Middle East | Saudi Arabia | 0.81 | High |
| North Africa and Middle East | Syrian Arab Republic | 0.62 | Middle |
| North Africa and Middle East | Tunisia | 0.68 | Middle |
| North Africa and Middle East | Türkiye | 0.71 | High middle |
| North Africa and Middle East | United Arab Emirates | 0.85 | High |
| North Africa and Middle East | Yemen | 0.45 | Low |
| North Africa and Middle East | Afghanistan | 0.34 | Low |
| South Asia | Bangladesh | 0.49 | Low middle |
| South Asia | Bhutan | 0.48 | Low middle |
| South Asia | India | 0.58 | Low middle |
| South Asia | Nepal | 0.43 | Low |
| South Asia | Pakistan | 0.50 | Low middle |
| Central Sub-Saharan Africa | Angola | 0.48 | Low middle |
| Central Sub-Saharan Africa | Central African Republic | 0.31 | Low |
| Central Sub-Saharan Africa | Congo | 0.59 | Low middle |
| Central Sub-Saharan Africa | Democratic Republic of the Congo | 0.39 | Low |
| Central Sub-Saharan Africa | Equatorial Guinea | 0.66 | Middle |
| Central Sub-Saharan Africa | Gabon | 0.64 | Middle |
| Eastern Sub-Saharan Africa | Burundi | 0.29 | Low |
| Eastern Sub-Saharan Africa | Comoros | 0.48 | Low middle |
| Eastern Sub-Saharan Africa | Djibouti | 0.49 | Low middle |
| Eastern Sub-Saharan Africa | Eritrea | 0.40 | Low |
| Eastern Sub-Saharan Africa | Ethiopia | 0.36 | Low |
| Eastern Sub-Saharan Africa | Kenya | 0.52 | Low middle |
| Eastern Sub-Saharan Africa | Madagascar | 0.40 | Low |
| Eastern Sub-Saharan Africa | Malawi | 0.38 | Low |
| Southeast Asia | Mauritius | 0.72 | High middle |
| Eastern Sub-Saharan Africa | Mozambique | 0.33 | Low |
| Eastern Sub-Saharan Africa | Rwanda | 0.44 | Low |
| Southeast Asia | Seychelles | 0.73 | High middle |
| Eastern Sub-Saharan Africa | Somalia | 0.08 | Low |
| Eastern Sub-Saharan Africa | United Republic of Tanzania | 0.45 | Low |
| Eastern Sub-Saharan Africa | Uganda | 0.43 | Low |
| Eastern Sub-Saharan Africa | Zambia | 0.51 | Low middle |
| Southern Sub-Saharan Africa | Botswana | 0.64 | Middle |
| Southern Sub-Saharan Africa | Lesotho | 0.51 | Low middle |
| Southern Sub-Saharan Africa | Namibia | 0.62 | Low middle |
| Southern Sub-Saharan Africa | South Africa | 0.68 | Middle |
| Southern Sub-Saharan Africa | Eswatini | 0.59 | Low middle |
| Southern Sub-Saharan Africa | Zimbabwe | 0.48 | Low middle |
| Western Sub-Saharan Africa | Benin | 0.37 | Low |
| Western Sub-Saharan Africa | Burkina Faso | 0.28 | Low |
| Western Sub-Saharan Africa | Cameroon | 0.48 | Low middle |
| Western Sub-Saharan Africa | Cabo Verde | 0.53 | Low middle |
| Western Sub-Saharan Africa | Chad | 0.24 | Low |
| Western Sub-Saharan Africa | Côte d'Ivoire | 0.42 | Low |
| Western Sub-Saharan Africa | Gambia | 0.41 | Low |
| Western Sub-Saharan Africa | Ghana | 0.56 | Low middle |
| Western Sub-Saharan Africa | Guinea | 0.34 | Low |
| Western Sub-Saharan Africa | Guinea-Bissau | 0.35 | Low |
| Western Sub-Saharan Africa | Liberia | 0.35 | Low |
| Western Sub-Saharan Africa | Mali | 0.27 | Low |
| Western Sub-Saharan Africa | Mauritania | 0.50 | Low middle |
| Western Sub-Saharan Africa | Niger | 0.17 | Low |
| Western Sub-Saharan Africa | Nigeria | 0.50 | Low middle |
| Western Sub-Saharan Africa | Sao Tome and Principe | 0.50 | Low middle |
| Western Sub-Saharan Africa | Senegal | 0.41 | Low |
| Western Sub-Saharan Africa | Sierra Leone | 0.36 | Low |
| Western Sub-Saharan Africa | Togo | 0.41 | Low |
| Oceania | American Samoa | 0.73 | High middle |
| Caribbean | Bermuda | 0.82 | High |
| Oceania | Cook Islands | 0.78 | High middle |
| High-income North America | Greenland | 0.84 | High |
| Oceania | Guam | 0.80 | High middle |
| Western Europe | Monaco | 0.91 | High |
| Oceania | Nauru | 0.63 | Middle |
| Oceania | Niue | 0.73 | High middle |
| Oceania | Northern Mariana Islands | 0.78 | High middle |
| Oceania | Palau | 0.75 | High middle |
| Caribbean | Puerto Rico | 0.82 | High |
| Caribbean | Saint Kitts and Nevis | 0.76 | High middle |
| Western Europe | San Marino | 0.89 | High |
| Oceania | Tokelau | 0.69 | Middle |
| Oceania | Tuvalu | 0.58 | Low middle |
| Caribbean | United States Virgin Islands | 0.82 | High |
| Eastern Sub-Saharan Africa | South Sudan | 0.28 | Low |
| North Africa and Middle East | Sudan | 0.54 | Low middle |

Note: SDI: Socio-Demographic Index.

Supplementary Table 11. Total expected time spent in low, middle, and high co-existence and co-occurrence patterns of early-onset and later-onset of dementia with eight mental disorders over the past 31 years

| Mental Disorders | Co-existence (year) | | | | | | Cooccurrence (year) | | | | | |
| --- | --- | --- | --- | --- | --- | --- | --- | --- | --- | --- | --- | --- |
|  | Early-Onset of Dementia | | | Late-Onset of Dementia | | | Early-Onset of Dementia | | | Late-Onset of Dementia | | |
|  | Low | Middle | High | Low | Middle | High | Low | Middle | High | Low | Middle | High |
| Overall Mental Disorders (Reference) | 2.05 | 3.51 | 4.06 | 2.43 | 3.30 | 3.20 | 1.69 | 3.07 | 3.41 | 1.76 | 3.36 | 2.21 |
| Schizophrenia | 1.97 | 1.98 | 3.82 | 4.59 | 3.74 | 4.34 | 1.30 | 3.71 | 3.91 | 2.87 | 1.03 | 1.19 |
| Depressive Disorders | 2.48 | 3.11 | 3.4 | 2.30 | 3.72 | 2.14 | 1.71 | 3.12 | 3.37 | 1.76 | 3.36 | 2.21 |
| Bipolar Disorders | 3.19 | 4.86 | 2.28 | 1.87 | 2.43 | 4.93 | 0.99 | 3.37 | 1.82 | 1.47 | 1.93 | 5.03 |
| Anxiety Disorders | 3.8 | 4.83 | 3.01 | 5.42 | 3.61 | 4.59 | 3.43 | 3.59 | 3.21 | 5.26 | 3.37 | 4.65 |
| Eating Disorders | 3.02 | 4.73 | 3.16 | NA | NA | NA | 3.52 | 3.08 | 2.24 | NA | NA | NA |
| Autism Spectrum Disorders | 4.1 | 3.91 | 2.61 | 4.17 | 3.08 | 4.58 | NA | NA | NA | NA | NA | NA |
| Attention-Deficit/Hyperactivity Disorder | 3.87 | 5.62 | 4.25 | 5.66 | 5.82 | 4.79 | NA | NA | NA | NA | NA | NA |
| Idiopathic Developmental Intellectual Disability | 5.14 | 4.35 | 5.98 | 6.32 | 4.41 | 5.11 | NA | NA | NA | NA | NA | NA |

Note: Low, middle, and high co-existence/co-occurrence mean that the prevalence/incidence rates of both dementia and mental disorders are at low, middle, and high levels, respectively, in a country or territory. There is no prevalence and incidence data of eating disorders in population aged 65 and above. NA: Not Available.

Supplementary Table 12. Total expected time spent in low, middle, and high co-existence and co-occurrence patterns of dementia with eight mental disorders stratified by sex over the past 31 years

| Mental Disorders | Co-existence (year) | | | | | | Cooccurrence (year) | | | | | |
| --- | --- | --- | --- | --- | --- | --- | --- | --- | --- | --- | --- | --- |
|  | Male | | | Female | | | Male | | | Female | | |
|  | Low | Middle | High | Low | Middle | High | Low | Middle | High | Low | Middle | High |
| Overall Mental Disorders (Reference) | 1.39 | 2.24 | 3.44 | 3.09 | 3.30 | 5.22 | 1.21 | 2.80 | 2.29 | 2.66 | 3.55 | 3.18 |
| Schizophrenia | 6.08 | 2.74 | 3.16 | 3.98 | 3.00 | 4.34 | 1.06 | 2.3 | 2.84 | 4.00 | 3.74 | 4.04 |
| Depressive Disorders | 1.80 | 4.02 | 2.27 | 2.8 | 3.25 | 2.97 | 1.21 | 2.88 | 2.16 | 2.77 | 3.55 | 3.15 |
| Bipolar Disorders | 1.21 | 1.45 | 4.58 | 4.24 | 4.98 | 3.91 | 1.29 | 1.22 | 4.5 | 2.55 | 3.89 | 5.08 |
| Anxiety Disorders | 4.63 | 4.42 | 4.34 | 5.02 | 5.58 | 5.11 | 5.17 | 5.37 | 5.2 | 4.83 | 4.92 | 4.78 |
| Eating Disorders | 5.48 | 3.88 | 5.1 | 3.53 | 4.87 | 5.2 | 5.86 | 4.58 | 4.94 | 3.33 | 2.68 | 2.97 |
| Autism Spectrum Disorders | 3.18 | 2.34 | 4.73 | 3.82 | 2.47 | 1.77 | NA | NA | NA | NA | NA | NA |
| Attention-Deficit/Hyperactivity Disorder | 6.91 | 5.71 | 5.23 | 4.25 | 5.71 | 4.93 | NA | NA | NA | NA | NA | NA |
| Idiopathic Developmental Intellectual Disability | 4.42 | 3.77 | 5.33 | 5.34 | 4.66 | 5.43 | NA | NA | NA | NA | NA | NA |

Note: Low, middle, and high co-existence/co-occurrence mean that the prevalence/incidence rates of both dementia and mental disorders are at low, middle, and high levels, respectively, in a country or territory. There is no prevalence and incidence data of eating disorders in population aged 65 and above. NA: Not Available.

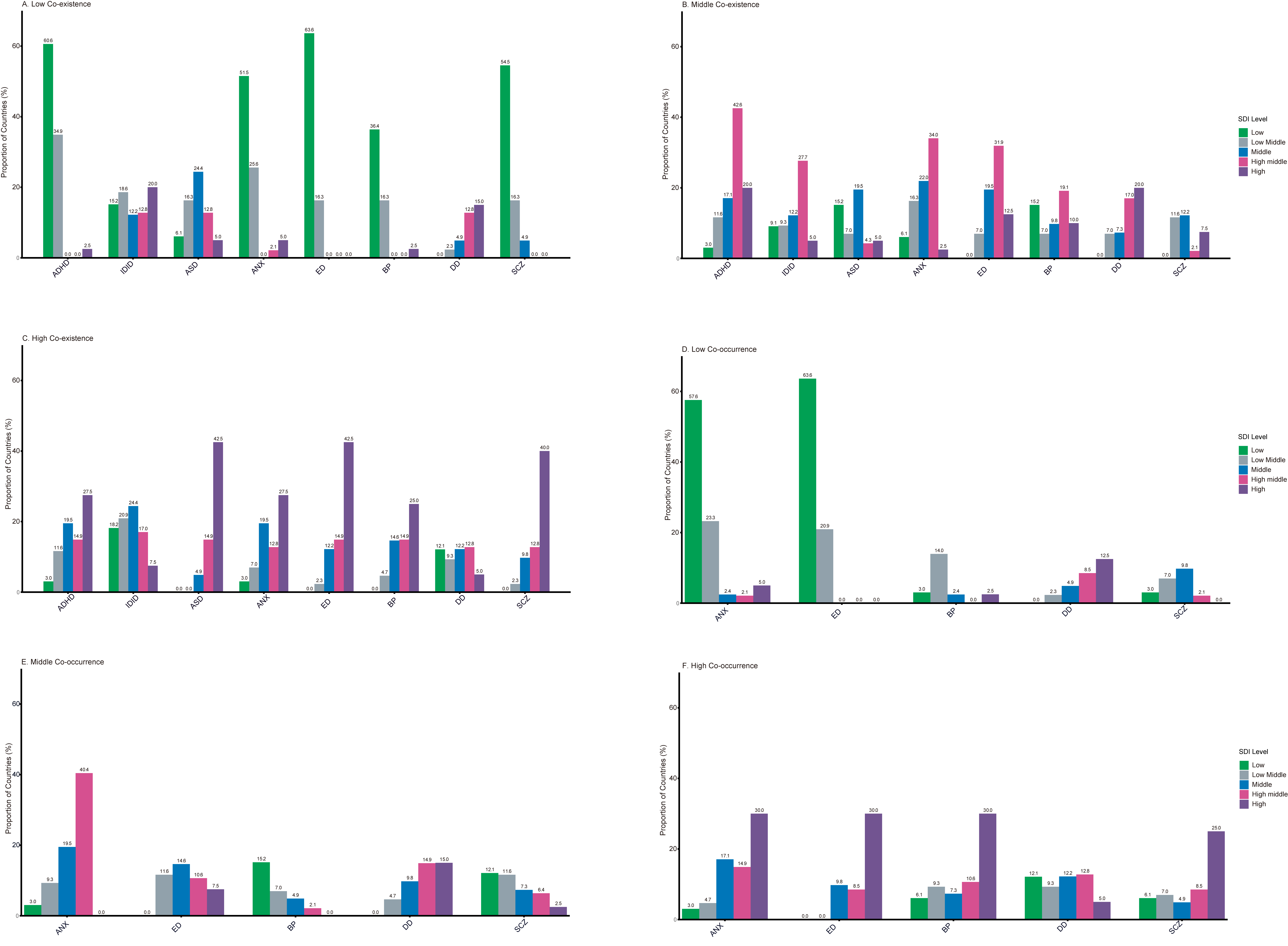

**Supplementary Figure 1: Proportion of countries with co-existence and co-occurrence across five SDI levels.**

Note: Low, Middle, or High co-existence/co-occurrence mean that the prevalence/incidence rates level of both dementia and mental disorders were at Low, Middle, or High levels, respectively, in a country or territory. SCZ: Schizophrenia; DD: Depressive Disorders; BD: Bipolar Disorders; ANX: Anxiety Disorders; ED: Eating Disorders; ASD: Autism Spectrum Disorders; ADHD: Attention-Deficit/Hyperactivity Disorder; IDID: Idiopathic Developmental Intellectual Disability.
